# Psychedelics distinctly alter brain entropy and complexity compared to psychostimulants

**DOI:** 10.64898/2026.01.15.26344193

**Authors:** K. Larsen, D.E. McCulloch, A.S. Olsen, F. Müller, M.E. Liechti, S. Borgwardt, F. Holze, L. Ley, P. Vizeli, A. Klaiber, A.M. Becker, B. Ozenne, M. Avram, P.M. Fisher

## Abstract

Classical psychedelics have regained interest for their potential to treat psychiatric and neurological disorders, and explore neural mechanisms supporting perception, cognition, and mood. Acute psychedelic effects have been linked to increases in brain complexity and entropy, but it is unclear if these changes are specific to psychedelics or reflect more general psychoactive effects. Here, we examined whether brain complexity and entropy metrics can identify features specific to the psychedelic state. Using resting-state fMRI from three placebo-controlled crossover trials (N=79; 255 sessions), we compared LSD, psilocybin, and mescaline with psychostimulants MDMA and d-Amphetamine. Compared to stimulants, psychedelics produced significant increases in meta-state complexity, short-timescale multiscale entropy, and dynamic conditional correlation entropy. Both drug classes increased Lempel–Ziv and spatial complexity, and decreased absolute modularity. Our findings highlight psychedelic-specific effects on brain signals that distinguish the acute psychedelic state from other psychoactive drug effects and may be relevant for understanding their therapeutic potential.

## Main

Psychedelic compounds have re-emerged as a major focus in psychiatry and neuroscience because of their potential to treat a range of psychiatric and neurological disorders, and enhance our understanding of brain processes underlying perception, cognition and mood. Classical psychedelics such as lysergic acid diethylamide (LSD), psilocybin, and mescaline induce profound changes in these domains, and although these drugs bind to a range of monoaminergic receptors, their characteristic psychoactive effects result primarily from stimulation of serotonin 2A (5-HT2A) receptors, as evidenced by the ability of selective antagonists to block these acute responses ^1,2^. At the same time, additional receptor systems may contribute to aspects of the subjective experience and longer-term effects, underscoring the complexity of psychedelic neuropharmacology ^3^.

LSD and psilocybin are currently in phase 3 clinical trials for the treatment of major depressive disorder and generalised anxiety disorder (LSD: NCT06941844, NCT06809595, NCT06741228, psilocybin: NCT03775200, NCT03866174) ^4–6^, and psilocybin has also demonstrated potential for treating neurological disorders such as cluster headache and Parkinson’s disease ^7–9^. The clinical potential and pronounced subjective effects of these compounds have motivated extensive neuroimaging research aimed at elucidating the underlying neural dynamics of these experiences and their potential relation to lasting therapeutic outcomes ^10^.

A prominent theory in the psychedelic neuroimaging literature is the Entropic Brain Hypothesis ^11^, which proposes that psychedelics shift brain dynamics toward a more flexible, high-entropy state, and thereby induce their characteristic subjective effects and therapeutic potential. Functional magnetic resonance imaging (fMRI) is widely used to study these acute effects of psychedelic drugs in humans, and several studies have applied quantitative measures of brain entropy to capture the complexity, variability, and spatial organisation of neural activity during psychedelic states. To date, more than a dozen studies have evaluated as many as a dozen unique brain entropy metrics ^11–23^. However, a recent comparative analysis found only limited correlation between many of these metrics in a cohort scanned multiple times acutely following psilocybin administration. This highlights the broad entropy landscape and motivates the need to identify the measure, or subset of measures, most strongly and reliably linked to psychedelic effects ^24^. Some entropy metrics have been consistently related to both subjective intensity and blood-measured drug levels, including the entropy of dynamic conditional correlation (DCC, a measure of the diversity of connectivity over time ^25^), multiscale sample entropy (MSSE, a measure of temporal signal variability ^12^), normalised spatial complexity (NSC, an entropy measure of spatial signal components ^23^), and meta-state complexity (MSC, a measure of brain state transitions ^21^). Other measures, such as brain modularity and Lempel-Ziv complexity (LZc), also show promising results in distinguishing various levels of consciousness and awareness ^26–31^.

Although these findings strongly link increased brain complexity and entropy to psychedelic effects, it remains an outstanding question whether these changes are specific to psychedelics. To address this, we leverage relevant comparator substances that produce psychoactive effects through shared monoaminergic mechanisms. 3,4-Methylenedioxymethamphetamine (MDMA) and d-Amphetamine are entactogens and stimulants, respectively, that produce their subjective effects by stimulating the release of monoamine neurotransmitters serotonin, dopamine, noradrenaline and oxytocin, specifically for MDMA ^32,33^. Although the R-enantiomer of MDMA has some affinity for the 5-HT2AR, this has been shown to mediate little of the subjective effects ^34^. MDMA in combination with psychotherapy has completed two phase 3 trials for the treatment of post-traumatic stress disorder ^35^, and d-Amphetamine is approved to treat attention-deficit hyperactivity disorder and narcolepsy. Although both drugs have established clinical profiles, their effects on brain entropy, as measured by fMRI, remain unstudied. Since these substances produce strong psychoactive effects through monoaminergic mechanisms, they serve as relevant comparators for evaluating the specificity of psychedelic-induced changes in brain entropy.

In this study, we provide the first comparison of classical psychedelics and psychoactive entactogen and stimulant drugs using several entropy metrics in the largest psychedelic neuroimaging dataset to date. These analyses provide critical insight into whether these entropy and complexity metrics represent specific signatures of the psychedelic state or are perhaps non-specific responses of psychoactive perturbation. We show that psychedelics specifically affect a subset of entropy and complexity metrics, especially in higher-order (transmodal) networks, increasing temporal variability at short timescales, decreasing it at longer timescales, and increasing the Shannon entropy of dynamic connectivity over time relative to stimulants. MSSE, DCC entropy, and meta-state complexity (MSC) captured the most pronounced psychedelic-specific effects. In contrast, modularity, LZc and NSC were similarly sensitive to stimulants. These findings highlight metrics that may be particularly informative for linking psychedelic-induced brain dynamics to altered states of consciousness and therapeutic outcomes.

## Results

We analysed resting-state fMRI data from 79 healthy participants across three randomised, placebo-controlled, crossover studies. Study 1 compared placebo and LSD (N=22, NCT02308969), Study 2 compared placebo, LSD, d-Amphetamine, and MDMA (N=25, NCT03019822), and Study 3 compared placebo, LSD, psilocybin, and mescaline (N=32, NCT04227756). A total of 255 nine-minute resting-state fMRI sessions acquired on a 3T Siemens scanner were included for analysis (see Supplementary Table S1 and Supplementary Fig. S1 for additional details). We considered 53 brain signal dynamics outcomes, derived from six entropy and complexity measures: metastate series complexity, modularity, Lempel-Ziv complexity, NSC, MSSE, and DCC entropy (see Fig. 1). While pointwise confidence intervals are reported, p-values were adjusted (p_FWER_) to control the family-wise error rate (FWER) over the 53 outcomes using the Holm–Bonferroni method (see Statistical Methods for details).

**Fig. 1.**
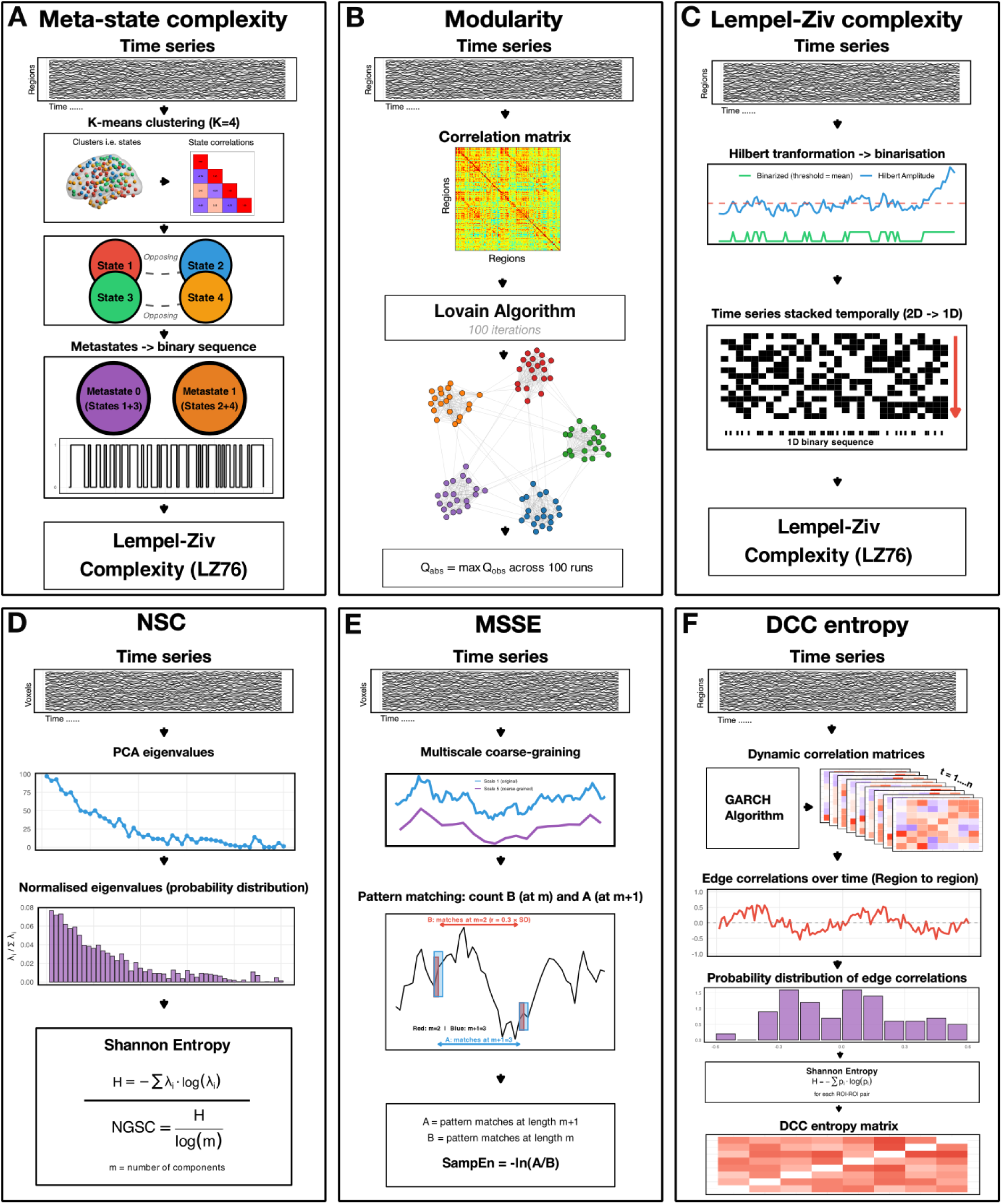
Schematic overview of the study metrics. **A) Meta-state complexity:** fMRI time series are clustered into four states using K-means with correlation distance, then grouped into two metastates composed of opposing centroids. Lempel-Ziv complexity using the 1976 algorithm (LZ76) is then calculated over time for the meta-state sequence. **B) Absolute modularity:** Correlation matrices from fMRI time series are analysed with the Louvain community detection algorithm, which is run 100 times to determine the optimal community structure. Absolute modularity (Q_abs) is the highest Q value observed across iterations and measures functional segregation; higher values indicate stronger community structure. **C) Lempel-Ziv complexity:** Time series are Hilbert transformed to extract amplitude envelopes. These are then binarised around the mean. The binary matrix is flattened temporally (2D→1D). LZ76 complexity is calculated on the resulting sequence. **D) Normalised spatial complexity (NSC):** Principal Component Analysis (PCA) is applied to the fMRI time series to extract eigenvalues representing temporospatial variance across components. These are normalised to form a probability distribution, and Shannon entropy is calculated. **NSC** is then normalised by log(m), where m is the number of components. **E) Multiscale Sample Entropy (MSSE):** The time series data is analysed at multiple levels of detail (scale 1 (original) and scale 5 (temporally downsampled). At each level, sample entropy assesses pattern regularity by identifying similar patterns of length m=2 (count B) throughout the time series. Patterns are considered similar if their values differ by no more than 0.3 times the signal’s standard deviation (r=0.3×SD). It then checks how many of these patterns remain similar when extended by one point to m+1=3 (count A). Sample entropy is calculated as SampEn = -ln(A/B). **F) Dynamic conditional correlation (DCC) entropy:** The DCC algorithm estimates time-varying correlation matrices at each time point to capture dynamic functional connectivity using the Generalised Autoregressive Conditional Heteroskedasticity (GARCH) algorithm. For each region pair, a probability distribution of correlation values over time is constructed, and Shannon entropy is calculated for each distribution. This produces a matrix that shows the temporal variability of functional connectivity between each region pair.

### Psychedelic-specific and non-specific effects on whole-brain measures

We first compared the effects of psychedelics and stimulants on whole-brain measures of brain signal entropy, complexity, and absolute modularity (Fig. 2, Table S2). Psychedelics significantly increased MSC compared to stimulants (β [95% CI] = 0.07 [0.04, 0.1]; p_FWER_ = 0.003). Psychedelics and stimulants produced numerically opposite effects relative to placebo, i.e., a positive effect of psychedelics and a negative effect of stimulants (Fig. 1B). There were no significant differences between drug classes for absolute modularity (β [95% CI] = −0.04 [-0.07, -0.1], p_FWER_ = 0.23), NSC (β [95% CI] = −0.007 [-0.01, 0.001], p_FWER_ = 1), or LZc (β [95% CI] = 0.003 [-0.001, 0.007], p_FWER_ = 1). Both drug classes increased NSC and LZc and decreased absolute modularity compared to placebo. For a detailed breakdown of individual drug effects and class comparisons, see Supplementary Figure S2 and Supplementary Tables S6-S9. Overall, group-level effects were consistent across drug classes, except for MSC, where MDMA and d-Amphetamine showed opposite effects.

**Fig. 2.**
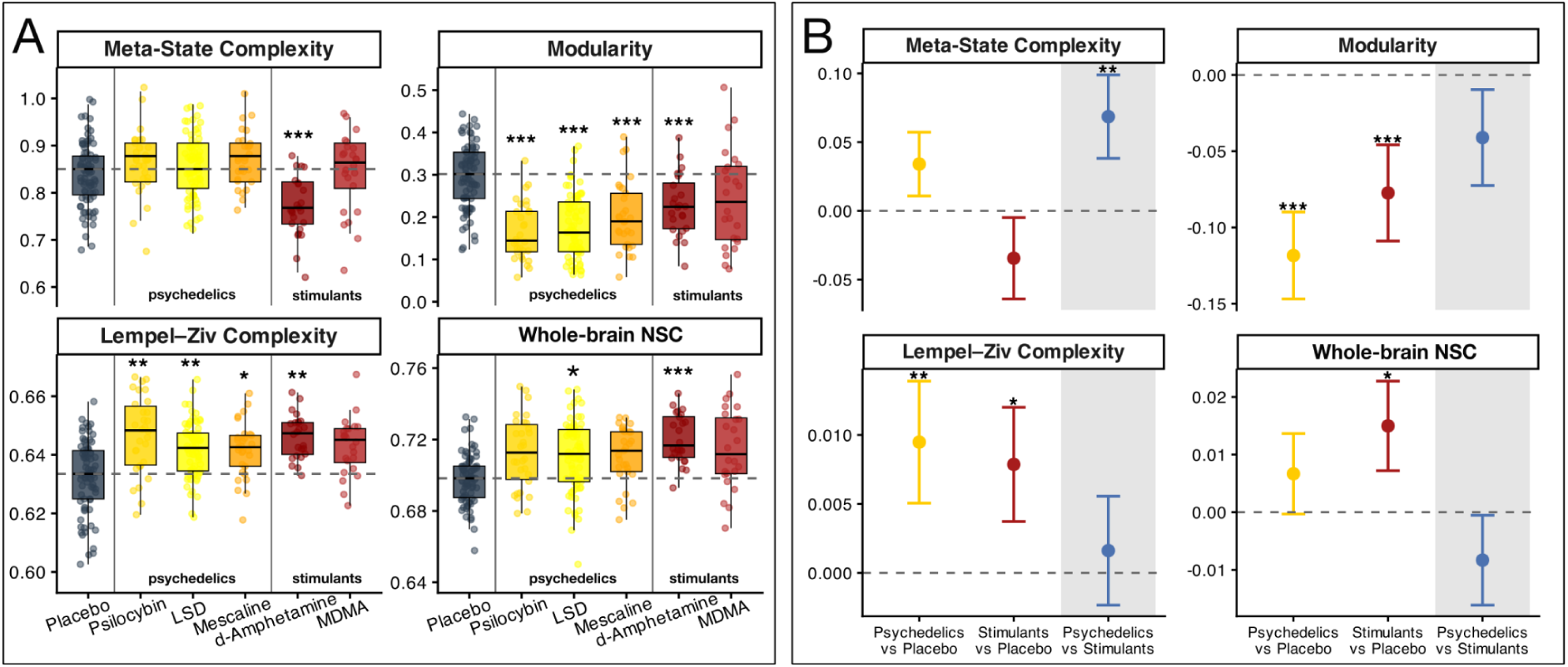
Differential effects of psychedelics and stimulants on whole-brain entropy, complexity, and modularity. **(A)** Box plots showing raw values for four whole-brain measures across drug conditions. Data include placebo (grey), psychedelics (shades of yellow), and stimulants (shades of red). Box plots display median, interquartile range, and individual data points. The horizontal dotted line indicates the median value for the placebo condition. **(B)** Forest plots displaying beta estimates and unadjusted 95% confidence intervals for three contrasts between drug classes: psychedelics vs. placebo (yellow), stimulants vs. placebo (red), and psychedelics vs. stimulants (primary comparison of interest, blue). Asterisks denote statistical significance (*p_FWER_ < 0.05, **p_FWER_ < 0.01, ***p_FWER_ < 0.001), adjusted for 53 outcomes for any given contrast.

### Psychedelic-specific effects on network entropy and complexity

Next, we applied the Schaefer-200 atlas with the Yeo-7 networks to compare psychedelic and stimulant effects on three network-level entropy and complexity measures: MSSE, network-level NSC, and DCC entropy. These capture temporal signal variability, spatiotemporal signal uniformity and connectivity dynamics, respectively. We report MSSE values for scale 1 and scale 5 only, as these have previously been linked to acute psilocybin effects ^24^.

#### Multiscale sample entropy

We pooled and compared regional MSSE across the Yeo-7 functional networks at short (Scale 1, 3.6-5.4 seconds) and long (Scale 5, 18-27 seconds) timescales (Fig. 3, Table S3). At scale 1, psychedelics induced higher MSSE than stimulants in transmodal networks, including the control (β [95% CI] = 0.01 [0.008, 0.017], p_FWER_ < 0.001), default mode (β [95% CI] = 0.015 [0.01, 0.02], p_FWER_ < 0.001), dorsal attention (β [95% CI] = 0.01 [0.007, 0.017], p_FWER_ < 0.001) and ventral attention networks (β [95% CI] = 0.007 [0.002, 0.011], p_FWER_ = 0.07). MSSE in unimodal networks, i.e., somatomotor and visual networks, increased for both psychedelics and stimulants; there was not a statistically significant difference between drug classes. At the long timescale, psychedelics induced lower MSSE relative to stimulants in transmodal networks, with statistically significant reductions in the control (β [95% CI] = −0.03 [−0.04, −0.02], p_FWER_ < 0.001), default mode (β [95% CI] = −0.03 [−0.04, −0.01], p_FWER_ = 0.003), and ventral attention networks (β [95% CI] = −0.02 [−0.03, −0.01], p_FWER_ = 0.02). No significant differences were observed for the dorsal attention, limbic, somatomotor, or visual networks. Supplementary Figure S3 shows full results for individual drug contrasts, and Supplementary Table S10 provides the complete set of regional and timescale results for all contrasts, networks, and scales.

**Fig 3.**
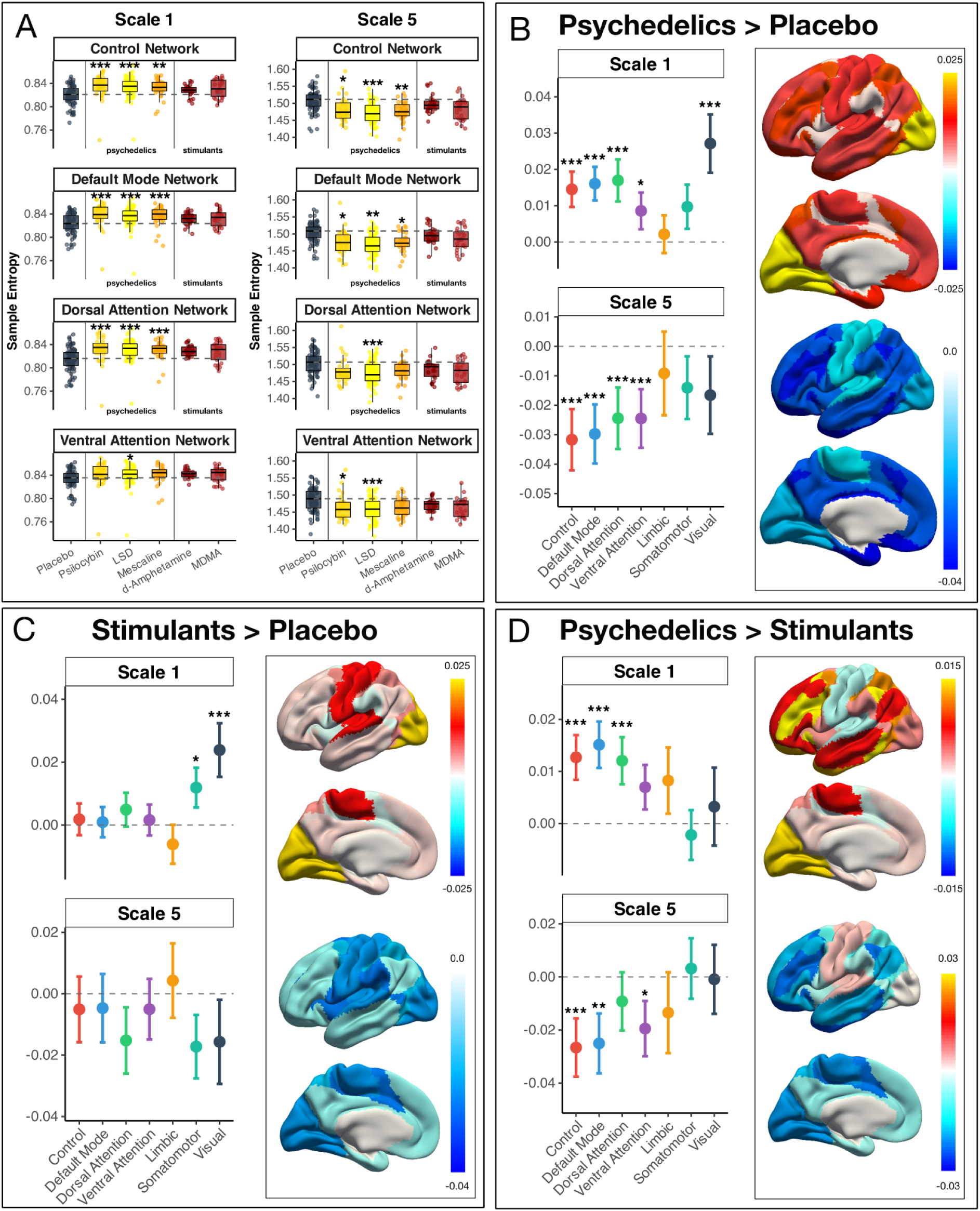
Psychedelics induce scale-dependent effects on multiscale sample entropy within transmodal networks. **(A)** Boxplots showing raw MSSE values at Scale 1 and Scale 5 across the transmodal brain networks. Data include placebo (grey), psychedelics (yellow), and stimulants (red). Box plots display median, interquartile range, and individual data points. **(B-D)** Forest plots displaying beta estimates and unadjusted 95% confidence intervals for three contrasts: (B) psychedelics vs. placebo, (C) stimulants vs. placebo, and (D) psychedelics vs. stimulants. Asterisks denote statistical significance (*p_FWER_ < 0.05, **p_FWER_ < 0.01, ***p_FWER_ < 0.001), adjusted for 53 outcomes for any given contrast.

#### Network-level normalised spatial complexity

We compared network-level NSC across the Yeo-7 networks between drug classes (Fig. 4, Table S4). Psychedelics and stimulants increased NSC compared to placebo in all networks except the limbic network (Fig. 3B–C). We did not observe significant differences between the two drug classes in transmodal networks (all p_FWER_ > 0.2), nor in the limbic network (p_FWER_ = 0.9). However, psychedelics showed significantly lower NSC than stimulants in the somatomotor network (β [CI 95%] = −0.04 [−0.06, −0.03], p_FWER_ < 0.001). Supplementary Figure S4 and Table S11 present all results for individual drugs compared to placebo, as well as within and between drug classes.

**Fig. 4.**
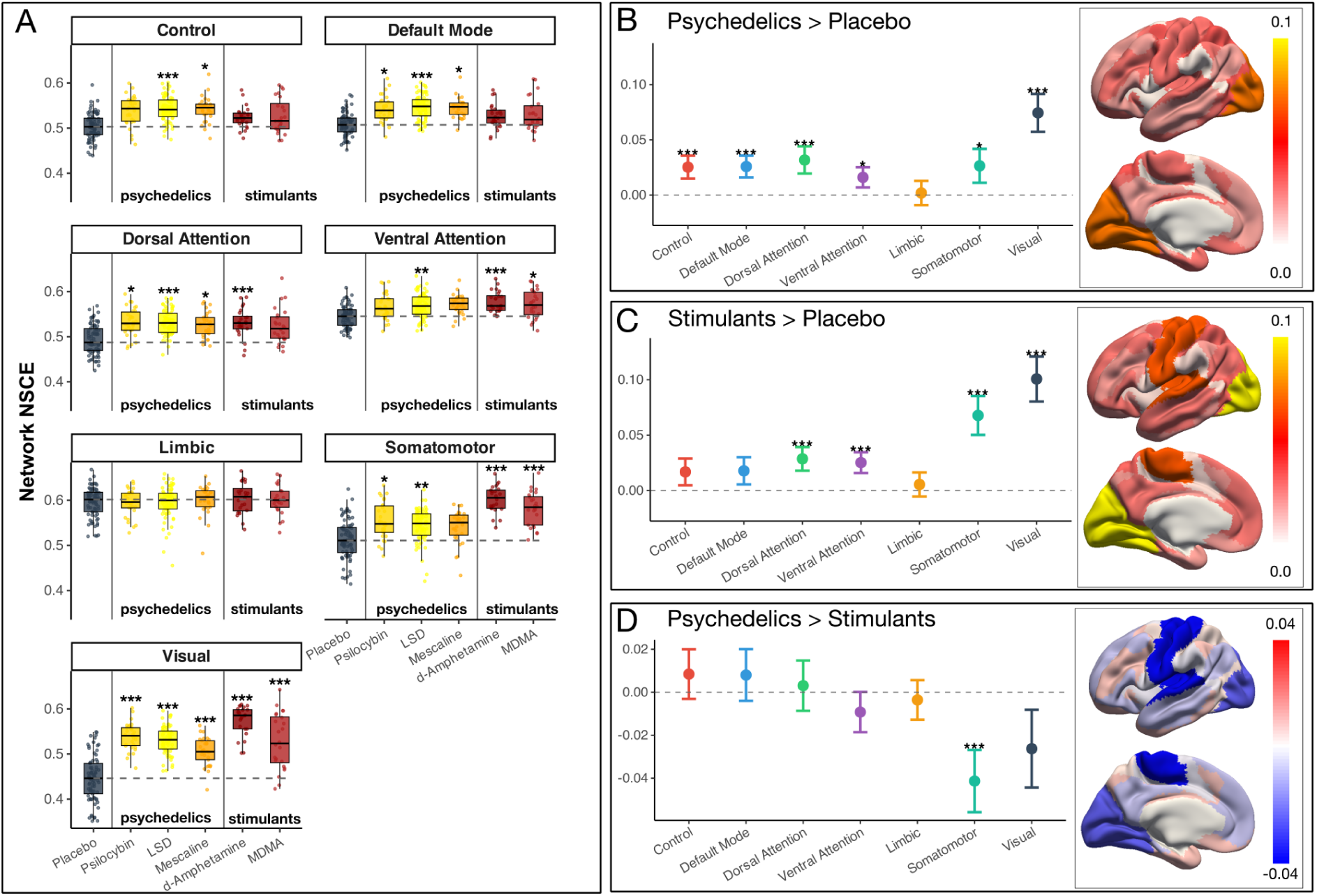
Network-level normalised spatial complexity (NSC) across drug conditions and brain networks. **(A)** Box plots showing NSC values for psychedelics (yellow) and stimulants (red) compared to placebo (grey) across seven brain networks. Individual data points are overlaid on box plots. Dashed horizontal lines indicate median placebo baseline levels for reference. **(B-D)** Brain surface maps displaying effect sizes for pairwise comparisons: **(B)** Psychedelics vs. Placebo, **(C)** Stimulants vs. Placebo, and **(D)** Psychedelics vs. Stimulants. Colour bars indicate effect size magnitude, with warm colours (red/orange) representing positive effects and cool colours (blue) representing negative effects. Error bars in accompanying plots show unadjusted 95% confidence intervals. Asterisks denote statistical significance (*p_FWER_ < 0.05, **p_FWER_ < 0.01, ***p_FWER_ < 0.001), adjusted for 53 outcomes for any given contrast.

#### Dynamic conditional correlation entropy

Psychedelics induced statistically significantly higher DCC entropy compared to stimulants within several transmodal networks as well as between networks (Figure 5; Supplementary Table S5). Significant increases occurred within and between the control, default mode, ventral attention, limbic and visual networks, indicating greater temporal variability and cross-network coupling in systems involved primarily in higher-order cognitive and affective processing. Stimulants showed limited effects on most network pairs, except for a significant DCC entropy increase within the somatomotor and visual network. No drug class differences were found between the unimodal visual except within somatomotor (all p_FWER_ > 0.05). Full model estimates and single-drug contrasts are provided in Supplementary Figures S5–S7 and Supplementary Table S12.

**Fig. 5.**
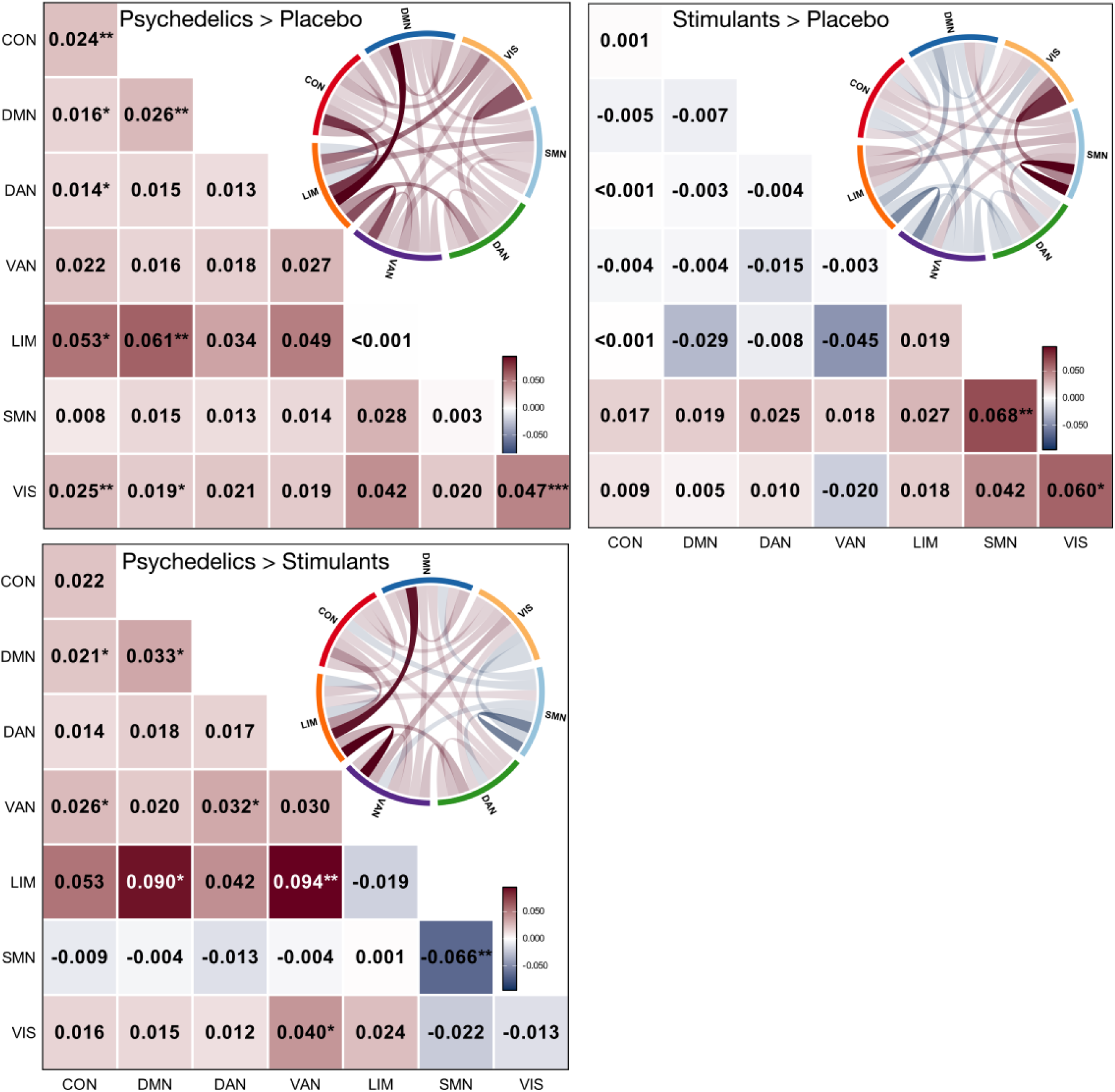
Psychedelic and stimulant effects on dynamic conditional correlation (DCC) entropy. Heat maps and chord diagrams show effect sizes for pairwise comparisons of within- and between-network DCC entropy: (A) Psychedelics vs. Placebo, (B) Stimulants vs. Placebo, and (C) Psychedelics vs. Stimulants. Heat maps display beta estimates using colour coding (warm colours indicate positive effects, cool colours indicate negative effects). Asterisks indicate statistical significance (*p_FWER_ < 0.05, **p_FWER_ < 0.01, ***p_FWER_ < 0.001), adjusted for 53 outcomes for any given contrast. Chord diagrams illustrate the magnitude and direction of the network effects, with line transparency proportional to effect size, i.e., stronger effects are more opaque. Network abbreviations: CON (Control), DMN (Default Mode), DAN (Dorsal Attention), VAN (Ventral Attention/Salience), LIM (Limbic), SMN (Somatomotor), VIS (Visual). Colour bars show the effect size scale.

## Discussion

To identify neural signatures unique to psychedelics, it is essential to distinguish brain effects caused by psychedelics from those produced by other psychoactive drugs. Here, we present the first direct comparison of several psychedelic drugs and their effects on brain entropy, complexity, and modularity. Compared to stimulants, psychedelics induced distinct changes in brain MSC, MSSE, and DCC entropy. By contrast, absolute modularity, NSC, and LZc effects were similar between both drug classes, indicating a general sensitivity to psychoactive drugs rather than a psychedelic-specific effect. Notably, all metrics considered in our study were sensitive to psychedelic effects to some degree.

Psychedelics decreased absolute modularity, increased LZc, NSC, MSC, and MSSE at short timescales, decreased MSSE at longer timescales, and increased DCC entropy within and between transmodal and limbic networks. These findings suggest that psychedelics distinctly make brain activity more temporally variable, network interactions more flexible, and large-scale systems more dynamic, particularly within transmodal networks. Although these effects distinguish psychedelics from stimulants in several domains, they are not necessarily exclusive to psychedelics. Rather, our results provide a basis for refining our understanding of the psychedelic brain state, and future studies comparing additional pharmacological classes or altered states of consciousness will help further clarify psychedelic specificity.

## Psychedelic-specific effects

### Psychedelic-specific effects on meta-state complexity

MSC showed psychedelic-specific increases, whereas stimulants produced the opposite effect. This finding replicates and extends previous findings for psilocybin and LSD ^24^ and supports MSC as a marker of psychedelic-induced alterations in brain-state dynamics. As a measure of the irregularity of transitions between recurrent patterns of functional connectivity, MSC captures the temporal flexibility of large-scale network configurations. Higher MSC under psychedelics indicates increased irregularity in switching between brain states, which possibly reflects a more dynamic brain state.

Future work should investigate the composition of these meta-states in more detail, i.e., their spatial patterns, temporal stability, and potential links to subjective experiences or therapeutic effects. Such analyses might help to elucidate which patterns of network transitions underpin specific phenomenological aspects of the psychedelic state.

### Multiscale sample entropy and dynamic conditional correlations entropy provide complementary psychedelic-specific effects

MSSE and DCC entropy both showed strong psychedelic-specific effects. MSSE assesses the repeatability of neural signals at specific time scales. At scale 1, reflecting short-term fluctuations (3.6-5.4 seconds, i.e., 2-3 brain volumes), psychedelics produced higher MSSE values, indicating increased signal sporadicity. This observation aligns with the Entropic Brain Hypothesis as well as previous findings for LSD and psilocybin ^12,24^. By contrast, psychedelics decreased MSSE at scale 5. We have previously observed this timescale-specific effect following psilocybin and a non-significant, yet directionally similar effect after LSD ^24^. The specific psychedelic-induced decrease at scale 5 indicates that, once faster fluctuations are smoothed out, psychedelics produce more regular and predictable temporal dynamics, compared to both stimulants and placebo. Taken together, these findings curiously indicate that psychedelics increase short-term unpredictability while also constraining longer-term temporal network dynamics, creating a form of ‘structured chaos’ in which rapid local fluctuations coexist with regular, slower dynamics.

DCC entropy measures the width of the distribution of functional connectivity between brain networks over time, capturing fluctuations in coupling strength. Unlike static connectivity, which averages correlations across a session, DCC entropy reflects the dynamic flexibility of network interactions. Higher DCC entropy values indicate greater unpredictability and less stability in these connections, highlighting differences between stable and dynamic network organisation.

Psychedelics increased DCC entropy across transmodal networks, including within and between the control and default mode networks, between these networks and attention systems, and with limbic and visual regions. In contrast, stimulants produced minimal DCC changes in transmodal circuits, affecting only the somatomotor and visual networks. This distinction suggests that the DCC entropy of higher-order networks may represent a psychedelic-specific phenomenon and capture alterations in brain function that underlie their acute perceptual effects. This aligns with our previous observation following psilocybin administration ^24^. Notably, an earlier study reported no persisting change in DCC entropy one week after psilocybin administration ^20^, which may indicate that these effects are temporally constrained to the acute psychedelic state.

MSSE scale 1 and DCC entropy provide complementary information about how psychedelics affect brain entropy. Both metrics assess short-timescale neural dynamics and consistently show increased entropy. These findings suggest that psychedelics disrupt the neural scaffolding that maintains ordered signalling on short timescales, possibly reflecting a more variable and flexible dynamic brain state.

This disruption at short timescales is most evident in transmodal association networks, which integrate information across sensory and cognitive domains. These networks support higher-order functions such as self-referential thinking, executive control, and attentional coordination, all of which are strongly modulated during psychedelic experiences ^18^. The finding that these networks show psychedelic-specific changes in both temporal variability and DCC entropy connects transmodal engagement to the unique network dynamics induced by psychedelics.

## Psychedelic-sensitive effects

Psychedelics and stimulants both similarly increased LZc and NSCs. LZc measures signal compressibility by counting the number of distinct patterns in a time series; more unique patterns yield a higher LZc. This measure has shown reliable increases under psychedelics across fMRI, EEG, and MEG, including LSD, psilocybin, and N, N-Dimethyltryptamine (DMT) ^13,24,36–38^. The similar increases we observed with stimulants, and previous reports of increased LZc under ketamine ^37,39^, indicate that although LZc is sensitive to various psychoactive effects (e.g., increased wakefulness), it is not straightforwardly attributable to the unique aspects of the psychedelic experience.

NSC quantifies the uniformity of variance explained across spatial components of a time series. Higher NSC values reflect greater spatial diversity in activity patterns. This metric was recently used to demonstrate psilocybin- and LSD-induced effects in the human brain ^23,24^. In our data, both psychedelics and stimulants increased NSC across most networks, with stimulants showing larger effects in somatomotor and visual systems.

It should be noted that increased LZc has also been observed using other imaging modalities such as EEG and MEG studies with psychedelics ^38–40^. Future studies should directly compare psychedelics and stimulants using EEG or MEG to determine whether this non-specificity extends across modalities.

We also observed decreased absolute modularity under both psychedelics and stimulants. Modularity measures how brain regions are organised into distinct communities; lower values indicate a shift toward more global network integration. This finding replicates a previous report of acute psilocybin-induced reductions in modularity ^41^. However, similar decreases for stimulants suggest that absolute modularity is broadly sensitive to acute pharmacological effects and does not differentiate between psychedelic and stimulant drug classes. Another study has also reported sub-acute changes in normalised modularity ^39^, which differs from the absolute metric applied here. We also estimated normalised modularity and found that psychedelics paradoxically increased this measure, whereas stimulants produced a slight decrease (Supplementary Fig. S8A). Additional analyses showed that normalised modularity is highly sensitive to drug-induced changes in global connectivity, thereby complicating interpretation. Therefore, we focus on absolute modularity in the main text and present normalised analyses as supplementary results.

## Drug class grouping

LSD, psilocybin, and mescaline are classified as classical serotonergic psychedelics because they produce their psychoactive effects via agonism at the 5-HT2A receptor, which sets them apart from other psychoactive drugs. However, LSD has the broadest receptor-binding profile, and interacts with serotonergic, dopaminergic, and adrenergic sites ^42^. Psilocybin is also selective for other serotonergic receptors, including 5-HT2C and 5-HT1A ^42,43^. Mescaline shows lower off-target binding, but the affinity for the 5-HT2A receptor is lower, which necessitates higher doses ^44^. Despite these pharmacological differences, all three compounds produced consistent effects across our entropy metrics (Supplementary Figures S2–S7), suggesting a shared neurodynamic mechanism underlying the psychedelic state. The psychedelic-specific effects we present here provide a unifying framework for understanding macroscopic psychedelic action. Still, individual receptor and pharmacokinetic/dynamic profiles may fine-tune how these compounds translate into therapeutic selectivity ^3^ and future studies should link such distinctions to clinical efficacy, side-effect profiles, and target indication.

We grouped MDMA and d-Amphetamine as psychostimulants based on their shared monoaminergic mechanisms ^45^. Both drugs act primarily on monoamine transporters, increasing extracellular dopamine and norepinephrine; additionally, MDMA promotes serotonin release via transporter reversal and increases central oxytocin levels ^33^. Because these mechanisms produce robust changes in domains such as arousal, mood, and cognition without inducing the same perceptual effects characteristic of serotonergic psychedelics, they provide an appropriate pharmacological contrast for assessing the specificity of psychedelic-related entropy changes. Supplementary Figures S2–S7 show that both stimulants produced increases in LZc and NSC comparable to those observed with psychedelics. These results illustrate why treating MDMA and d-Amphetamine as a unified psychostimulant class is analytically informative. They captured non-psychedelic brain entropy effects, which help to clarify entropy and complexity metrics that respond more generally to psychoactive stimulation, and isolate effects that appear more specific to psychedelics.

## Limitations

Several methodological limitations should be considered. 1) Although data were pooled from three independent studies, identical imaging acquisition parameters and preprocessing pipeline, and similar inclusion criteria support comparability. Placebo and LSD effects (present in all studies) were similar across studies (Supplementary Table S15, Supplementary Figure SX). Nevertheless, our statistical models included ‘project’ as a covariate to account for any systematic differences. 2) The number of scan sessions varied by drug, with LSD contributing the largest number of sessions. To limit bias due to session prevalence, we first estimated single-session level effects; drug-class contrasts (e.g., psychedelics vs stimulants) were subsequently calculated as the average of drug-specific effects, not weighted by session or participant count. 3) Two of the three pooled studies used crossover designs with multiple psychoactive compounds administered in a randomised order. To address this, our analyses adjusted for session order and prior drug exposure. Although washout periods and randomisation were applied, residual or carryover effects may remain, and future studies using parallel-group designs or longer washout intervals could help to further clarify compound-specific effects. 4) Although all data were acquired using the same scanning protocol, preprocessing choices can affect entropy metrics ^24^. We used the same standardised preprocessing pipeline for consistency across all data; however, results may vary with other denoising or motion correction methods, and future studies could test the robustness of our findings across a range of different denoising strategies. 5) Regional coverage varied by session; we calculated measures only for session-specific regions passing quality control (see Methods and Supplementary Materials for details). This prioritises use of available data with the trade-off of some variability in regional sampling. 6) Although MSC and modularity showed psychedelic-sensitive effects, both measures present interpretive challenges. MSC is calculated from a shared state space across conditions. This may obscure drug-specific connectivity patterns and thus reduce specific network dynamics to a limited number of meta-states. This might attenuate spatial specificity and information about brain-state stability. Modularity is sensitive to analytical choices, including normalisation, handling of negative correlations, and variability in the community detection algorithm, as well as changes in overall connection strength distribution. Consequently, changes in modularity may reflect altered network topology, global integration, or partition stability, complicating straightforward interpretation (See supplementary Figure S8). Future studies should address these considerations through more detailed analyses of meta-state composition and by carefully evaluating how methodological choices influence modularity estimates.

## Conclusion

Here, we provide insight into potentially psychedelic-specific effects on brain entropy and complexity. MSC, MSSE, and DCC entropy showed significantly distinct effects vis-à-vis MDMA and d-Amphetamine. By contrast, absolute modularity, LZc, and NSC increased after both psychedelics and stimulants, indicating general sensitivity to psychoactive substances rather than a psychedelic-specific effect. Notably, psychedelic effects on MSSE were timescale-dependent, with increased dynamical variability at the short timescale, which reduced in the long timescale. This could indicate that psychedelics reorganise brain dynamics rather than producing a uniform increase in disorder. These findings refine the entropic brain hypothesis by identifying metrics and their spatial patterns that distinguish psychedelic effects from those of other psychoactive drugs. Our findings highlight potentially psychedelic-specific effects on brain signals that may relate to the acute subjective experience and perhaps its promising therapeutic outcomes.

## Methods

### Ethics information

This research complies with all relevant ethical regulations and was approved by the Ethics Committee of Northwest Switzerland (EKNZ). Informed consent was obtained from all participants before enrollment. Our study draws on data from three clinical trials registered at ClinicalTrials.gov (registration numbers for projects one, two and three: P1: NCT02308969, P2: NCT03019822, P3: NCT04227756), involving the following sessions across the projects: placebo (N=79), 100 µg LSD (N=77), 20 mg psilocybin (N=31), 300/500 mg mescaline (N=32), 40 mg d-Amphetamine (N=25), and 125 mg MDMA (N=25), combined N=79 unique participants with fMRI data. All MRI data were collected using a 3T Siemens scanner at the University Hospital, Basel, Switzerland, in accordance with the Declaration of Helsinki. All participants received financial remuneration for their participation.

### Magnetic resonance imaging

Brain scanning across the three projects was performed on a Siemens 3 Tesla MRI Magnetom Prisma, using a 20-channel phased array radio frequency head coil. Anatomical images were acquired using a T1-weighted magnetisation prepared rapid acquisition gradient (MPRAGE) sequence (field of view: 256×256 mm; resolution: 1×1×1 mm; repetition time: 2000 ms; echo time: 3.37 ms; flip angle: 8°; bandwidth: 147 Hz/pixel). The BOLD fMRI acquisition was based on a T2*-weighted echo planar imaging sequence (repetition time: 1800 ms, echo time: 28 ms, bandwidth: 2442 Hz/pixel, field of view: 224 x 224 mm, resolution: 3.5 x 3.5 mm, slice thickness: 3.5 mm (0.5 mm gap), interleaved slice acquisition, number of axial slices: 35, in-plane matrix: 64 x 64, scan time: 9 minutes, number of volumes acquired: 300). Each participant’s head was fixed using two foam wedges to restrict motion. During the scan, participants were instructed to close their eyes and not to fall asleep.

### Pre-processing steps

MRI data were preprocessed using the Configurable Pipeline for the Analysis of Connectomes (version 1.7.1, https://fcp-indi.github.io/) with the default preconfigured pipeline (https://fcp-indi.github.io/docs/v1.8.3/user/pipelines/preconfig). Preprocessing included slice-timing correction, motion correction, scrubbing (FD Jenkinson <0.2), intensity normalisation, nuisance signal regression followed by bandpass filtering (0.01–0.1 Hz), registration to anatomical space, and normalisation to a 3 mm isotropic voxel size in Montreal Neurological Institute (MNI) space using FSL FLIRT/FNIRT. The normalised images were smoothed with a 5 mm full-width at half-maximum isotropic Gaussian kernel. Nuisance signal regression used a unified multiple linear regression model with component-based noise correction (aCompCor) to reduce physiological artefacts ^46^. Five components, each from white matter (WM) and cerebrospinal fluid (CSF) signals, were regressed out. Head motion effects were regressed using the Friston 24-parameter model, which included six motion parameters from the current volume, six derivatives, and their twelve squared terms. Global signal regression (GSR) was not applied, as it has been shown to influence substance-induced changes in functional connectivity distinctly ^47^.

### Atlas preparation

We used the Schaefer 200-parcellation 7-network atlas ^48^, which divides the cortex into 200 functionally distinct regions within 7 large-scale networks. The atlas was resampled to 3mm isotropic resolution to match our functional data and combined to create a comprehensive brain parcellation with 200 distinct regions.

### Time series extraction

For each subject and session, we extracted regional time series from atlas-defined regions. Quality control was applied at both voxel and regional levels. Voxels with NaN values, zero values, or no signal variation were excluded. We then calculated the percentage of valid voxels per region. Next, we calculated the regional time series as the median across all valid voxels for each frame. Regions with fewer than 50% valid voxels across all sessions for a participant were excluded from further analysis. These steps were performed at the participant level to account for individual anatomical differences and artefacts.

### Quality control and data exclusion

We estimated head motion using framewise displacement and applied scrubbing for FD values greater than 0.2 mm. Sessions with more than 20% scrubbed volumes were excluded. Fourteen sessions from 11 participants were excluded: one participant had three sessions excluded, one had two sessions, and the others had one session each. Supplementary Figure 1 provides an overview of motion-related exclusions, and Supplementary Table 1 details the exclusion of sessions across drug conditions.

Across all our network analyses, estimates involving the limbic network are prone to additional uncertainty due to specific susceptibility to noise and signal dropout. This issue is most relevant for DCC entropy, in which within-limbic connectivity could not be estimated in 62 sessions (23%) because only a single limbic region survived quality control. Even when multiple limbic regions were available, coverage was generally poor, and between-network estimates involving the limbic system should be interpreted with caution. Full details of regional survival and quality control are provided in the Supplementary Materials table S13-S14.

### Entropy metrics

We computed brain complexity and entropy measures using the in-house Copenhagen Brain Entropy (CopBET) toolbox, available at https://github.com/anders-s-olsen/CopBET. CopBET provides a range of metrics that have been used in previous studies to quantify neural signal complexity. Our analysis focused on entropy metrics that have demonstrated the most robust neurobiological effects in prior psychedelic work ^24^ (See Figure 1).

### Lempel-Ziv complexity

We first applied the Hilbert transformation to the regional time series in order to obtain instantaneous amplitude envelopes. We then converted these to binary sequences where each timepoint above each region’s mean amplitude were assigned 1, and those below were assigned 0. This binary data forms a matrix of timepoints by regions. To calculate the Lempel-Ziv complexity (LZc), we concatenated the binary time series from consecutive spatial regions (2D to 1D) and applied the LZ76 compression algorithm. Then, we normalised the complexity score against LZc calculated on randomly permuted data to control for sequence length, which produces a single whole-brain spatial complexity score per scan session. Higher scores indicate more complexity, i.e. less compressible temporal patterns. This method provides a measure of how stable versus variable the brain’s configuration is over time.

### Meta-state complexity

We quantified brain state complexity by analysing temporal sequences of functional brain configurations across the entire brain. Regional time series from all subjects were concatenated and mean-centred. Common brain states were identified using K-means clustering (K = 4) with Pearson correlation distance, 200 random initialisations, and up to 1000 iterations per run. Consistent with previous studies using this method, the four clusters were reduced to two meta-states by pairing the most anti-correlated centroids ^21,50^. Each subject’s fMRI volumes were assigned to either meta-state 0 or 1, yielding individual binary sequences that represented brain-state transitions. The LZ76 compression algorithm was then used to calculate a single complexity score per subject, reflecting the incompressibility of brain-state switching ^49^.

### Modularity

The modularity of functional brain networks was quantified to assess the degree of network segregation into distinct communities. For each session, we applied the Louvain community detection algorithm as implemented in the *community_louvain* function from the Brain Connectivity Toolbox ^51^. This algorithm was applied to the interregional correlation matrices, which included both positive and negative correlations, using the ‘negative_asym’ flag to handle negative weights appropriately. To account for the stochasticity of the Louvain algorithm, we ran it 100 times for each correlation matrix from a scan session. Absolute modularity is the maximum modularity value (Q) across all iterations, and provides a measure of functional segregation in the brain. Higher values indicate stronger community structure, in which brain regions are more tightly connected within modules than between them. We also estimated normalised modularity to assess if the community structures exceeded chance expectations. To do this, we generated 100 null-model networks using the null_model_und_sign function in BCT, which preserves the weight and sign distributions but randomises the topology. For each null model, we repeated the community detection 100 times and saved the maximum Q. Normalised modularity was then calculated as the ratio of empirical modularity to the mean modularity of the null models. We report absolute modularity as our primary metric and normalised modularity in the supplementary materials (See supplementary Figure S8).

### Normalised spatial complexity

For normalised spatial complexity analysis (NSC) ^23^, voxel-wise time series are extracted and standardised to a mean of zero and a variance of one. Principal component analysis is then applied to the time-by-voxel matrix in order to identify principal components, which represent spatially distinct patterns of brain activity. The eigenvalues from the PCA are normalised by dividing each by the sum of probabilities, which results in a probability distribution that reflects the relative contribution of variance explained by each component. NSC is calculated using the Shannon entropy formula, NSC = -Σᵢ₌₁ᵐ λᵢ’ × log(λᵢ’) / ln(m), where λᵢ’ are the normalised eigenvalues and m is the number of components. This normalisation constrains NSC values between 0 and 1. Higher NSC values indicate that variance is distributed across many brain patterns, whereas lower values indicate that the spatial variance is concentrated in fewer patterns. Analyses were conducted both globally, using all brain voxels, and regionally, within individual brain networks.

### Multi-scale sample entropy

MSSE quantifies the regularity of temporal patterns by calculating the negative natural logarithm of the probability that recurring vectors of length m remain similar when extended to length m+1. We used a pattern length of m = 2 and a tolerance threshold of r = 0.3 times the standard deviation of each time series. To examine complexity across temporal scales, we applied multiscale analysis at two scales based on previous findings showing significant effects at these specific temporal resolutions ^24^. Scale 1 examines patterns at short time scales (3.6-5.4 seconds, as the TR was 1.8s), and scale 5 examines patterns at long time scales (18-27 seconds) by averaging consecutive non-overlapping groups of five volumes to capture slower neural dynamics. Sample entropy was calculated independently for each voxel; network-level values were derived by averaging across all voxels in each network. Higher entropy values represent more unpredictable and complex neural signals, whereas lower values indicate more regular activity patterns.

### Dynamical conditional correlation entropy

Dynamic conditional correlation (DCC) entropy captures the dynamic changes in functional connectivity between brain regions over time ^20^. We first extracted and mean-centred regional time series from the Schaefer 200-parcel atlas. For each session, the DCC algorithm estimated frame-wise connectivity by computing instantaneous correlations between all region pairs at each time point, resulting in a three-dimensional tensor describing the temporal evolution of connectivity. This was performed using a Generalised Autoregressive Conditional Heteroskedasticity model (GARCH).

To quantify the temporal variability of correlation values, we calculated Shannon entropy from all correlation values for each region pair. Specifically, we approximated the correlation distribution using MATLAB’s *histcounts* with probability normalisation. Higher DCC entropy indicates a broader distribution of connectivity values across the duration of the scan. Finally, to assess network-level effects, we averaged the DCC entropy within and between the seven brain networks. We summarised DCC entropy as the median across all region pairs within each network combination (e.g., motor–motor, motor–visual).

### Statistical analyses

Analyses were conducted in R version 4.4.0 (version 2024-04-24) using the *LMMstar* (version 1.1.0) package for repeated-measures linear mixed-effects models ^52^. Models included fixed effects for drug condition, sex, mean framewise displacement, previous drug exposure, number of visits, and study project. We specified subject-specific random intercepts with unstructured residual variance-covariance to account for within-subject correlation across drug sessions. The analysis framework was predefined and tested on simulated data in February 2025, before evaluating any statistical models on observed data (https://github.com/kristian1801/entropsi_planned_analysis_february_2025). The final analysis code can be found here (https://github.com/kristian1801/Entropy_final_analysis). Two notable analytical choices were changed towards a more conservative assessment of effects: 1) drug class effects are estimated weighting individual drug contributions evenly to limit bias due to imbalanced session frequency (e.g., many more LSD sessions vs. psilocybin or mescaline); 2) Holm-Bonferroni is applied across all 53 estimates, instead of only within related estimates (e.g., seven NSC networks).

#### Primary analysis

We hypothesised that psychedelics would differ from stimulants in their average effects on brain entropy measures. To test this, we evaluated the pooled effect of psychedelics vs. stimulants. The drug class (i.e., psychedelic/stimulant) effects were defined by pooling the drug-specific effects with equal weights (weight of ⅓ for psychedelics: LSD, psilocybin, mescaline; weight of ½ for stimulants: MDMA, d-Amphetamine). Effects were estimated using a linear mixed-effects model and evaluated with univariate Wald tests.

Our primary contrast of interest was psychedelics vs. stimulants, which we tested across 53 measures:

- **4 global measures:** Lempel–Ziv complexity, meta-state complexity, modularity, NSC
- **7 network-level NSC measures** (one per network)
- **14 multiscale sample entropy measures** (seven networks × two timescales)
- **28 dynamic conditional correlation measures** (seven within-network + 21 between-network pairs)

To control the family-wise error rate, we adjust p-values using the Holm-Bonferroni correction across all 53 measures ^53^.

#### Secondary analyses

Secondary analyses evaluated (i) psychedelics vs. placebo, (ii) stimulants vs. placebo, (iii) each drug vs. placebo, (iv) within-class drug comparisons (e.g., LSD vs. psilocybin), and (v) between-class comparisons (e.g., LSD vs. MDMA), a total of 17 additional contrasts. These contrasts were estimated using the same linear mixed-effects model and tested across the same 53 measures, with Holm–Bonferroni correction applied independently within each contrast, identical to the main analysis.

Results are reported as either the unstandardised regression coefficient or as a percentage change relative to placebo. Percentage change was calculated by dividing the model coefficient for each drug by the model intercept and expressing the result as a percentage.

## Code availability

The code used for the analyses is available on GitHub. The analysis framework was predefined and tested on simulated data in February 2025, before evaluating any statistical models on observed data (https://github.com/kristian1801/entropsi_planned_analysis_february_2025). The final analysis code can be found here (https://github.com/kristian1801/Entropy_final_analysis).

## Data availability

Data can be made available upon request to the corresponding author.

## Funding

The studies received funding from the Swiss National Science Foundation, specifically grant number 32003B_185111 for Matthias E. Liechti and grant number 320030_170249 for both Matthias E. Liechti and Stefan Borgwardt. Brice Ozenne states that part of his salary while working on this project was covered by a grant from Novo Nordisk A/S.

## Author contributions

P.M. Fisher, M. Avram, and K. Larsen conceptualised the study. M. Avram preprocessed the data. K. Larsen and B. Ozenne established the statistical framework. The analysis code was written by K. Larsen and reviewed by P.M. Fisher and B. Ozenne. K. Larsen analysed the data with feedback from B. Ozenne and P.M. Fisher. D.E. McCulloch and A.S. Olsen provided support for analyses and feedback on results. K. Larsen and P.M. Fisher wrote the original draft. F. Holze, L. Ley, P. Vizeli, A.M. Becker, F. Müller, S. Borgwardt, and M.E. Liechti acquired the data. All authors reviewed, edited, and approved the manuscript.

## Supplementary materials

### Supplementary Figures

- S1. Quality control summary
- S2. Drug effects on whole-brain entropy, complexity, and modularity
- S3. Drug effects on multiscale sample entropy across brain networks
- S4. Drug effects on normalised spatial complexity across brain networks
- S5. Drug effects on dynamic conditional correlation entropy across brain networks
- S6. Within drug class comparison of dynamic conditional correlation entropy change
- S7. Between-class comparisons of dynamic conditional correlation entropy
- S8. Absolute vs. normalised modularity under psychedelics and stimulants
- S9. Comparison of whole-brain metrics across studies

### Supplementary Tables

- S1. fMRI session counts across studies: Included and excluded sessions
- S2. Whole-brain entropy: Main effects (class-level contrasts)
- S3. Multiscale sample entropy (MSSE): Main effects (class-level contrasts)
- S4. Network-level NSC: Main effects (class-level contrasts)
- S5. DCC entropy: Main effects (class-level contrasts)
- S6. Meta-state complexity: All-drug comparisons (individual-drug within/between contrasts)
- S7. Modularity: All-drug comparisons (individual-drug within/between contrasts)
- S8. Lempel–Ziv complexity: All-drug comparisons (individual-drug within/between contrasts)
- S9. Whole-brain NSC: All-drug comparisons (individual-drug within/between contrasts)
- S10. Multiscale sample entropy (MSSE): All-drug comparisons (individual-drug within/between contrasts)
- S11. Network-level NSC: All-drug comparisons (individual-drug within/between contrasts)
- S12. DCC entropy: All-drug comparisons (individual-drug within/between contrasts)
- S13. QC: Regional coverage summary
- S14. QC: Network coverage summary
- S15. Consistency of placebo and LSD effects across studies

**Supplementary Figure S1.**
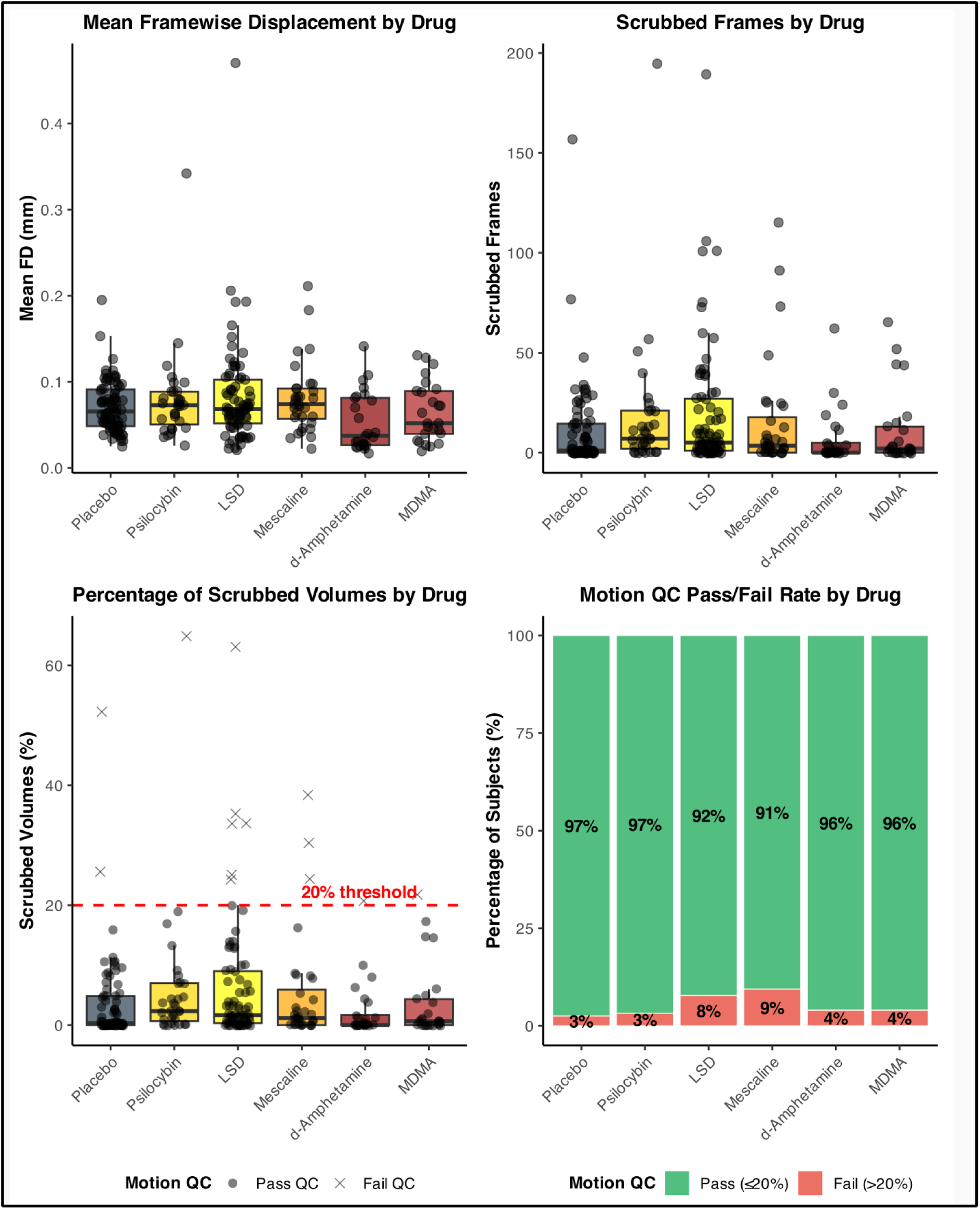
Quality control summary. (A) Mean framewise displacement (FD) across all drug conditions. (B) Number of scrubbed frames (FD > 0.2 mm). (C) Percentage of scrubbed frames 20% exclusion threshold (red dashed line). (D) Pass/fail rates: overall motion QC pass rates were high across all conditions, with slightly more exclusions in the LSD (8%) and mescaline (9%) conditions.

**Supplementary Figure S2.**
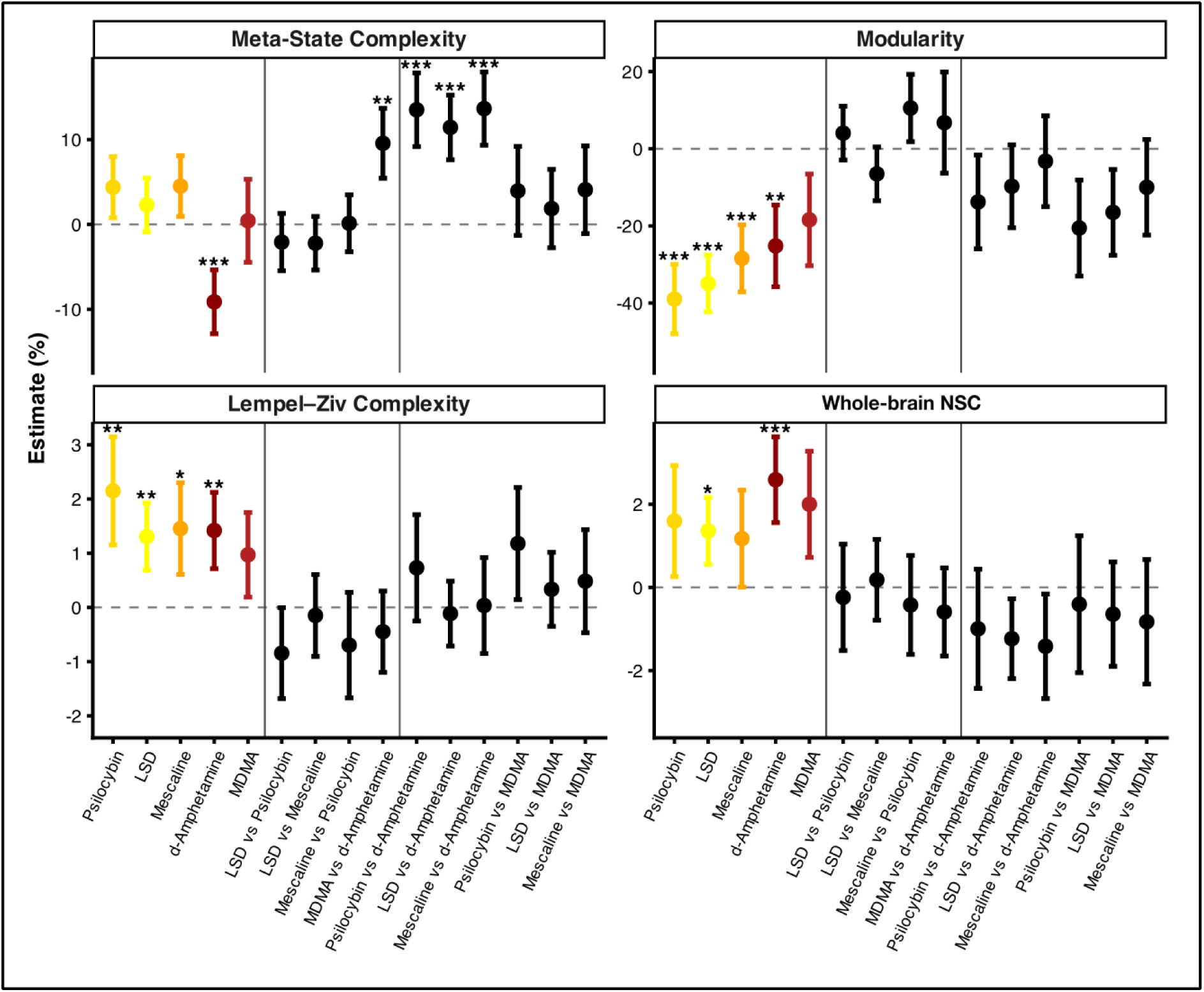
Drug effects on whole-brain entropy, complexity, and modularity. Forest plots showing model-based estimates of percentage change from placebo for whole-brain measures. Each panel is divided into sections displaying (i) individual drug effects relative to placebo (psychedelics in yellow: psilocybin, LSD, mescaline; stimulants in red: d-Amphetamine, MDMA), (ii) within-class pairwise comparisons, and (iii) between-class pairwise comparisons. Percentage changes were derived from model coefficients relative to the intercept (placebo), and pairwise contrasts were calculated directly from model coefficients. Points denote model estimates, and error bars indicate 95% confidence intervals. Asterisks indicate statistical significance after Holm-Bonferroni correction for 53 multiple comparisons (*p_FWER_ < 0.05, **p_FWER_ < 0.01, ***p_FWER_ < 0.001).

**Supplementary Figure S3.**
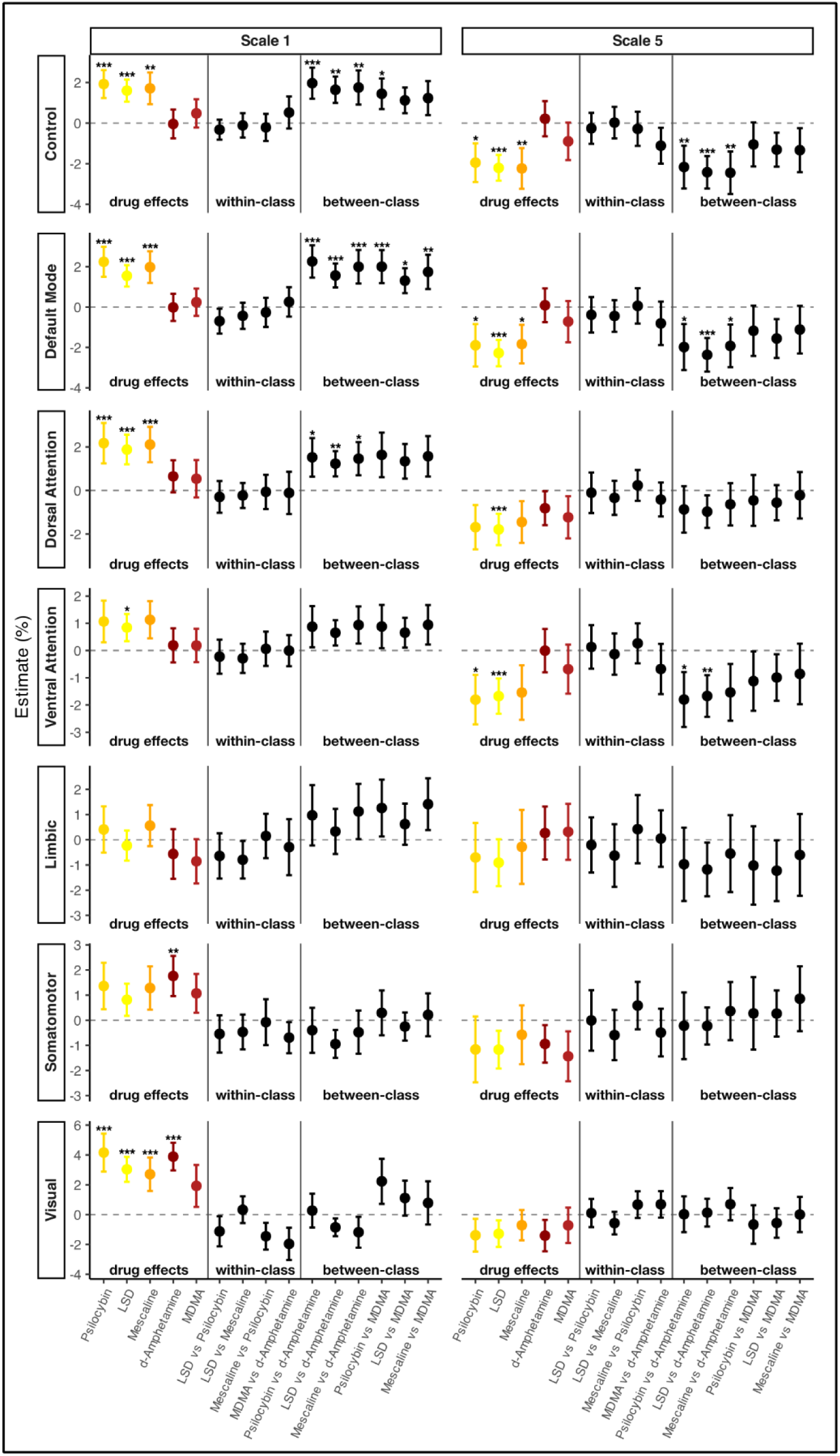
Drug effects on multiscale sample entropy across brain networks. Forest plots showing model-based estimates of percentage change from placebo for multiscale sample entropy at two temporal scales (Scale 1 and Scale 5) across the Yeo-7 functional brain networks (rows). Each panel is divided into sections displaying (i) individual drug effects relative to placebo (psychedelics in yellow: psilocybin, LSD, mescaline; stimulants in red: d-Amphetamine, MDMA), (ii) within-class pairwise comparisons, and (iii) between-class pairwise comparisons. Percentage changes were derived from model coefficients relative to the intercept (placebo), and pairwise contrasts were calculated directly from model coefficients. Points denote model estimates, and error bars indicate 95% confidence intervals. Asterisks indicate statistical significance after Holm-Bonferroni correction for 53 multiple comparisons (*p_FWER_ < 0.05, **p_FWER_ < 0.01, ***p_FWER_ < 0.001).

**Supplementary Figure S4.**
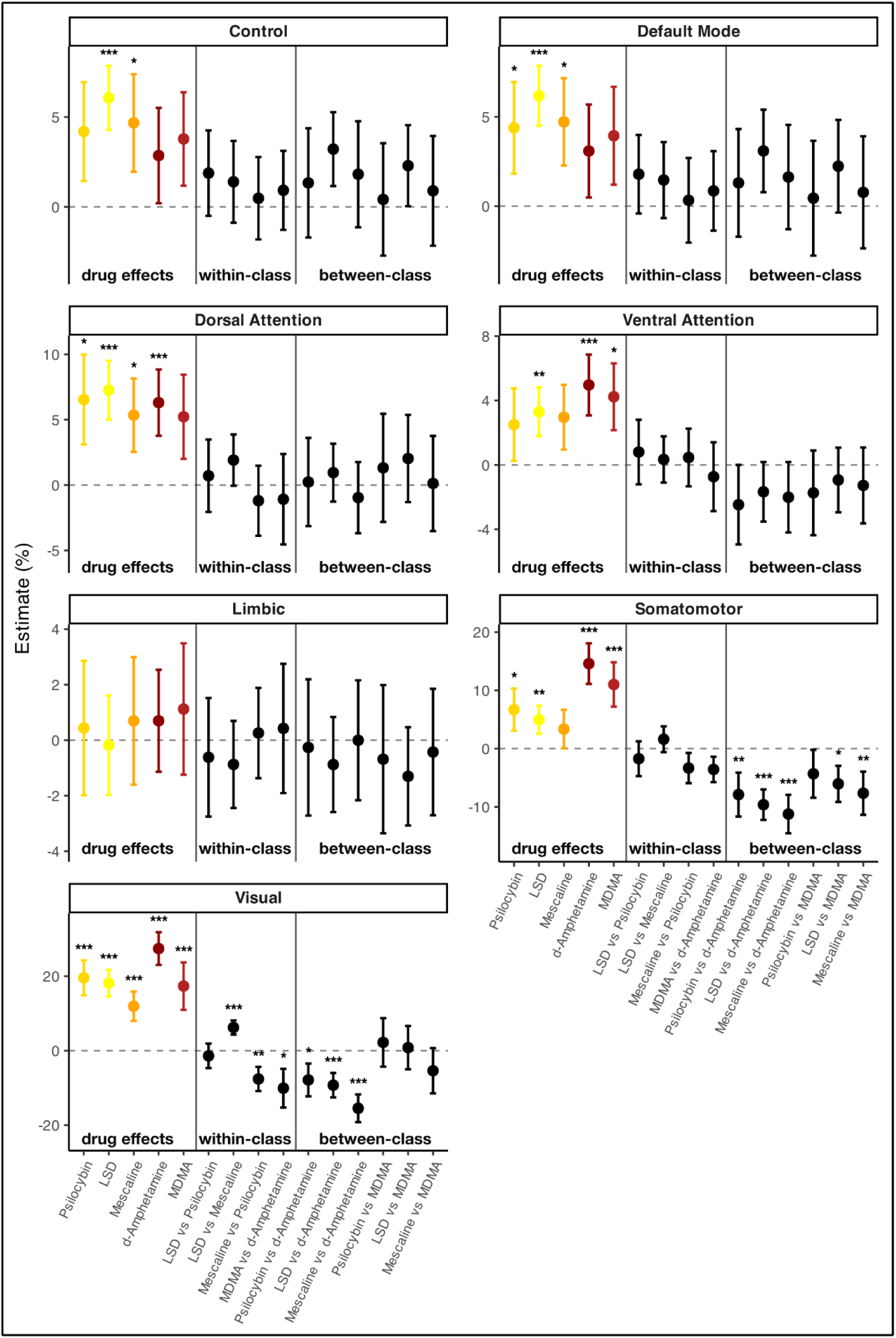
Drug effects on normalised spatial complexity across brain networks. Forest plots show model-based estimates of percentage change from placebo for normalised spatial complexity across the Yeo-7 functional brain networks (rows). Each panel is divided into sections displaying (i) individual drug effects relative to placebo (psychedelics in yellow: psilocybin, LSD, mescaline; stimulants in red: d-Amphetamine, MDMA), (ii) within-class pairwise comparisons, and (iii) between-class pairwise comparisons. Percentage changes were derived from model coefficients relative to the intercept (placebo), and pairwise contrasts were calculated directly from model coefficients. Points denote model estimates, and error bars indicate 95% confidence intervals. Asterisks indicate statistical significance after Holm-Bonferroni correction for 53 multiple comparisons (*p_FWER_ < 0.05, **p_FWER_ < 0.01, ***p_FWER_ < 0.001).

**Supplementary Figure S5.**
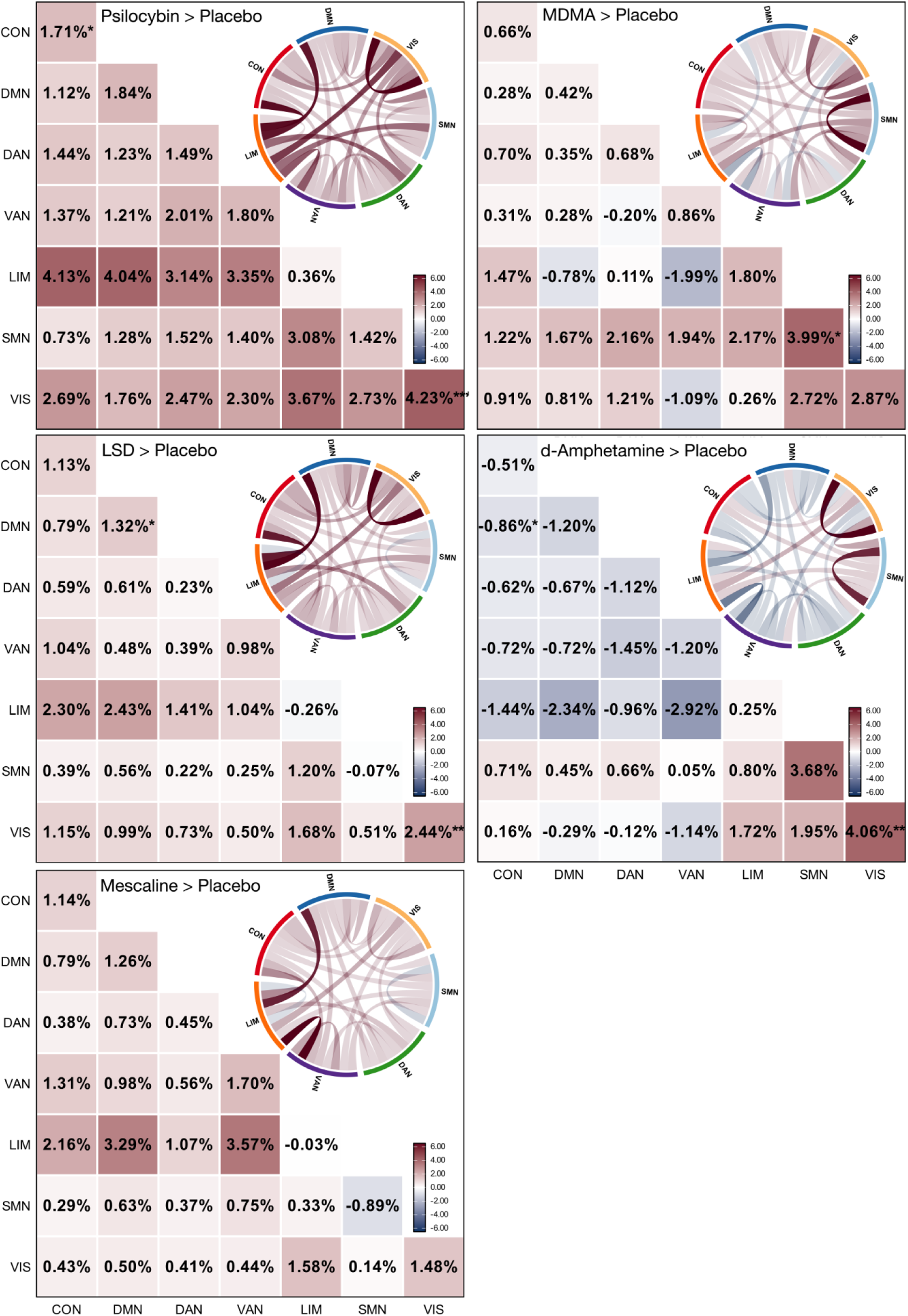
Drug effects on dynamic conditional correlation entropy across brain networks. Heat maps show changes in dynamic conditional correlation entropy relative to placebo, within- and between-networks, across the Yeo-7 seven functional brain networks. Each panel displays results for a single drug: psilocybin, LSD, mescaline, MDMA, and d-Amphetamine. Network abbreviations: CON = Control, DMN = Default Mode Network, DAN = Dorsal Attention Network, VAN = Ventral Attention Network, LIM = Limbic, SMN = Somatomotor Network, VIS = Visual. Colour scale ranges from -6.00% (blue, decreased DCC entropy) through 0% (white, no change) to +6.00% (red, increased DCC entropy). Asterisks denote statistically significant effects after Holm-Bonferroni correction for 53 multiple comparisons within each drug condition (*p_FWER_ < 0.05, **p_FWER_ < 0.01, ***p_FWER_ < 0.001). Chord diagram connections are colored by effect magnitude (red = positive, blue = negative).

**Supplementary Figure S6.**
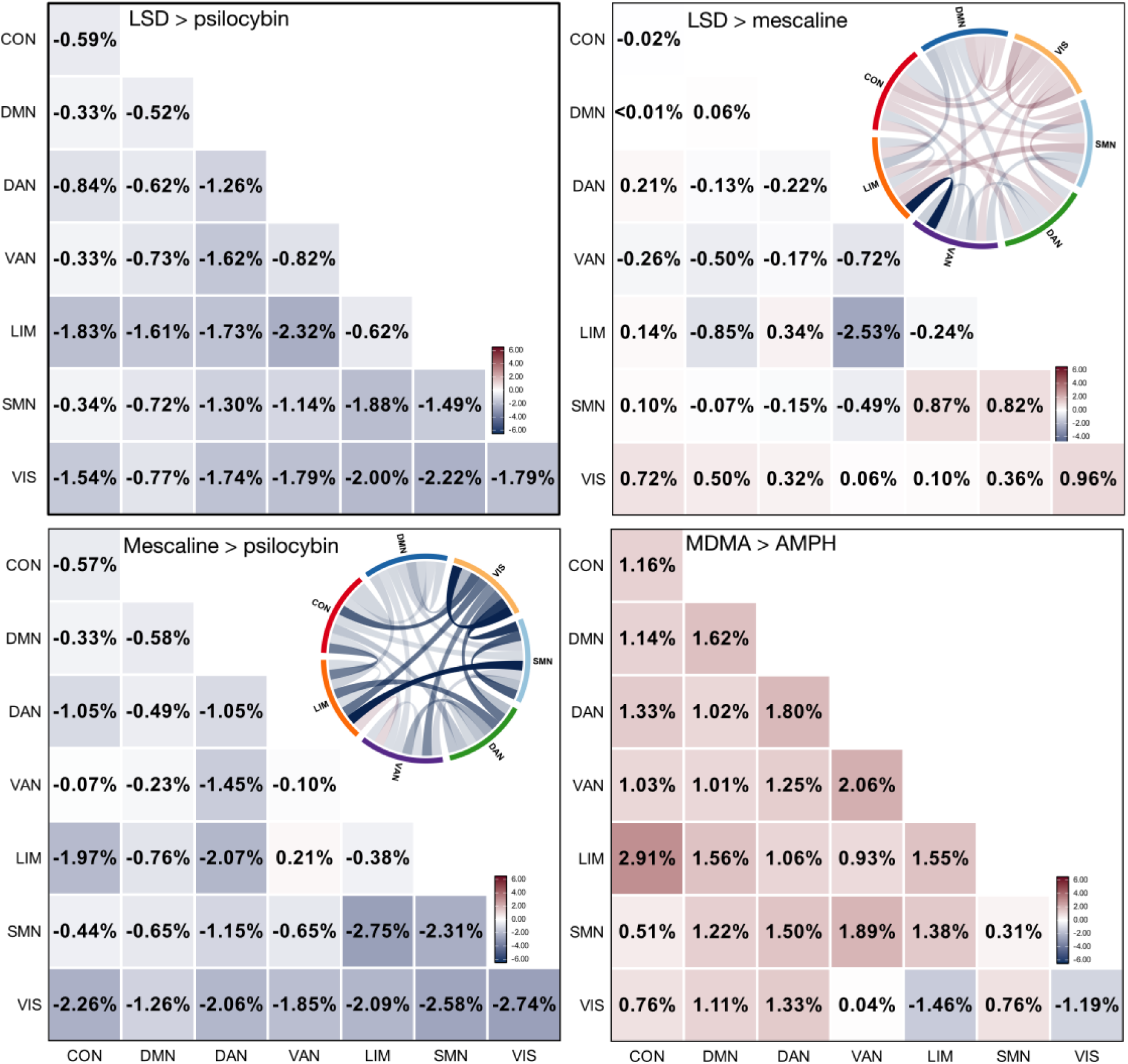
Within drug class comparison of dynamic conditional correlation entropy change. Network-to-network DCC entropy changes comparing classical psychedelics to other classical psychedelics (LSD vs. Psilocybin, LSD vs. Mescaline, Mescaline vs. Psilocybin) and stimulants to other stimulants (MDMA vs. d-Amphetamine). Each panel displays results for a single pairwise comparison. Network abbreviations: CON = Control, DMN = Default Mode Network, DAN = Dorsal Attention Network, VAN = Ventral Attention Network, LIM = Limbic, SMN = Somatomotor Network, VIS = Visual. Colour scale ranges from -6.00% (blue, decreased DCC entropy) through 0% (white, no change) to +6.00% (red, increased DCC entropy). Chord diagram connections are colored by effect magnitude (red = positive, blue = negative).

**Supplementary Figure S7.**
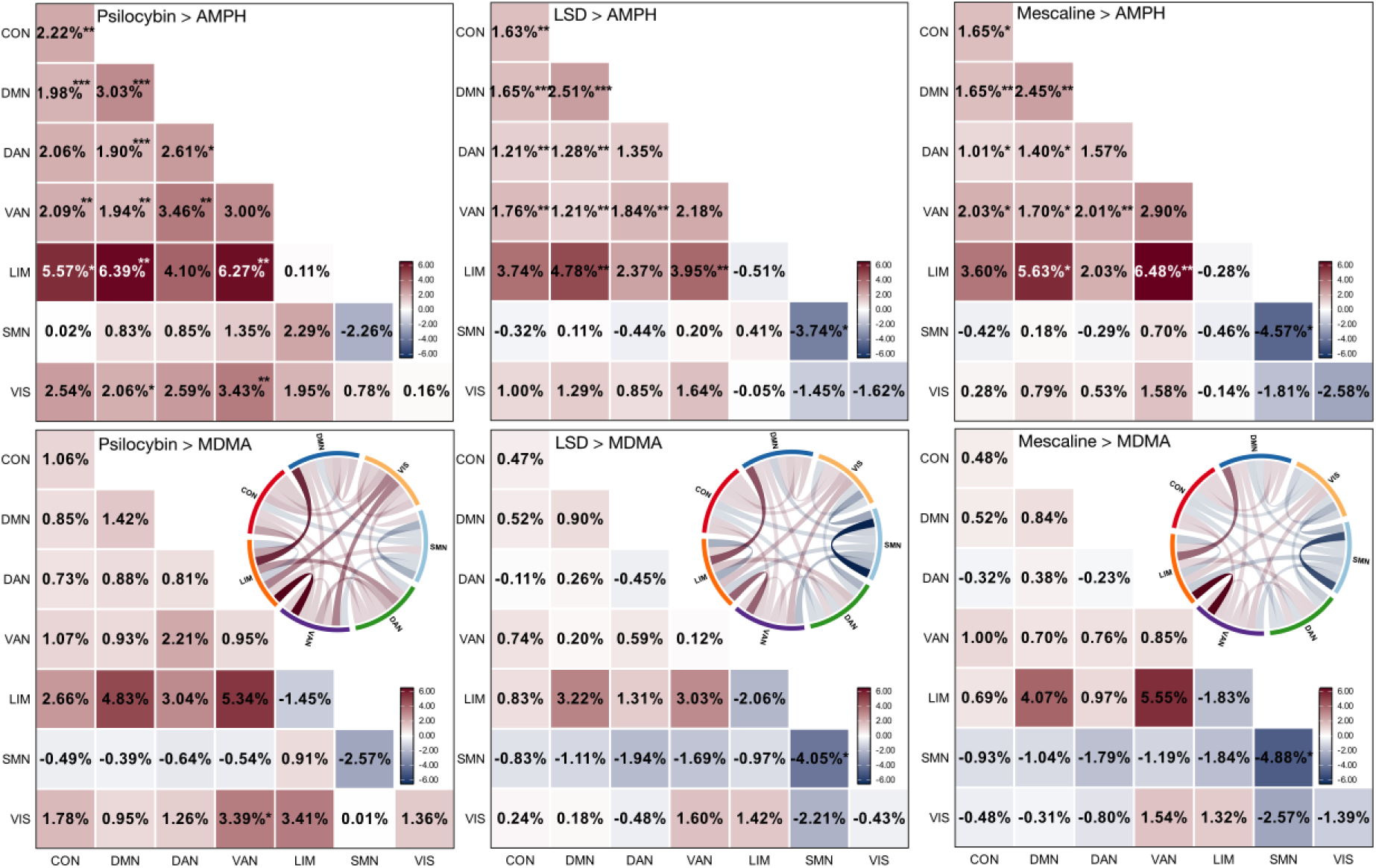
Between-class comparisons of dynamic conditional correlation entropy. Network-to-network DCC entropy changes between classical psychedelics (Psilocybin, LSD, Mescaline) and stimulants (d-Amphetamine, MDMA). Each panel displays results for a single pairwise comparison between drug classes. Network abbreviations: CON = Control, DMN = Default Mode Network, DAN = Dorsal Attention Network, VAN = Ventral Attention Network, LIM = Limbic, SMN = Somatomotor Network, VIS = Visual. Colour scale ranges from -6.00% (blue, decreased DCC entropy) through 0% (white, no change) to +6.00% (red, increased DCC entropy). Asterisks denote statistically significant effects after Holm-Bonferroni correction for 53 multiple comparisons within each pairwise comparison (*p_FWER_ < 0.05, **p_FWER_ < 0.01, ***p_FWER_ < 0.001). Chord diagram connections are colored by effect magnitude (red = positive, blue = negative).

**Supplementary Figure S8.**
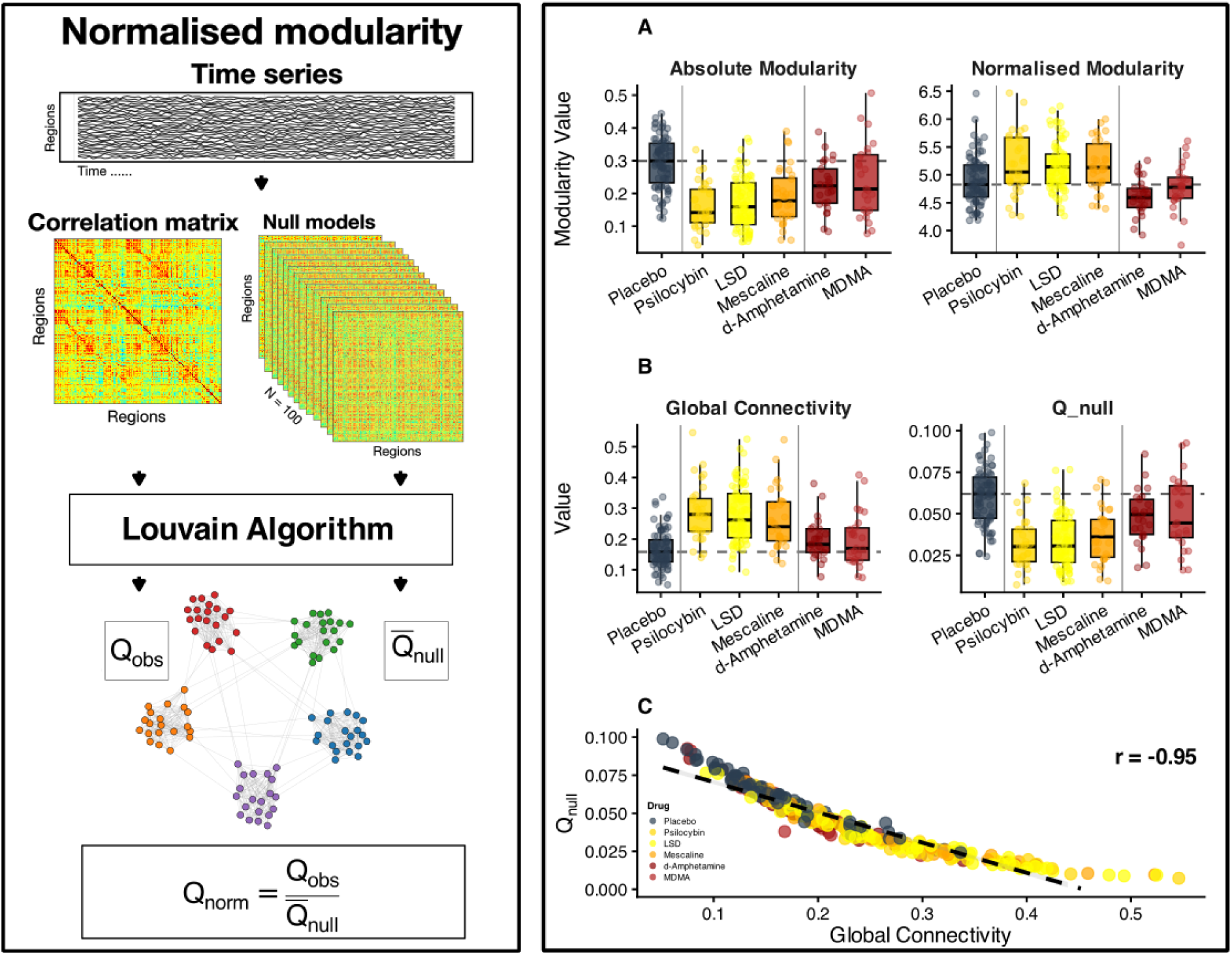
Absolute vs. normalised modularity under psychedelics and stimulants. **A.** Absolute modularity decreased both with psychedelics (yellow) and stimulants (red) compared to placebo (grey). By contrast, normalised modularity (Q_norm = Q_obs/Q_null) increased only for psychedelics. **B.** The changes in global connectivity (left: mean correlation across all region pairs) and the null model modularity scores (right: Q_null values). **C.** The correlation between global connectivity and Q_null. This strong negative correlation (r = -0.945) indicates that increases in global connectivity are strongly associated with lower null-model modularity; i.e., higher mean correlation produces a more uniform connectivity distribution. Importantly, when global connectivity increases, the null models approach random graphs, which will have modularity scores near zero. This explains why drugs that increase global connectivity, such as psychedelics, cause disproportionate reductions in Q_null, which leads to increased normalised modularity scores.

**Supplementary Figure 9.**
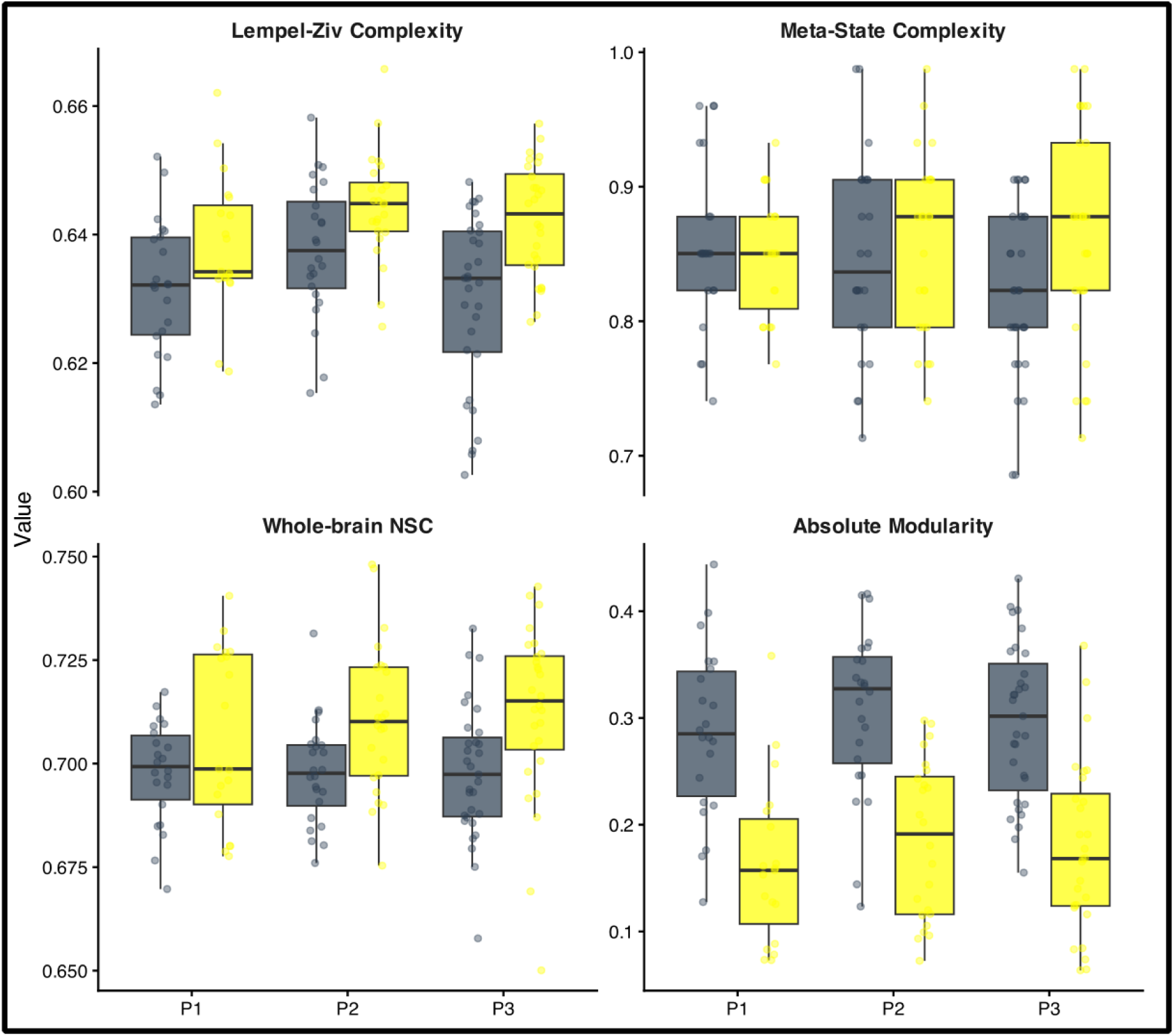
Comparison of whole-brain metrics across studies. Boxplots showing placebo (grey) and LSD (yellow) values for Lempel-Ziv complexity, meta-state complexity, whole-brain normalised signal complexity (NSC), and absolute modularity across the three independent studies (P1, P2, P3). Placebo values are similar across studies, and LSD shows directionally similar effects in all studies (increases in complexity measures, decreases in modularity), supporting the validity of pooling data.

## Notes

### Competing Interest Statement

The authors have declared no competing interest.

### Clinical Trial

Registration numbers for projects one, two and three: P1: NCT02308969, P2: NCT03019822, P3: NCT04227756

### Author Declarations

This research complies with all relevant ethical regulations and was approved by the Ethics Committee of Northwest Switzerland (EKNZ). Informed consent was obtained from all participants before enrollment. Our study draws on data from three clinical trials registered at ClinicalTrials.gov (registration numbers for projects one, two and three: P1: NCT02308969, P2: NCT03019822, P3: NCT04227756) combined N=79 unique participants with fMRI data. All MRI data were collected using a 3T Siemens scanner at the University Hospital, Basel, Switzerland, in accordance with the Declaration of Helsinki.

