## Supplemental Table 1 for "Psychedelics distinctly alter brain entropy and complexity compared to psychostimulants"

| Project | Sex f/m | Placebo | d-Amphetamine (40 mg) | MDMA (125 mg) | Psilocybin (20 mg) | LSD (100 µg) | Mescaline (300/500 mg) | Total |
|---|---|---|---|---|---|---|---|---|
| P1 | 11/11 | 22 (0) | 0 (0) | 0 (0) | 0 (0) | 19 (3) | 0 (0) | **41 (3)** |
| P2 | 12/13 | 24 (1) | 24 (1) | 24 (1) | 0 (0) | 24 (1) | 0 (0) | **96 (4)** |
| P3 | 16/16 | 31 (1) | 0 (0) | 0 (0) | 30 (1) | 28 (2) | 29 (3) | **118 (7)** |
| **Total** | **39/40** | **77 (2)** | **24 (1)** | **24 (1)** | **30 (1)** | **71 (6)** | **29 (3)** | **255 (14)** |

*Clinical Trial Registration: P1: NCT02308969; P2: NCT03019822; P3: NCT04227756, numbers under placebo and drug conditions indicate number of fMRI sessions and (n) excluded sessions*
