## Supplemental Table 2 for "Psychedelics distinctly alter brain entropy and complexity compared to psychostimulants"

| Contrast | Beta | Lower | Upper | p (unadjusted) | p (FWER) |
|---|---|---|---|---|---|
| **Psychedelics vs Stimulants** | | | | | |
| *Lempel-Ziv complexity* | 0.003 | -0.001 | 0.007 | 0.146 | 1.000 |
| *Global NSC* | -0.007 | -0.014 | 0.001 | 0.088 | 1.000 |
| ***Meta-state complexity*** | **0.068** | **0.037** | **0.098** | **<0.001** | **0.003** |
| *Absolute modularity* | -0.043 | -0.074 | -0.013 | 0.007 | 0.226 |
| **Stimulants vs placebo** | | | | | |
| ***Lempel-Ziv complexity*** | **0.008** | **0.004** | **0.012** | **<0.001** | **0.021** |
| ***Global NSC*** | **0.016** | **0.009** | **0.024** | **<0.001** | **0.004** |
| *Meta-state complexity* | -0.037 | -0.067 | -0.006 | 0.020 | 0.743 |
| ***Modularity*** | **-0.077** | **-0.109** | **-0.046** | **<0.001** | **<0.001** |
| **Psychedelics vs placebo** | | | | | |
| ***Lempel-Ziv complexity*** | **0.010** | **0.006** | **0.015** | **<0.001** | **<0.001** |
| *Global NSC* | 0.010 | 0.003 | 0.016 | 0.003 | 0.068 |
| *Meta-state complexity* | 0.031 | 0.008 | 0.055 | 0.008 | 0.160 |
| ***Absolute modularity*** | **-0.121** | **-0.149** | **-0.093** | **<0.001** | **<0.001** |

*Lower and Upper denote the lower and upper 95% confidence intervals, respectively; p (FWER) reflect Holm-Bonferroni adjusted p-values across 53 tests within a contrast (e.g., psychedelics vs. stimulants); NSC, Normalised spatial complexity*
