## Supplemental Table 3 for "Psychedelics distinctly alter brain entropy and complexity compared to psychostimulants"

| Multiscale sample entropy | | | | | | |
|---|---|---|---|---|---|---|
| **Contrast** | **Network** | **Beta** | **Lower** | **Upper** | **p** (unadjusted) | **p** (FWER) |
| **Psychedelics vs stimulants** | | | | | | |
| ***Scale 1*** | **Control** | **0.013** | **0.008** | **0.017** | **<0.001** | **<0.001** |
| | **Default mode** | **0.015** | **0.011** | **0.020** | **<0.001** | **<0.001** |
| | **Dorsal Attention** | **0.012** | **0.008** | **0.017** | **<0.001** | **<0.001** |
| | Ventral Attention | 0.007 | 0.003 | 0.011 | 0.002 | 0.072 |
| | Limbic | 0.008 | 0.002 | 0.015 | 0.012 | 0.363 |
| | Somatomotor | -0.002 | -0.007 | 0.003 | 0.357 | 1.000 |
| | Visual | 0.003 | -0.004 | 0.011 | 0.389 | 1.000 |
| ***Scale 5*** | **Control** | **-0.027** | **-0.038** | **-0.016** | **<0.001** | **<0.001** |
| | **Default mode** | **-0.025** | **-0.036** | **-0.014** | **<0.001** | **0.003** |
| | Dorsal Attention | -0.009 | -0.020 | 0.002 | 0.097 | 1.000 |
| | **Ventral Attention** | **-0.019** | **-0.030** | **-0.009** | **<0.001** | **0.019** |
| | Limbic | -0.013 | -0.029 | 0.002 | 0.082 | 1.000 |
| | Somatomotor | 0.003 | -0.008 | 0.015 | 0.578 | 1.000 |
| | Visual | -0.001 | -0.014 | 0.012 | 0.889 | 1.000 |
| **Stimulants vs placebo** | | | | | | |
| *Scale 1* | Control | 0.002 | -0.003 | 0.007 | 0.473 | 1.000 |
| | Default mode | 0.001 | -0.004 | 0.006 | 0.698 | 1.000 |
| | Dorsal Attention | 0.005 | 0.000 | 0.010 | 0.074 | 1.000 |
| | Ventral Attention | 0.002 | -0.003 | 0.007 | 0.520 | 1.000 |
| | Limbic | -0.006 | -0.012 | 0.000 | 0.053 | 1.000 |
| | **Somatomotor** | **0.012** | **0.006** | **0.018** | **<0.001** | **0.018** |
| | **Visual** | **0.024** | **0.015** | **0.032** | **<0.001** | **<0.001** |
| *Scale 5* | Control | -0.005 | -0.016 | 0.006 | 0.344 | 1.000 |
| | Default mode | -0.005 | -0.016 | 0.006 | 0.402 | 1.000 |
| | Dorsal Attention | -0.015 | -0.026 | -0.004 | 0.007 | 0.268 |
| | Ventral Attention | -0.005 | -0.015 | 0.005 | 0.312 | 1.000 |
| | Limbic | 0.004 | -0.008 | 0.016 | 0.481 | 1.000 |
| | Somatomotor | -0.017 | -0.028 | -0.007 | 0.002 | 0.064 |
| | Visual | -0.016 | -0.029 | -0.002 | 0.026 | 0.929 |
| **Psychedelics vs placebo** | | | | | | |
| ***Scale 1*** | **Control** | **0.015** | **0.010** | **0.019** | **<0.001** | **<0.001** |
| | **Default mode** | **0.016** | **0.011** | **0.021** | **<0.001** | **<0.001** |
| | **Dorsal Attention** | **0.017** | **0.011** | **0.023** | **<0.001** | **<0.001** |
| | **Ventral Attention** | **0.009** | **0.004** | **0.014** | **0.001** | **0.032** |
| | Limbic | 0.002 | -0.003 | 0.007 | 0.416 | 1.000 |
| | Somatomotor | 0.010 | 0.004 | 0.016 | 0.002 | 0.052 |
| | **Visual** | **0.027** | **0.019** | **0.035** | **<0.001** | **<0.001** |

| | | | | | | |
|---|---|---|---|---|---|---|
| ***Scale 5*** | **Control** | **-0.032** | **-0.042** | **-0.021** | **<0.001** | **<0.001** |
| | **Default mode** | **-0.030** | **-0.040** | **-0.020** | **<0.001** | **<0.001** |
| | **Dorsal Attention** | **-0.024** | **-0.035** | **-0.014** | **<0.001** | **<0.001** |
| | **Ventral Attentior** | **-0.024** | **-0.034** | **-0.015** | **<0.001** | **<0.001** |
| | Limbic | -0.009 | -0.023 | 0.005 | 0.201 | 1.000 |
| | Somatomotor | -0.014 | -0.025 | -0.003 | 0.010 | 0.178 |
| | Visual | -0.017 | -0.030 | -0.003 | 0.014 | 0.213 |

*Lower and Upper denote the lower and upper 95% confidence intervals, respectively; p (FWER) reflect Holm-Bonferroni adjusted p-values across 53 tests within a contrast (e.g., psychedelics vs. stimulants)*
