## Supplemental Table 4 for "Psychedelics distinctly alter brain entropy and complexity compared to psychostimulants"

| Network-level NSC | | | | | | |
| --- | --- | --- | --- | --- | --- | --- |
| **Contrast** | **Network** | **Beta** | **Lower** | **Upper** | **p** (unadjusted) | **p** (FWER) |
| **Psychedelics vs stimulants** | | | | | | |
| | Control | 0.008 | -0.003 | 0.020 | 0.146 | 1.000 |
| | Default mode | 0.008 | -0.004 | 0.020 | 0.185 | 1.000 |
| | Dorsal Attention | 0.003 | -0.009 | 0.015 | 0.599 | 1.000 |
| | Ventral Attention | -0.009 | -0.019 | 0.000 | 0.053 | 1.000 |
| | Limbic | -0.004 | -0.013 | 0.006 | 0.439 | 1.000 |
| | **Somatomotor** | **-0.041** | **-0.056** | **-0.027** | **<0.001** | **<0.001** |
| | Visual | -0.026 | -0.044 | -0.008 | 0.006 | 0.197 |
| **Stimulants vs placebo** | | | | | | |
| | Control | 0.017 | 0.005 | 0.029 | 0.007 | 0.280 |
| | Default mode | 0.018 | 0.006 | 0.030 | 0.005 | 0.225 |
| | **Dorsal Attentior** | **0.029** | **0.018** | **0.039** | **<0.001** | **<0.001** |
| | **Ventral Attentio** | **0.025** | **0.016** | **0.035** | **<0.001** | **<0.001** |
| | Limbic | 0.005 | -0.005 | 0.016 | 0.318 | 1.000 |
| | **Somatomotor** | **0.068** | **0.050** | **0.085** | **<0.001** | **<0.001** |
| | **Visual** | **0.101** | **0.080** | **0.121** | **<0.001** | **<0.001** |
| **Psychedelics vs placebo** | | | | | | |
| | **Control** | **0.025** | **0.015** | **0.036** | **<0.001** | **<0.001** |
| | **Default mode** | **0.026** | **0.016** | **0.036** | **<0.001** | **<0.001** |
| | **Dorsal Attentior** | **0.032** | **0.019** | **0.044** | **<0.001** | **<0.001** |
| | **Ventral Attentio** | **0.016** | **0.007** | **0.025** | **<0.001** | **0.023** |
| | Limbic | 0.002 | -0.009 | 0.013 | 0.730 | 1.000 |
| | **Somatomotor** | **0.026** | **0.011** | **0.042** | **<0.001** | **0.029** |
| | **Visual** | **0.074** | **0.057** | **0.092** | **<0.001** | **<0.001** |

*Lower and Upper denote the lower and upper 95% confidence intervals, respectively; p (FWER) reflect Holm-Bonferroni adjusted p-values across 53 tests within a contrast (e.g., psychedelics vs. stimulants); NSC, Normalised spatial compl*
