## Supplemental Table 5 for "Psychedelics distinctly alter brain entropy and complexity compared to psychostimulants"

### *Dynamic condition correlation entropy*

| Contrast | Network | Network | Beta | Lower | Upper | p (unadjusted) | p (FWER) |
|---|---|---|---|---|---|---|---|
| **Psychedelics vs stimulants** | | | | | | | |
| | Control | Control | 0.022 | 0.009 | 0.036 | 0.002 | 0.067 |
| | **Control** | **Default** | **0.021** | **0.010** | **0.033** | **<0.001** | **0.027** |
| | Control | Dorsal Attention | 0.014 | 0.001 | 0.026 | 0.034 | 1.000 |
| | Control | Limbic | 0.053 | -0.001 | 0.107 | 0.053 | 1.000 |
| | **Control** | **Ventral Attention** | **0.026** | **0.012** | **0.040** | **<0.001** | **0.021** |
| | Control | Somatomotor | -0.009 | -0.023 | 0.006 | 0.230 | 1.000 |
| | Control | Visual | 0.016 | -0.001 | 0.032 | 0.061 | 1.000 |
| | **Default** | **Default** | **0.033** | **0.015** | **0.051** | **<0.001** | **0.035** |
| | Default | Dorsal Attention | 0.018 | 0.004 | 0.032 | 0.010 | 0.315 |
| | **Default** | **Limbic** | **0.090** | **0.043** | **0.137** | **<0.001** | **0.015** |
| | Default | Ventral Attention | 0.020 | 0.006 | 0.033 | 0.005 | 0.187 |
| | Default | Somatomotor | -0.004 | -0.027 | 0.018 | 0.701 | 1.000 |
| | Default | Visual | 0.015 | -0.003 | 0.032 | 0.103 | 1.000 |
| | Dorsal Attention | Dorsal Attention | 0.017 | -0.006 | 0.040 | 0.147 | 1.000 |
| | Dorsal Attention | Limbic | 0.042 | -0.015 | 0.100 | 0.145 | 1.000 |
| | **Dorsal Attention** | **Ventral Attention** | **0.032** | **0.015** | **0.050** | **<0.001** | **0.021** |
| | Dorsal Attention | Somatomotor | -0.013 | -0.035 | 0.010 | 0.258 | 1.000 |
| | Dorsal Attention | Visual | 0.012 | -0.011 | 0.035 | 0.312 | 1.000 |
| | Limbic | Limbic | -0.019 | -0.093 | 0.055 | 0.602 | 1.000 |
| | **Limbic** | **Ventral Attention** | **0.094** | **0.051** | **0.137** | **<0.001** | **0.003** |
| | Limbic | Somatomotor | 0.001 | -0.064 | 0.066 | 0.975 | 1.000 |
| | Limbic | Visual | 0.024 | -0.025 | 0.073 | 0.328 | 1.000 |
| | Ventral Attention | Ventral Attention | 0.030 | 0.009 | 0.051 | 0.007 | 0.217 |
| | Ventral Attention | Somatomotor | -0.004 | -0.023 | 0.016 | 0.719 | 1.000 |
| | **Ventral Attention** | **Visual** | **0.040** | **0.017** | **0.062** | **<0.001** | **0.033** |
| | **Somatomotor** | **Somatomotor** | **-0.066** | **-0.098** | **-0.034** | **<0.001** | **0.008** |
| | Somatomotor | Visual | -0.022 | -0.053 | 0.010 | 0.174 | 1.000 |
| | Visual | Visual | -0.013 | -0.043 | 0.017 | 0.394 | 1.000 |
| **Stimulants vs placebo** | | | | | | | |
| | Control | Control | 0.001 | -0.011 | 0.013 | 0.820 | 1.000 |
| | Control | Default | -0.005 | -0.014 | 0.004 | 0.262 | 1.000 |
| | Control | Dorsal Attention | 0.001 | -0.010 | 0.011 | 0.894 | 1.000 |
| | Control | Limbic | 0.000 | -0.054 | 0.055 | 0.993 | 1.000 |
| | Control | Ventral Attention | -0.004 | -0.019 | 0.012 | 0.639 | 1.000 |
| | Control | Somatomotor | 0.017 | 0.002 | 0.033 | 0.030 | 1.000 |
| | Control | Visual | 0.009 | -0.003 | 0.022 | 0.134 | 1.000 |
| | Default | Default | -0.007 | -0.021 | 0.007 | 0.319 | 1.000 |
| | Default | Dorsal Attention | -0.003 | -0.014 | 0.008 | 0.603 | 1.000 |
| | Default | Limbic | -0.029 | -0.072 | 0.013 | 0.174 | 1.000 |
| | Default | Ventral Attention | -0.004 | -0.016 | 0.008 | 0.506 | 1.000 |
| | Default | Somatomotor | 0.019 | -0.002 | 0.040 | 0.072 | 1.000 |

| | | | | | | | |
|---|---|---|---|---|---|---|---|
| | Default | Visual | 0.005 | -0.011 | 0.020 | 0.553 | 1.000 |
| | Dorsal Attention | Dorsal Attention | -0.004 | -0.023 | 0.015 | 0.684 | 1.000 |
| | Dorsal Attention | Limbic | -0.008 | -0.058 | 0.042 | 0.756 | 1.000 |
| | Dorsal Attention | Ventral Attention | -0.015 | -0.032 | 0.003 | 0.100 | 1.000 |
| | Dorsal Attention | Somatomotor | 0.025 | 0.002 | 0.048 | 0.031 | 1.000 |
| | Dorsal Attention | Visual | 0.010 | -0.012 | 0.032 | 0.378 | 1.000 |
| | Limbic | Limbic | 0.019 | -0.064 | 0.103 | 0.640 | 1.000 |
| | Limbic | Ventral Attention | -0.045 | -0.086 | -0.005 | 0.028 | 0.990 |
| | Limbic | Somatomotor | 0.027 | -0.034 | 0.088 | 0.370 | 1.000 |
| | Limbic | Visual | 0.018 | -0.028 | 0.064 | 0.431 | 1.000 |
| | Ventral Attention | Ventral Attention | -0.003 | -0.021 | 0.015 | 0.731 | 1.000 |
| | Ventral Attention | Somatomotor | 0.018 | -0.001 | 0.037 | 0.067 | 1.000 |
| | Ventral Attention | Visual | -0.020 | -0.043 | 0.003 | 0.085 | 1.000 |
| | **Somatomotor** | **Somatomotor** | **0.068** | **0.037** | **0.100** | **<0.001** | **0.004** |
| | Somatomotor | Visual | 0.042 | 0.007 | 0.076 | 0.019 | 0.711 |
| | **Visual** | **Visual** | **0.060** | **0.029** | **0.091** | **<0.001** | **0.016** |
| **Psychedelics vs placebo** | | | | | | | |
| | **Control** | **Control** | **0.024** | **0.011** | **0.036** | **<0.001** | **0.008** |
| | **Control** | **Default** | **0.016** | **0.007** | **0.025** | **<0.001** | **0.012** |
| | **Control** | **Dorsal Attention** | **0.014** | **0.006** | **0.023** | **0.001** | **0.031** |
| | **Control** | **Limbic** | **0.053** | **0.022** | **0.084** | **0.001** | **0.032** |
| | Control | Ventral Attention | 0.022 | 0.008 | 0.036 | 0.002 | 0.050 |
| | Control | Somatomotor | 0.008 | 0.000 | 0.016 | 0.039 | 0.380 |
| | **Control** | **Visual** | **0.025** | **0.013** | **0.037** | **<0.001** | **0.005** |
| | **Default** | **Default** | **0.026** | **0.013** | **0.039** | **<0.001** | **0.008** |
| | Default | Dorsal Attention | 0.015 | 0.006 | 0.025 | 0.002 | 0.057 |
| | **Default** | **Limbic** | **0.061** | **0.032** | **0.090** | **<0.001** | **0.006** |
| | Default | Ventral Attention | 0.016 | 0.005 | 0.026 | 0.004 | 0.078 |
| | Default | Somatomotor | 0.015 | 0.002 | 0.028 | 0.026 | 0.310 |
| | **Default** | **Visual** | **0.019** | **0.009** | **0.030** | **<0.001** | **0.016** |
| | Dorsal Attention | Dorsal Attention | 0.013 | -0.003 | 0.029 | 0.109 | 0.766 |
| | Dorsal Attention | Limbic | 0.034 | 0.004 | 0.065 | 0.028 | 0.310 |
| | Dorsal Attention | Ventral Attention | 0.018 | 0.003 | 0.032 | 0.020 | 0.281 |
| | Dorsal Attention | Somatomotor | 0.013 | -0.001 | 0.026 | 0.072 | 0.578 |
| | **Dorsal Attention** | **Visual** | **0.021** | **0.007** | **0.035** | **0.004** | **0.078** |
| | Limbic | Limbic | 0.000 | -0.042 | 0.042 | 0.984 | 1.000 |
| | Limbic | Ventral Attention | 0.049 | 0.017 | 0.081 | 0.003 | 0.068 |
| | Limbic | Somatomotor | 0.028 | -0.014 | 0.071 | 0.187 | 1.000 |
| | Limbic | Visual | 0.042 | 0.014 | 0.071 | 0.004 | 0.082 |
| | Ventral Attention | Ventral Attention | 0.027 | 0.006 | 0.048 | 0.013 | 0.211 |
| | Ventral Attention | Somatomotor | 0.014 | 0.001 | 0.028 | 0.038 | 0.380 |
| | Ventral Attention | Visual | 0.019 | 0.005 | 0.034 | 0.009 | 0.160 |
| | Somatomotor | Somatomotor | 0.003 | -0.023 | 0.028 | 0.831 | 1.000 |
| | Somatomotor | Visual | 0.020 | 0.003 | 0.037 | 0.021 | 0.281 |
| | **Visual** | **Visual** | **0.047** | **0.026** | **0.067** | **<0.001** | **<0.001** |

*Lower and Upper denote the lower and upper 95% confidence intervals, respectively; p (FWER) reflect Holm-Bonferroni adjusted p-values across 53 tests within a contrast (e.g., psychedelics vs. stimulants)*
