## Supplemental Table 6 for "Psychedelics distinctly alter brain entropy and complexity compared to psychostimulants"

| Meta-State Complexity | | | | | |
|---|---|---|---|---|---|
| **Contrast** | **% Difference** | **Lower** | **Upper** | **p (unadjusted)** | **p (FWER)** |
| **Drug vs placebo** | | | | | |
| *LSD* | 2.303 | -0.857 | 5.463 | 0.151 | 1.000 |
| *Psilocybin* | 4.383 | 0.775 | 7.991 | 0.018 | 0.310 |
| *Mescaline* | 4.518 | 0.951 | 8.085 | 0.014 | 0.492 |
| *MDMA* | 0.434 | -4.456 | 5.323 | 0.858 | 1.000 |
| ***d-Amphetamine*** | **-9.125** | **-12.890** | **-5.361** | **<0.001** | **<0.001** |
| **Within-class comparison** | | | | | |
| *LSD vs Psilocybin* | -2.080 | -5.454 | 1.294 | 0.221 | 1.000 |
| *LSD vs Mescaline* | -2.215 | -5.374 | 0.944 | 0.164 | 1.000 |
| *Mescaline vs Psilocybin* | 0.135 | -3.217 | 3.486 | 0.935 | 1.000 |
| ***MDMA vs d-Amphetamine*** | **9.559** | **5.438** | **13.680** | **<0.001** | **0.005** |
| **Between-class comparison** | | | | | |
| ***Psilocybin vs d-Amphetamin*** | **13.508** | **9.166** | **17.850** | **<0.001** | **<0.001** |
| ***LSD vs d-Amphetamine*** | **11.428** | **7.619** | **15.238** | **<0.001** | **<0.001** |
| ***Mescaline vs d-Amphetamine*** | **13.643** | **9.337** | **17.949** | **<0.001** | **<0.001** |
| *Psilocybin vs MDMA* | 3.949 | -1.291 | 9.190 | 0.136 | 1.000 |
| *LSD vs MDMA* | 1.870 | -2.752 | 6.491 | 0.413 | 1.000 |
| *Mescaline vs MDMA* | 4.084 | -1.076 | 9.244 | 0.118 | 1.000 |

*Lower and Upper denote the lower and upper 95% confidence intervals, respectively; p (FWER) reflect Holm-Bonferroni adjusted p-values across 53 tests within a contrast (e.g., psychedelics vs. stimulants)*
