## Supplemental Table 7 for "Psychedelics distinctly alter brain entropy and complexity compared to psychostimulants"

| Absolute modularity | | | | | |
|---|---|---|---|---|---|
| **Contrast** | **% Difference** | **Lower** | **Upper** | **p (unadjusted)** | **p (FWER)** |
| **Drug vs placebo** | | | | | |
| ***LSD*** | **-34.915** | **-42.290** | **-27.540** | **0.007** | **<0.001** |
| ***Psilocybin*** | **-38.968** | **-47.976** | **-29.959** | **0.029** | **<0.001** |
| ***Mescaline*** | **-28.412** | **-37.092** | **-19.732** | **0.019** | **<0.001** |
| ***MDMA*** | **-18.422** | **-30.311** | **-6.534** | **0.100** | **0.003** |
| ***d-Amphetamine*** | **-25.189** | **-35.780** | **-14.598** | **<0.001** | **<0.001** |
| **Within-class comparison** | | | | | |
| *LSD vs Psilocybin* | 4.053 | -2.907 | 11.013 | 0.583 | 0.242 |
| *LSD vs Mescaline* | -6.503 | -13.477 | 0.471 | 0.515 | 0.066 |
| *Mescaline vs Psilocybin* | 10.556 | 1.839 | 19.272 | 0.968 | 0.019 |
| *MDMA vs d-Amphetamine* | 6.767 | -6.344 | 19.878 | 0.234 | 0.297 |
| **Between-class comparison** | | | | | |
| *Psilocybin vs d-Amphetamine* | -13.778 | -25.921 | -1.636 | <0.001 | 0.027 |
| *LSD vs d-Amphetamine* | -9.726 | -20.483 | 1.031 | <0.001 | 0.074 |
| *Mescaline vs d-Amphetamine* | -3.223 | -14.989 | 8.544 | <0.001 | 0.583 |
| ***Psilocybin vs MDMA*** | **-20.545** | **-32.995** | **-8.095** | **0.003** | **0.002** |
| ***LSD vs MDMA*** | **-16.493** | **-27.627** | **-5.359** | **0.002** | **0.005** |
| *Mescaline vs MDMA* | -9.990 | -22.388 | 2.408 | 0.002 | 0.111 |

*Lower and Upper denote the lower and upper 95% confidence intervals, respectively; p (FWER) reflect Holm-Bonferroni adjusted p-values across 53 tests within a contrast (e.g., psychedelics vs. stimulants)*
