## Supplemental Table 8 for "Psychedelics distinctly alter brain entropy and complexity compared to psychostimulants"

| *Lempel-Ziv complexity* | | | | | |
|---|---|---|---|---|---|
| **Contrast** | **% Difference** | **Lower** | **Upper** | **p (unadjusted)** | **p (FWER)** |
| **Drug vs placebo** | | | | | |
| ***LSD*** | **1.306** | **0.687** | **1.925** | **<0.001** | **0.002** |
| ***Psilocybin*** | **2.149** | **1.153** | **3.146** | **<0.001** | **0.003** |
| ***Mescaline*** | **1.454** | **0.609** | **2.300** | **0.001** | **0.048** |
| *MDMA* | 0.971 | 0.190 | 1.751 | 0.016 | 0.633 |
| ***d-Amphetamine*** | **1.419** | **0.715** | **2.123** | **<0.001** | **0.008** |
| **Within-class comparison** | | | | | |
| *LSD vs Psilocybin* | -0.844 | -1.682 | -0.005 | 0.049 | 1.000 |
| *LSD vs Mescaline* | -0.149 | -0.905 | 0.607 | 0.691 | 1.000 |
| *Mescaline vs Psilocybin* | -0.695 | -1.668 | 0.278 | 0.154 | 1.000 |
| *MDMA vs d-Amphetamine* | -0.448 | -1.198 | 0.302 | 0.227 | 1.000 |
| **Between-class comparison** | | | | | |
| *Psilocybin vs d-Amphetamine* | 0.731 | -0.250 | 1.711 | 0.140 | 1.000 |
| *LSD vs d-Amphetamine* | -0.113 | -0.711 | 0.485 | 0.701 | 1.000 |
| *Mescaline vs d-Amphetamine* | 0.036 | -0.849 | 0.920 | 0.936 | 1.000 |
| *Psilocybin vs MDMA* | 1.179 | 0.146 | 2.212 | 0.026 | 1.000 |
| *LSD vs MDMA* | 0.335 | -0.348 | 1.018 | 0.320 | 1.000 |
| *Mescaline vs MDMA* | 0.484 | -0.467 | 1.435 | 0.311 | 1.000 |

*Lower and Upper denote the lower and upper 95% confidence intervals, respectively; p (FWER) reflect Holm-Bonferroni adjusted p-values across 53 tests within a contrast (e.g., psychedelics vs. stimulants)*
