## Supplemental Table 9 for "Psychedelics distinctly alter brain entropy and complexity compared to psychostimulants"

| Whole-brain NSC | | | | | |
|---|---|---|---|---|---|
| **Contrast** | **% Difference** | **Lower** | **Upper** | **p (unadjusted)** | **p (FWER)** |
| **Drug vs placebo** | | | | | |
| ***LSD*** | **1.358** | **0.555** | **2.161** | **0.001** | **0.037** |
| *Psilocybin* | 1.596 | 0.261 | 2.932 | 0.020 | 0.323 |
| *Mescaline* | 1.174 | 0.008 | 2.341 | 0.049 | 1.000 |
| *MDMA* | 2.000 | 0.723 | 3.278 | 0.003 | 0.158 |
| ***d-Amphetamine*** | **2.590** | **1.560** | **3.621** | **<0.001** | **<0.001** |
| **Within-class comparison** | | | | | |
| *LSD vs Psilocybin* | -0.238 | -1.516 | 1.039 | 0.708 | 1.000 |
| *LSD vs Mescaline* | 0.184 | -0.788 | 1.156 | 0.702 | 1.000 |
| *Mescaline vs Psilocybin* | -0.422 | -1.611 | 0.767 | 0.472 | 1.000 |
| *MDMA vs d-Amphetamine* | -0.590 | -1.649 | 0.470 | 0.261 | 1.000 |
| **Between-class comparison** | | | | | |
| *Psilocybin vs d-Amphetamine* | -0.994 | -2.427 | 0.439 | 0.170 | 1.000 |
| *LSD vs d-Amphetamine* | -1.232 | -2.192 | -0.272 | 0.014 | 0.304 |
| *Mescaline vs d-Amphetamine* | -1.416 | -2.674 | -0.157 | 0.028 | 0.734 |
| *Psilocybin vs MDMA* | -0.404 | -2.050 | 1.241 | 0.624 | 1.000 |
| *LSD vs MDMA* | -0.642 | -1.898 | 0.613 | 0.301 | 1.000 |
| *Mescaline vs MDMA* | -0.826 | -2.323 | 0.671 | 0.272 | 1.000 |

*Lower and Upper denote the lower and upper 95% confidence intervals, respectively; p (FWER) reflect Holm-Bonferroni adjusted p-values across 53 tests within a contrast (e.g., psychedelics vs. stimulants); NSC, Normalised spatial complexity*
