## Supplemental Table 10 for "Psychedelics distinctly alter brain entropy and complexity compared to psychostimulants"

| *Multiscale sample entropy* | | | | | | |
|---|---|---|---|---|---|---|
| **Contrast** | **Network** | **% Difference** | **Lower** | **Upper** | **p (unadjusted)** | **p (FWER)** |
| **Drug vs placebo** | | | | | | |
| ***LSD*** | | | | | | |
| ***Scale 1*** | **Control** | **1.600** | **1.059** | **2.141** | **<0.001** | **<0.001** |
| | **Default mode** | **1.548** | **1.019** | **2.076** | **<0.001** | **<0.001** |
| | **Dorsal Attention** | **1.878** | **1.200** | **2.556** | **<0.001** | **<0.001** |
| | **Ventral Attention** | **0.843** | **0.346** | **1.340** | **0.001** | **0.037** |
| | Limbic | -0.230 | -0.827 | 0.367 | 0.447 | 1.000 |
| | Somatomotor | 0.815 | 0.175 | 1.455 | 0.013 | 0.340 |
| | **Visual** | **3.036** | **2.197** | **3.874** | **<0.001** | **<0.001** |
| ***Scale 5*** | **Control** | **-2.204** | **-2.839** | **-1.570** | **<0.001** | **<0.001** |
| | **Default mode** | **-2.278** | **-2.931** | **-1.625** | **<0.001** | **<0.001** |
| | **Dorsal Attention** | **-1.787** | **-2.507** | **-1.068** | **<0.001** | **<0.001** |
| | **Ventral Attention** | **-1.675** | **-2.323** | **-1.027** | **<0.001** | **<0.001** |
| | Limbic | -0.907 | -1.836 | 0.022 | 0.056 | 1.000 |
| | Somatomotor | -1.169 | -1.920 | -0.418 | 0.003 | 0.086 |
| | Visual | -1.279 | -2.171 | -0.387 | 0.005 | 0.155 |
| ***Psilocybin*** | | | | | | |
| ***Scale 1*** | **Control** | **1.923** | **1.236** | **2.611** | **<0.001** | **<0.001** |
| | **Default mode** | **2.245** | **1.499** | **2.990** | **<0.001** | **<0.001** |
| | **Dorsal Attention** | **2.172** | **1.244** | **3.099** | **<0.001** | **<0.001** |
| | Ventral Attention | 1.067 | 0.301 | 1.833 | 0.007 | 0.159 |
| | Limbic | 0.410 | -0.508 | 1.328 | 0.374 | 1.000 |
| | Somatomotor | 1.361 | 0.437 | 2.284 | 0.005 | 0.128 |
| | **Visual** | **4.159** | **2.889** | **5.430** | **<0.001** | **<0.001** |
| ***Scale 5*** | **Control** | **-1.947** | **-2.900** | **-0.995** | **<0.001** | **0.010** |
| | **Default mode** | **-1.894** | **-2.943** | **-0.846** | **<0.001** | **0.035** |
| | Dorsal Attention | -1.685 | -2.704 | -0.666 | 0.002 | 0.072 |
| | **Ventral Attention** | **-1.806** | **-2.714** | **-0.899** | **<0.001** | **0.010** |
| | Limbic | -0.702 | -2.073 | 0.669 | 0.310 | 1.000 |
| | Somatomotor | -1.162 | -2.474 | 0.150 | 0.081 | 0.568 |
| | Visual | -1.382 | -2.478 | -0.286 | 0.015 | 0.296 |
| ***Mescaline*** | | | | | | |
| ***Scale 1*** | **Control** | **1.712** | **0.933** | **2.491** | **<0.001** | **0.002** |
| | **Default mode** | **1.981** | **1.196** | **2.766** | **<0.001** | **<0.001** |
| | **Dorsal Attention** | **2.108** | **1.293** | **2.923** | **<0.001** | **<0.001** |
| | Ventral Attention | 1.130 | 0.447 | 1.812 | 0.002 | 0.064 |
| | Limbic | 0.562 | -0.254 | 1.378 | 0.173 | 1.000 |
| | Somatomotor | 1.283 | 0.423 | 2.143 | 0.004 | 0.163 |
| | **Visual** | **2.708** | **1.588** | **3.828** | **<0.001** | **<0.001** |

| | | | | | | |
|---|---|---|---|---|---|---|
| ***Scale 5*** | **Control** | **-2.231** | **-3.231** | **-1.231** | **<0.001** | **0.005** |
| | **Default mode** | **-1.835** | **-2.792** | **-0.879** | **<0.001** | **0.020** |
| | Dorsal Attention | -1.449 | -2.407 | -0.491 | 0.004 | 0.165 |
| | Ventral Attention | -1.541 | -2.544 | -0.539 | 0.003 | 0.130 |
| | Limbic | -0.280 | -1.749 | 1.189 | 0.702 | 1.000 |
| | Somatomotor | -0.578 | -1.749 | 0.593 | 0.324 | 1.000 |
| | Visual | -0.710 | -1.726 | 0.305 | 0.166 | 1.000 |
| ***MDMA*** | | | | | | |
| *Scale 1* | Control | 0.480 | -0.216 | 1.176 | 0.171 | 1.000 |
| | Default mode | 0.240 | -0.437 | 0.918 | 0.477 | 1.000 |
| | Dorsal Attention | 0.538 | -0.320 | 1.395 | 0.211 | 1.000 |
| | Ventral Attention | 0.186 | -0.428 | 0.799 | 0.544 | 1.000 |
| | Limbic | -0.853 | -1.730 | 0.025 | 0.056 | 1.000 |
| | Somatomotor | 1.068 | 0.296 | 1.841 | 0.008 | 0.342 |
| | Visual | 1.926 | 0.523 | 3.329 | 0.009 | 0.363 |
| *Scale 5* | Control | -0.897 | -1.819 | 0.024 | 0.056 | 1.000 |
| | Default mode | -0.719 | -1.742 | 0.303 | 0.162 | 1.000 |
| | Dorsal Attention | -1.230 | -2.202 | -0.257 | 0.014 | 0.574 |
| | Ventral Attention | -0.683 | -1.583 | 0.216 | 0.132 | 1.000 |
| | Limbic | 0.319 | -0.793 | 1.430 | 0.564 | 1.000 |
| | Somatomotor | -1.434 | -2.429 | -0.440 | 0.006 | 0.268 |
| | Visual | -0.717 | -1.903 | 0.469 | 0.229 | 1.000 |
| ***d-Amphetamine*** | | | | | | |
| *Scale 1* | Control | -0.042 | -0.746 | 0.663 | 0.906 | 1.000 |
| | Default mode | -0.016 | -0.687 | 0.654 | 0.961 | 1.000 |
| | Dorsal Attention | 0.649 | -0.087 | 1.385 | 0.083 | 1.000 |
| | Ventral Attention | 0.191 | -0.435 | 0.818 | 0.544 | 1.000 |
| | Limbic | -0.562 | -1.549 | 0.426 | 0.255 | 1.000 |
| | **Somatomotor** | **1.761** | **0.963** | **2.558** | **<0.001** | **0.002** |
| | **Visual** | **3.887** | **2.964** | **4.811** | **<0.001** | **<0.001** |
| *Scale 5* | Control | 0.215 | -0.650 | 1.080 | 0.619 | 1.000 |
| | Default mode | 0.089 | -0.752 | 0.929 | 0.832 | 1.000 |
| | Dorsal Attention | -0.815 | -1.593 | -0.037 | 0.041 | 1.000 |
| | Ventral Attention | -0.005 | -0.801 | 0.791 | 0.990 | 1.000 |
| | Limbic | 0.268 | -0.783 | 1.320 | 0.609 | 1.000 |
| | Somatomotor | -0.943 | -1.692 | -0.193 | 0.015 | 0.491 |
| | Visual | -1.409 | -2.464 | -0.354 | 0.010 | 0.368 |
| **Within-class comparison** | | | | | | |
| ***LSD vs Psilocybin*** | | | | | | |
| *Scale 1* | Control | -0.324 | -0.816 | 0.169 | 0.190 | 1.000 |
| | Default mode | -0.697 | -1.312 | -0.082 | 0.028 | 1.000 |
| | Dorsal Attention | -0.294 | -1.025 | 0.437 | 0.417 | 1.000 |
| | Ventral Attention | -0.224 | -0.850 | 0.402 | 0.470 | 1.000 |

| | | | | | | |
|---|---|---|---|---|---|---|
| | Limbic | -0.640 | -1.539 | 0.259 | 0.158 | 1.000 |
| | Somatomotor | -0.546 | -1.285 | 0.194 | 0.143 | 1.000 |
| | Visual | -1.124 | -2.131 | -0.116 | 0.030 | 1.000 |
| *Scale 5* | Control | -0.257 | -1.021 | 0.508 | 0.493 | 1.000 |
| | Default mode | -0.384 | -1.265 | 0.498 | 0.373 | 1.000 |
| | Dorsal Attention | -0.103 | -1.035 | 0.829 | 0.823 | 1.000 |
| | Ventral Attention | 0.131 | -0.668 | 0.930 | 0.741 | 1.000 |
| | Limbic | -0.205 | -1.298 | 0.888 | 0.706 | 1.000 |
| | Somatomotor | -0.007 | -1.210 | 1.195 | 0.990 | 1.000 |
| | Visual | 0.103 | -0.842 | 1.047 | 0.826 | 1.000 |
| ***LSD vs Mescaline*** | | | | | | |
| *Scale 1* | Control | -0.112 | -0.716 | 0.492 | 0.709 | 1.000 |
| | Default mode | -0.433 | -1.081 | 0.215 | 0.183 | 1.000 |
| | Dorsal Attention | -0.230 | -0.808 | 0.348 | 0.423 | 1.000 |
| | Ventral Attention | -0.287 | -0.824 | 0.250 | 0.284 | 1.000 |
| | Limbic | -0.792 | -1.538 | -0.046 | 0.038 | 1.000 |
| | Somatomotor | -0.468 | -1.161 | 0.226 | 0.178 | 1.000 |
| | Visual | 0.327 | -0.571 | 1.226 | 0.464 | 1.000 |
| *Scale 5* | Control | 0.026 | -0.749 | 0.801 | 0.943 | 1.000 |
| | Default mode | -0.443 | -1.223 | 0.338 | 0.251 | 1.000 |
| | Dorsal Attention | -0.339 | -1.124 | 0.447 | 0.382 | 1.000 |
| | Ventral Attention | -0.134 | -0.892 | 0.624 | 0.720 | 1.000 |
| | Limbic | -0.627 | -1.869 | 0.615 | 0.311 | 1.000 |
| | Somatomotor | -0.591 | -1.593 | 0.411 | 0.236 | 1.000 |
| | Visual | -0.569 | -1.323 | 0.185 | 0.134 | 1.000 |
| ***Mescaline vs Psilocybin*** | | | | | | |
| *Scale 1* | Control | -0.211 | -0.876 | 0.453 | 0.519 | 1.000 |
| | Default mode | -0.264 | -0.983 | 0.456 | 0.457 | 1.000 |
| | Dorsal Attention | -0.063 | -0.849 | 0.722 | 0.868 | 1.000 |
| | Ventral Attention | 0.063 | -0.566 | 0.692 | 0.838 | 1.000 |
| | Limbic | 0.152 | -0.727 | 1.031 | 0.726 | 1.000 |
| | Somatomotor | -0.078 | -0.991 | 0.836 | 0.862 | 1.000 |
| | Visual | -1.451 | -2.345 | -0.557 | 0.003 | 0.135 |
| *Scale 5* | Control | -0.283 | -1.122 | 0.556 | 0.490 | 1.000 |
| | Default mode | 0.059 | -0.821 | 0.939 | 0.891 | 1.000 |
| | Dorsal Attention | 0.236 | -0.472 | 0.944 | 0.499 | 1.000 |
| | Ventral Attention | 0.265 | -0.468 | 0.997 | 0.467 | 1.000 |
| | Limbic | 0.422 | -0.934 | 1.777 | 0.527 | 1.000 |
| | Somatomotor | 0.584 | -0.360 | 1.528 | 0.215 | 1.000 |
| | Visual | 0.672 | -0.225 | 1.569 | 0.135 | 1.000 |
| *MDMA vs d-Amphetamine* | | | | | | |
| *Scale 1* | Control | 0.521 | -0.266 | 1.309 | 0.184 | 1.000 |

| | | | | | | |
|---|---|---|---|---|---|---|
| | Default mode | 0.257 | -0.470 | 0.983 | 0.471 | 1.000 |
| | Dorsal Attention | -0.111 | -1.083 | 0.860 | 0.813 | 1.000 |
| | Ventral Attention | -0.006 | -0.578 | 0.567 | 0.984 | 1.000 |
| | Limbic | -0.291 | -1.402 | 0.820 | 0.591 | 1.000 |
| | Somatomotor | -0.692 | -1.311 | -0.074 | 0.031 | 1.000 |
| | Visual | -1.962 | -3.045 | -0.878 | 0.001 | 0.055 |
| *Scale 5* | Control | -1.113 | -1.999 | -0.226 | 0.016 | 0.769 |
| | Default mode | -0.808 | -1.884 | 0.268 | 0.133 | 1.000 |
| | Dorsal Attention | -0.415 | -1.193 | 0.364 | 0.282 | 1.000 |
| | Ventral Attention | -0.678 | -1.602 | 0.246 | 0.143 | 1.000 |
| | Limbic | 0.050 | -1.071 | 1.171 | 0.927 | 1.000 |
| | Somatomotor | -0.492 | -1.441 | 0.457 | 0.292 | 1.000 |
| | Visual | 0.692 | -0.194 | 1.578 | 0.120 | 1.000 |
| **Between-class comparison** | | | | | | |
| ***Psilocybin vs d-Amphetamine*** | | | | | | |
| ***Scale 1*** | **Control** | **1.965** | **1.197** | **2.733** | **<0.001** | **<0.001** |
| | **Default mode** | **2.261** | **1.460** | **3.062** | **<0.001** | **<0.001** |
| | **Dorsal Attention** | **1.522** | **0.639** | **2.406** | **0.001** | **0.040** |
| | Ventral Attention | 0.876 | 0.120 | 1.632 | 0.024 | 0.627 |
| | Limbic | 0.972 | -0.226 | 2.169 | 0.109 | 1.000 |
| | Somatomotor | -0.400 | -1.296 | 0.495 | 0.373 | 1.000 |
| | Visual | 0.272 | -0.865 | 1.409 | 0.633 | 1.000 |
| ***Scale 5*** | **Control** | **-2.163** | **-3.215** | **-1.110** | **<0.001** | **0.007** |
| | **Default mode** | **-1.983** | **-3.123** | **-0.843** | **0.001** | **0.040** |
| | Dorsal Attention | -0.870 | -1.934 | 0.195 | 0.106 | 1.000 |
| | **Ventral Attention** | **-1.801** | **-2.810** | **-0.792** | **<0.001** | **0.027** |
| | Limbic | -0.970 | -2.426 | 0.485 | 0.187 | 1.000 |
| | Somatomotor | -0.219 | -1.545 | 1.107 | 0.740 | 1.000 |
| | Visual | 0.027 | -1.170 | 1.224 | 0.964 | 1.000 |
| ***LSD vs d-Amphetamine*** | | | | | | |
| ***Scale 1*** | **Control** | **1.641** | **0.991** | **2.292** | **<0.001** | **0.001** |
| | **Default mode** | **1.564** | **0.971** | **2.157** | **<0.001** | **<0.001** |
| | **Dorsal Attention** | **1.229** | **0.650** | **1.807** | **<0.001** | **0.007** |
| | Ventral Attention | 0.652 | 0.188 | 1.115 | 0.008 | 0.210 |
| | Limbic | 0.332 | -0.562 | 1.226 | 0.452 | 1.000 |
| | Somatomotor | -0.946 | -1.501 | -0.391 | 0.002 | 0.060 |
| | Visual | -0.852 | -1.444 | -0.259 | 0.006 | 0.171 |
| ***Scale 5*** | **Control** | **-2.420** | **-3.217** | **-1.622** | **<0.001** | **<0.001** |
| | **Default mode** | **-2.367** | **-3.205** | **-1.529** | **<0.001** | **<0.001** |
| | Dorsal Attention | -0.972 | -1.718 | -0.227 | 0.013 | 0.293 |
| | **Ventral Attention** | **-1.670** | **-2.433** | **-0.907** | **<0.001** | **0.004** |
| | Limbic | -1.175 | -2.242 | -0.109 | 0.032 | 0.634 |

| | | | | | | |
|---|---|---|---|---|---|---|
| | Somatomotor | -0.227 | -0.964 | 0.511 | 0.535 | 1.000 |
| | Visual | 0.130 | -0.800 | 1.059 | 0.778 | 1.000 |
| ***Mescaline vs d-Amphetamine*** | | | | | | |
| ***Scale 1*** | **Control** | **1.754** | **0.914** | **2.594** | **<0.001** | **0.005** |
| | **Default mode** | **1.997** | **1.168** | **2.826** | **<0.001** | **<0.001** |
| | **Dorsal Attention** | **1.459** | **0.697** | **2.221** | **<0.001** | **0.015** |
| | Ventral Attention | 0.939 | 0.258 | 1.619 | 0.008 | 0.242 |
| | Limbic | 1.124 | 0.026 | 2.222 | 0.045 | 1.000 |
| | Somatomotor | -0.478 | -1.338 | 0.382 | 0.269 | 1.000 |
| | Visual | -1.179 | -2.212 | -0.147 | 0.026 | 0.702 |
| ***Scale 5*** | **Control** | **-2.446** | **-3.495** | **-1.397** | **<0.001** | **0.002** |
| | **Default mode** | **-1.924** | **-2.983** | **-0.865** | **<0.001** | **0.030** |
| | Dorsal Attention | -0.634 | -1.608 | 0.340 | 0.194 | 1.000 |
| | Ventral Attention | -1.536 | -2.578 | -0.495 | 0.005 | 0.154 |
| | Limbic | -0.549 | -2.075 | 0.978 | 0.473 | 1.000 |
| | Somatomotor | 0.364 | -0.790 | 1.519 | 0.527 | 1.000 |
| | Visual | 0.698 | -0.385 | 1.782 | 0.200 | 1.000 |
| ***Psilocybin vs MDMA*** | | | | | | |
| ***Scale 1*** | **Control** | **1.444** | **0.686** | **2.201** | **<0.001** | **0.020** |
| | **Default mode** | **2.004** | **1.188** | **2.821** | **<0.001** | **<0.001** |
| | Dorsal Attention | 1.634 | 0.613 | 2.655 | 0.002 | 0.115 |
| | Ventral Attention | 0.882 | 0.087 | 1.676 | 0.030 | 1.000 |
| | Limbic | 1.262 | 0.134 | 2.391 | 0.029 | 1.000 |
| | Somatomotor | 0.292 | -0.600 | 1.184 | 0.513 | 1.000 |
| | Visual | 2.234 | 0.721 | 3.746 | 0.005 | 0.217 |
| *Scale 5* | Control | -1.050 | -2.133 | 0.033 | 0.057 | 1.000 |
| | Default mode | -1.175 | -2.423 | 0.072 | 0.064 | 1.000 |
| | Dorsal Attention | -0.455 | -1.623 | 0.713 | 0.436 | 1.000 |
| | Ventral Attention | -1.123 | -2.210 | -0.035 | 0.043 | 1.000 |
| | Limbic | -1.021 | -2.576 | 0.535 | 0.194 | 1.000 |
| | Somatomotor | 0.273 | -1.172 | 1.718 | 0.706 | 1.000 |
| | Visual | -0.665 | -1.954 | 0.624 | 0.304 | 1.000 |
| ***LSD vs MDMA*** | | | | | | |
| *Scale 1* | Control | 1.120 | 0.490 | 1.750 | 0.001 | 0.060 |
| | **Default mode** | **1.307** | **0.690** | **1.925** | **<0.001** | **0.010** |
| | Dorsal Attention | 1.340 | 0.546 | 2.134 | 0.002 | 0.095 |
| | Ventral Attention | 0.657 | 0.110 | 1.205 | 0.021 | 0.892 |
| | Limbic | 0.623 | -0.195 | 1.440 | 0.129 | 1.000 |
| | Somatomotor | -0.253 | -0.811 | 0.305 | 0.352 | 1.000 |
| | Visual | 1.110 | -0.066 | 2.285 | 0.063 | 1.000 |
| *Scale 5* | Control | -1.307 | -2.141 | -0.473 | 0.004 | 0.170 |
| | Default mode | -1.559 | -2.521 | -0.597 | 0.003 | 0.134 |

| | | | | | | |
|---|---|---|---|---|---|---|
| | Dorsal Attention | -0.558 | -1.363 | 0.247 | 0.164 | 1.000 |
| | Ventral Attention | -0.992 | -1.849 | -0.135 | 0.025 | 1.000 |
| | Limbic | -1.226 | -2.431 | -0.021 | 0.046 | 1.000 |
| | Somatomotor | 0.265 | -0.655 | 1.186 | 0.557 | 1.000 |
| | Visual | -0.562 | -1.552 | 0.428 | 0.251 | 1.000 |
| ***Mescaline vs MDMA*** | | | | | | |
| *Scale 1* | Control | 1.232 | 0.397 | 2.067 | 0.005 | 0.219 |
| | **Default mode** | **1.741** | **0.892** | **2.589** | **<0.001** | **0.007** |
| | Dorsal Attention | 1.570 | 0.645 | 2.496 | 0.001 | 0.068 |
| | Ventral Attention | 0.944 | 0.219 | 1.670 | 0.012 | 0.543 |
| | Limbic | 1.415 | 0.388 | 2.441 | 0.008 | 0.376 |
| | Somatomotor | 0.214 | -0.639 | 1.068 | 0.616 | 1.000 |
| | Visual | 0.782 | -0.665 | 2.230 | 0.282 | 1.000 |
| *Scale 5* | Control | -1.333 | -2.418 | -0.248 | 0.017 | 0.767 |
| | Default mode | -1.116 | -2.296 | 0.064 | 0.063 | 1.000 |
| | Dorsal Attention | -0.219 | -1.287 | 0.848 | 0.681 | 1.000 |
| | Ventral Attention | -0.858 | -1.971 | 0.255 | 0.128 | 1.000 |
| | Limbic | -0.599 | -2.224 | 1.026 | 0.462 | 1.000 |
| | Somatomotor | 0.856 | -0.434 | 2.147 | 0.188 | 1.000 |
| | Visual | 0.007 | -1.169 | 1.182 | 0.991 | 1.000 |

*Lower and Upper denote the lower and upper 95% confidence intervals, respectively; p (FWER) reflect Holm-Bonferroni adjusted p-values across 53 tests within a contrast (e.g., psychedelics vs. stimulants)*
