## Supplemental Table 11 for "Psychedelics distinctly alter brain entropy and complexity compared to psychostimulants"

### *Network NSC*

| Contrast | Network | % Difference | Lower | Upper | p (unadjusted) | p (FWER) |
|---|---|---|---|---|---|---|
| **Drug vs placebo** | | | | | | |
| ***LSD*** | **Control** | **6.070** | **4.289** | **7.852** | **<0.001** | **<0.001** |
| | **Default mode** | **6.181** | **4.499** | **7.863** | **<0.001** | **<0.001** |
| | **Dorsal Attention** | **7.264** | **5.008** | **9.521** | **<0.001** | **<0.001** |
| | **Ventral Attention** | **3.300** | **1.787** | **4.813** | **<0.001** | **0.001** |
| | Limbic | -0.180 | -1.972 | 1.612 | 0.843 | 1.000 |
| | **Somatomotor** | **4.950** | **2.526** | **7.375** | **<0.001** | **0.003** |
| | **Visual** | **18.131** | **14.575** | **21.687** | **<0.001** | **<0.001** |
| *Psilocybin* | Control | 4.193 | 1.444 | 6.941 | 0.003 | 0.107 |
| | **Default mode** | **4.391** | **1.825** | **6.957** | **0.001** | **0.045** |
| | **Dorsal Attention** | **6.549** | **3.110** | **9.987** | **<0.001** | **0.015** |
| | Ventral Attention | 2.499 | 0.251 | 4.748 | 0.030 | 0.407 |
| | Limbic | 0.437 | -1.984 | 2.858 | 0.719 | 1.000 |
| | **Somatomotor** | **6.686** | **3.057** | **10.315** | **<0.001** | **0.022** |
| | **Visual** | **19.538** | **14.830** | **24.247** | **<0.001** | **<0.001** |
| ***Mescaline*** | **Control** | **4.673** | **1.960** | **7.386** | **0.001** | **0.046** |
| | **Default mode** | **4.720** | **2.275** | **7.164** | **<0.001** | **0.014** |
| | **Dorsal Attention** | **5.353** | **2.547** | **8.159** | **<0.001** | **0.016** |
| | Ventral Attention | 2.960 | 0.950 | 4.970 | 0.004 | 0.165 |
| | Limbic | 0.694 | -1.601 | 2.990 | 0.549 | 1.000 |
| | Somatomotor | 3.345 | 0.034 | 6.656 | 0.048 | 1.000 |
| | **Visual** | **11.926** | **7.984** | **15.868** | **<0.001** | **<0.001** |
| *MDMA* | Control | 3.778 | 1.179 | 6.378 | 0.006 | 0.256 |
| | Default mode | 3.944 | 1.205 | 6.683 | 0.006 | 0.272 |
| | Dorsal Attention | 5.227 | 2.006 | 8.449 | 0.002 | 0.115 |
| | **Ventral Attention** | **4.233** | **2.159** | **6.307** | **<0.001** | **0.011** |
| | Limbic | 1.122 | -1.242 | 3.485 | 0.346 | 1.000 |
| | **Somatomotor** | **11.006** | **7.199** | **14.812** | **<0.001** | **<0.001** |
| | **Visual** | **17.329** | **10.949** | **23.709** | **<0.001** | **<0.001** |
| *d-Amphetamine* | Control | 2.856 | 0.202 | 5.509 | 0.035 | 1.000 |
| | Default mode | 3.088 | 0.485 | 5.691 | 0.021 | 0.683 |
| | **Dorsal Attention** | **6.310** | **3.775** | **8.845** | **<0.001** | **<0.001** |
| | **Ventral Attention** | **4.966** | **3.079** | **6.853** | **<0.001** | **<0.001** |
| | Limbic | 0.698 | -1.139 | 2.535 | 0.448 | 1.000 |
| | **Somatomotor** | **14.581** | **11.105** | **18.057** | **<0.001** | **<0.001** |
| | **Visual** | **27.411** | **23.020** | **31.802** | **<0.001** | **<0.001** |
| **Within-class comparison** | | | | | | |

| | | | | | | |
|---|---|---|---|---|---|---|
| *LSD vs Psilocybin* | Control | 1.878 | -0.499 | 4.255 | 0.117 | 1.000 |
| | Default mode | 1.790 | -0.409 | 3.989 | 0.107 | 1.000 |
| | Dorsal Attention | 0.716 | -2.054 | 3.486 | 0.602 | 1.000 |
| | Ventral Attention | 0.801 | -1.205 | 2.806 | 0.423 | 1.000 |
| | Limbic | -0.617 | -2.753 | 1.520 | 0.563 | 1.000 |
| | Somatomotor | -1.736 | -4.724 | 1.253 | 0.246 | 1.000 |
| | Visual | -1.407 | -4.705 | 1.890 | 0.392 | 1.000 |
| *LSD vs Mescaline* | Control | 1.397 | -0.879 | 3.674 | 0.220 | 1.000 |
| | Default mode | 1.462 | -0.666 | 3.589 | 0.170 | 1.000 |
| | Dorsal Attention | 1.912 | -0.051 | 3.874 | 0.056 | 1.000 |
| | Ventral Attention | 0.340 | -1.095 | 1.775 | 0.633 | 1.000 |
| | Limbic | -0.874 | -2.441 | 0.693 | 0.265 | 1.000 |
| | Somatomotor | 1.605 | -0.610 | 3.821 | 0.148 | 1.000 |
| | **Visual** | **6.205** | **4.300** | **8.110** | **<0.001** | **<0.001** |
| *Mescaline vs Psilocybin* | Control | 0.481 | -1.812 | 2.773 | 0.670 | 1.000 |
| | Default mode | 0.329 | -2.042 | 2.699 | 0.778 | 1.000 |
| | Dorsal Attention | -1.196 | -3.875 | 1.484 | 0.364 | 1.000 |
| | Ventral Attention | 0.461 | -1.330 | 2.252 | 0.602 | 1.000 |
| | Limbic | 0.257 | -1.369 | 1.884 | 0.749 | 1.000 |
| | Somatomotor | -3.341 | -5.935 | -0.747 | 0.014 | 0.627 |
| | **Visual** | **-7.612** | **-10.860** | **-4.365** | **<0.001** | **0.003** |
| *MDMA vs d-Amphetamine* | Control | 0.923 | -1.277 | 3.122 | 0.395 | 1.000 |
| | Default mode | 0.856 | -1.370 | 3.082 | 0.434 | 1.000 |
| | Dorsal Attention | -1.083 | -4.543 | 2.378 | 0.523 | 1.000 |
| | Ventral Attention | -0.733 | -2.869 | 1.403 | 0.484 | 1.000 |
| | Limbic | 0.424 | -1.904 | 2.751 | 0.713 | 1.000 |
| | Somatomotor | -3.575 | -5.757 | -1.394 | 0.003 | 0.132 |
| | **Visual** | **-10.082** | **-15.289** | **-4.876** | **<0.001** | **0.030** |
| **Between-class comparison** | | | | | | |
| *Psilocybin vs d-Amphetamine* | Control | 1.337 | -1.702 | 4.376 | 0.382 | 1.000 |
| | Default mode | 1.303 | -1.710 | 4.317 | 0.389 | 1.000 |
| | Dorsal Attention | 0.239 | -3.134 | 3.611 | 0.888 | 1.000 |
| | Ventral Attention | -2.467 | -4.939 | 0.005 | 0.050 | 1.000 |
| | Limbic | -0.261 | -2.716 | 2.193 | 0.832 | 1.000 |
| | **Somatomotor** | **-7.895** | **-11.667** | **-4.123** | **<0.001** | **0.004** |
| | **Visual** | **-7.873** | **-12.279** | **-3.467** | **<0.001** | **0.027** |
| *LSD vs d-Amphetamine* | Control | 3.215 | 1.161 | 5.269 | 0.004 | 0.106 |
| | Default mode | 3.093 | 0.783 | 5.404 | 0.011 | 0.255 |
| | Dorsal Attention | 0.954 | -1.261 | 3.169 | 0.385 | 1.000 |
| | Ventral Attention | -1.666 | -3.515 | 0.182 | 0.076 | 1.000 |

| | | | | | | |
|---|---|---|---|---|---|---|
| | Limbic | -0.878 | -2.590 | 0.834 | 0.304 | 1.000 |
| | **Somatomotor** | **-9.631** | **-12.247** | **-7.015** | **<0.001** | **<0.001** |
| | **Visual** | **-9.281** | **-12.560** | **-6.001** | **<0.001** | **<0.001** |
| *Mescaline vs d-Amphetamine* | Control | 1.817 | -1.130 | 4.765 | 0.222 | 1.000 |
| | Default mode | 1.632 | -1.291 | 4.554 | 0.267 | 1.000 |
| | Dorsal Attention | -0.957 | -3.680 | 1.766 | 0.482 | 1.000 |
| | Ventral Attention | -2.006 | -4.194 | 0.182 | 0.072 | 1.000 |
| | Limbic | -0.004 | -2.163 | 2.155 | 0.997 | 1.000 |
| | **Somatomotor** | **-11.236** | **-14.539** | **-7.934** | **<0.001** | **<0.001** |
| | **Visual** | **-15.485** | **-19.202** | **-11.769** | **<0.001** | **<0.001** |
| *Psilocybin vs MDMA* | Control | 0.414 | -2.712 | 3.541 | 0.791 | 1.000 |
| | Default mode | 0.447 | -2.771 | 3.665 | 0.781 | 1.000 |
| | Dorsal Attention | 1.321 | -2.817 | 5.460 | 0.525 | 1.000 |
| | Ventral Attention | -1.734 | -4.363 | 0.895 | 0.191 | 1.000 |
| | Limbic | -0.685 | -3.354 | 1.984 | 0.609 | 1.000 |
| | Somatomotor | -4.320 | -8.434 | -0.206 | 0.040 | 1.000 |
| | Visual | 2.209 | -4.305 | 8.723 | 0.496 | 1.000 |
| *LSD vs MDMA* | Control | 2.292 | 0.038 | 4.547 | 0.047 | 1.000 |
| | Default mode | 2.237 | -0.357 | 4.831 | 0.088 | 1.000 |
| | Dorsal Attention | 2.037 | -1.299 | 5.373 | 0.223 | 1.000 |
| | Ventral Attention | -0.933 | -2.943 | 1.076 | 0.349 | 1.000 |
| | Limbic | -1.302 | -3.071 | 0.468 | 0.140 | 1.000 |
| | **Somatomotor** | **-6.055** | **-9.146** | **-2.965** | **<0.001** | **0.031** |
| | Visual | 0.802 | -5.018 | 6.622 | 0.778 | 1.000 |
| *Mescaline vs MDMA* | Control | 0.895 | -2.158 | 3.948 | 0.559 | 1.000 |
| | Default mode | 0.776 | -2.363 | 3.915 | 0.621 | 1.000 |
| | Dorsal Attention | 0.125 | -3.514 | 3.765 | 0.945 | 1.000 |
| | Ventral Attention | -1.273 | -3.627 | 1.081 | 0.282 | 1.000 |
| | Limbic | -0.428 | -2.704 | 1.849 | 0.707 | 1.000 |
| | **Somatomotor** | **-7.661** | **-11.367** | **-3.955** | **<0.001** | **0.009** |
| | Visual | -5.403 | -11.480 | 0.674 | 0.079 | 1.000 |

*Lower and Upper denote the lower and upper 95% confidence intervals, respectively; p (FWER) reflect Holm-Bonferroni adjusted p-values across 53 tests within a contrast (e.g., psychedelics vs. stimulants); NSC, Normalised spatial complexity*
