## Supplemental Table 12 for "Psychedelics distinctly alter brain entropy and complexity compared to psychostimulants"

| Dynamic condition correlation entropy | | | | | | | |
|---|---|---|---|---|---|---|---|
| Contrast | Network | Network | % Difference | Lower | Upper | p (unadjusted) | p (FWER) |
| **Drug vs. placebo** | | | | | | | |
| ***LSD*** | **Control** | **Control** | **1.125** | **0.390** | **1.861** | **0.003** | **0.094** |
| | Control | Default | 0.792 | 0.148 | 1.436 | 0.016 | 0.394 |
| | Control | Dorsal Attention | 0.591 | -0.055 | 1.237 | 0.072 | 1.000 |
| | Control | Limbic | 2.296 | 0.480 | 4.113 | 0.014 | 0.343 |
| | Control | Ventral Attention | 1.044 | 0.280 | 1.807 | 0.008 | 0.221 |
| | Control | Somatomotor | 0.393 | -0.276 | 1.063 | 0.247 | 1.000 |
| | Control | Visual | 1.153 | 0.355 | 1.950 | 0.005 | 0.155 |
| | **Default** | **Default** | **1.317** | **0.597** | **2.037** | **<0.001** | **0.016** |
| | Default | Dorsal Attention | 0.605 | -0.007 | 1.218 | 0.053 | 1.000 |
| | Default | Limbic | 2.434 | 0.856 | 4.013 | 0.003 | 0.090 |
| | Default | Ventral Attention | 0.482 | -0.146 | 1.111 | 0.131 | 1.000 |
| | Default | Somatomotor | 0.561 | -0.055 | 1.178 | 0.074 | 1.000 |
| | Default | Visual | 0.995 | 0.248 | 1.741 | 0.010 | 0.259 |
| | Dorsal Attention | Dorsal Attention | 0.230 | -0.787 | 1.246 | 0.655 | 1.000 |
| | Dorsal Attention | Limbic | 1.414 | -0.307 | 3.134 | 0.106 | 1.000 |
| | Dorsal Attention | Ventral Attention | 0.391 | -0.449 | 1.230 | 0.359 | 1.000 |
| | Dorsal Attention | Somatomotor | 0.220 | -0.655 | 1.095 | 0.619 | 1.000 |
| | Dorsal Attention | Visual | 0.731 | -0.019 | 1.482 | 0.056 | 1.000 |
| | Limbic | Limbic | -0.262 | -2.818 | 2.294 | 0.839 | 1.000 |
| | Limbic | Ventral Attention | 1.038 | -0.789 | 2.866 | 0.263 | 1.000 |
| | Limbic | Somatomotor | 1.202 | -0.685 | 3.088 | 0.209 | 1.000 |
| | Limbic | Visual | 1.677 | -0.131 | 3.485 | 0.069 | 1.000 |
| | Ventral Attention | Ventral Attention | 0.980 | -0.160 | 2.120 | 0.091 | 1.000 |
| | Ventral Attention | Somatomotor | 0.254 | -0.597 | 1.105 | 0.555 | 1.000 |
| | Ventral Attention | Visual | 0.505 | -0.419 | 1.429 | 0.281 | 1.000 |
| | Somatomotor | Somatomotor | -0.067 | -1.330 | 1.196 | 0.916 | 1.000 |
| | Somatomotor | Visual | 0.506 | -0.535 | 1.548 | 0.338 | 1.000 |
| | **Visual** | **Visual** | **2.441** | **1.280** | **3.603** | **<0.001** | **0.002** |
| ***Psilocybin*** | **Control** | **Control** | **1.715** | **0.746** | **2.683** | **<0.001** | **0.035** |
| | Control | Default | 1.123 | 0.353 | 1.893 | 0.005 | 0.128 |
| | Control | Dorsal Attention | 1.435 | 0.499 | 2.371 | 0.004 | 0.110 |
| | Control | Limbic | 4.127 | 1.647 | 6.608 | 0.002 | 0.066 |
| | Control | Ventral Attention | 1.374 | 0.485 | 2.263 | 0.003 | 0.106 |
| | Control | Somatomotor | 0.733 | -0.107 | 1.573 | 0.085 | 0.568 |
| | Control | Visual | 2.695 | 0.997 | 4.392 | 0.003 | 0.098 |
| | Default | Default | 1.836 | 0.704 | 2.968 | 0.002 | 0.080 |
| | Default | Dorsal Attention | 1.227 | 0.504 | 1.950 | 0.002 | 0.058 |
| | Default | Limbic | 4.044 | 1.480 | 6.608 | 0.003 | 0.107 |
| | Default | Ventral Attention | 1.210 | 0.408 | 2.013 | 0.004 | 0.128 |
| | Default | Somatomotor | 1.280 | 0.264 | 2.296 | 0.015 | 0.296 |
| | Default | Visual | 1.763 | 0.579 | 2.947 | 0.005 | 0.128 |
| | Dorsal Attention | Dorsal Attention | 1.491 | 0.077 | 2.905 | 0.039 | 0.434 |
| | Dorsal Attention | Limbic | 3.143 | 0.125 | 6.160 | 0.042 | 0.434 |

| | | | | | | | |
|---|---|---|---|---|---|---|---|
| | Dorsal Attention | Ventral Attention | 2.009 | 0.556 | 3.462 | 0.008 | 0.171 |
| | Dorsal Attention | Somatomotor | 1.516 | 0.221 | 2.812 | 0.024 | 0.353 |
| | Dorsal Attention | Visual | 2.467 | 0.823 | 4.112 | 0.005 | 0.128 |
| | Limbic | Limbic | 0.356 | -3.143 | 3.855 | 0.837 | 1.000 |
| | Limbic | Ventral Attention | 3.355 | 0.364 | 6.346 | 0.029 | 0.407 |
| | Limbic | Somatomotor | 3.082 | 0.093 | 6.072 | 0.044 | 0.434 |
| | Limbic | Visual | 3.674 | 0.753 | 6.596 | 0.015 | 0.296 |
| | Ventral Attention | Ventral Attention | 1.802 | -0.038 | 3.643 | 0.055 | 0.438 |
| | Ventral Attention | Somatomotor | 1.399 | 0.103 | 2.695 | 0.035 | 0.422 |
| | Ventral Attention | Visual | 2.296 | 0.779 | 3.814 | 0.005 | 0.128 |
| | Somatomotor | Somatomotor | 1.420 | -0.584 | 3.424 | 0.160 | 0.801 |
| | Somatomotor | Visual | 2.728 | 0.806 | 4.651 | 0.007 | 0.159 |
| | **Visual** | **Visual** | **4.227** | **2.583** | **5.872** | **<0.001** | **<0.001** |
| *Mescaline* | Control | Control | 1.144 | 0.171 | 2.116 | 0.022 | 0.709 |
| | Control | Default | 0.791 | -0.038 | 1.620 | 0.061 | 1.000 |
| | Control | Dorsal Attention | 0.383 | -0.174 | 0.941 | 0.173 | 1.000 |
| | Control | Limbic | 2.156 | -0.789 | 5.102 | 0.130 | 1.000 |
| | Control | Ventral Attention | 1.306 | 0.137 | 2.474 | 0.029 | 0.913 |
| | Control | Somatomotor | 0.293 | -0.343 | 0.929 | 0.359 | 1.000 |
| | Control | Visual | 0.433 | -0.019 | 0.885 | 0.060 | 1.000 |
| | Default | Default | 1.258 | 0.225 | 2.292 | 0.019 | 0.611 |
| | Default | Dorsal Attention | 0.733 | -0.025 | 1.491 | 0.058 | 1.000 |
| | Default | Limbic | 3.286 | 0.713 | 5.858 | 0.014 | 0.492 |
| | Default | Ventral Attention | 0.978 | -0.012 | 1.967 | 0.053 | 1.000 |
| | Default | Somatomotor | 0.633 | -0.344 | 1.610 | 0.198 | 1.000 |
| | Default | Visual | 0.500 | -0.063 | 1.062 | 0.081 | 1.000 |
| | Dorsal Attention | Dorsal Attention | 0.446 | -0.776 | 1.668 | 0.466 | 1.000 |
| | Dorsal Attention | Limbic | 1.073 | -1.835 | 3.981 | 0.442 | 1.000 |
| | Dorsal Attention | Ventral Attention | 0.561 | -0.401 | 1.523 | 0.247 | 1.000 |
| | Dorsal Attention | Somatomotor | 0.367 | -0.938 | 1.673 | 0.571 | 1.000 |
| | Dorsal Attention | Visual | 0.409 | -0.456 | 1.273 | 0.348 | 1.000 |
| | Limbic | Limbic | -0.026 | -3.381 | 3.328 | 0.987 | 1.000 |
| | Limbic | Ventral Attention | 3.566 | 1.010 | 6.121 | 0.008 | 0.276 |
| | Limbic | Somatomotor | 0.332 | -3.033 | 3.696 | 0.840 | 1.000 |
| | Limbic | Visual | 1.580 | -0.654 | 3.815 | 0.161 | 1.000 |
| | Ventral Attention | Ventral Attention | 1.702 | -0.143 | 3.548 | 0.070 | 1.000 |
| | Ventral Attention | Somatomotor | 0.748 | -0.541 | 2.037 | 0.248 | 1.000 |
| | Ventral Attention | Visual | 0.442 | -0.413 | 1.297 | 0.303 | 1.000 |
| | Somatomotor | Somatomotor | -0.891 | -2.972 | 1.190 | 0.390 | 1.000 |
| | Somatomotor | Visual | 0.144 | -1.118 | 1.406 | 0.819 | 1.000 |
| | Visual | Visual | 1.484 | 0.028 | 2.940 | 0.046 | 1.000 |
| *MDMA* | Control | Control | 0.659 | -0.470 | 1.788 | 0.242 | 1.000 |
| | Control | Default | 0.275 | -0.714 | 1.265 | 0.573 | 1.000 |
| | Control | Dorsal Attention | 0.702 | -0.484 | 1.887 | 0.234 | 1.000 |
| | Control | Limbic | 1.466 | -1.866 | 4.798 | 0.380 | 1.000 |
| | Control | Ventral Attention | 0.306 | -0.666 | 1.278 | 0.528 | 1.000 |
| | Control | Somatomotor | 1.219 | -0.325 | 2.762 | 0.116 | 1.000 |

| | | | | | | | |
|---|---|---|---|---|---|---|---|
| | Control | Visual | 0.913 | -0.364 | 2.190 | 0.152 | 1.000 |
| | Default | Default | 0.421 | -0.848 | 1.690 | 0.498 | 1.000 |
| | Default | Dorsal Attention | 0.350 | -0.687 | 1.387 | 0.495 | 1.000 |
| | Default | Limbic | -0.783 | -3.324 | 1.758 | 0.538 | 1.000 |
| | Default | Ventral Attention | 0.282 | -0.879 | 1.443 | 0.621 | 1.000 |
| | Default | Somatomotor | 1.672 | -0.336 | 3.680 | 0.098 | 1.000 |
| | Default | Visual | 0.814 | -0.727 | 2.354 | 0.285 | 1.000 |
| | Dorsal Attention | Dorsal Attention | 0.679 | -1.222 | 2.579 | 0.471 | 1.000 |
| | Dorsal Attention | Limbic | 0.105 | -3.158 | 3.368 | 0.948 | 1.000 |
| | Dorsal Attention | Ventral Attention | -0.199 | -1.571 | 1.173 | 0.770 | 1.000 |
| | Dorsal Attention | Somatomotor | 2.159 | -0.054 | 4.372 | 0.055 | 1.000 |
| | Dorsal Attention | Visual | 1.211 | -0.752 | 3.173 | 0.216 | 1.000 |
| | Limbic | Limbic | 1.803 | -3.686 | 7.291 | 0.503 | 1.000 |
| | Limbic | Ventral Attention | -1.987 | -4.729 | 0.754 | 0.150 | 1.000 |
| | Limbic | Somatomotor | 2.172 | -1.274 | 5.617 | 0.209 | 1.000 |
| | Limbic | Visual | 0.261 | -2.605 | 3.127 | 0.854 | 1.000 |
| | Ventral Attention | Ventral Attention | 0.857 | -0.724 | 2.438 | 0.278 | 1.000 |
| | Ventral Attention | Somatomotor | 1.942 | 0.225 | 3.659 | 0.028 | 1.000 |
| | Ventral Attention | Visual | -1.094 | -2.261 | 0.072 | 0.065 | 1.000 |
| | **Somatomotor** | **Somatomotor** | **3.988** | **1.795** | **6.181** | **<0.001** | **0.045** |
| | Somatomotor | Visual | 2.716 | 0.370 | 5.061 | 0.024 | 0.929 |
| | Visual | Visual | 2.870 | 0.626 | 5.114 | 0.014 | 0.569 |
| *d-Amphetamine* | Control | Control | -0.506 | -1.232 | 0.221 | 0.168 | 1.000 |
| | Control | Default | -0.861 | -1.366 | -0.356 | 0.001 | 0.050 |
| | Control | Dorsal Attention | -0.623 | -1.099 | -0.148 | 0.011 | 0.391 |
| | Control | Limbic | -1.440 | -4.165 | 1.284 | 0.292 | 1.000 |
| | Control | Ventral Attention | -0.720 | -1.454 | 0.015 | 0.055 | 1.000 |
| | Control | Somatomotor | 0.710 | -0.263 | 1.683 | 0.149 | 1.000 |
| | Control | Visual | 0.156 | -0.332 | 0.643 | 0.522 | 1.000 |
| | Default | Default | -1.197 | -1.996 | -0.397 | 0.005 | 0.176 |
| | Default | Dorsal Attention | -0.672 | -1.193 | -0.151 | 0.012 | 0.420 |
| | Default | Limbic | -2.344 | -4.780 | 0.093 | 0.059 | 1.000 |
| | Default | Ventral Attention | -0.725 | -1.273 | -0.177 | 0.010 | 0.376 |
| | Default | Somatomotor | 0.453 | -0.811 | 1.718 | 0.467 | 1.000 |
| | Default | Visual | -0.294 | -0.838 | 0.249 | 0.280 | 1.000 |
| | Dorsal Attention | Dorsal Attention | -1.121 | -2.159 | -0.082 | 0.035 | 1.000 |
| | Dorsal Attention | Limbic | -0.958 | -3.661 | 1.745 | 0.479 | 1.000 |
| | Dorsal Attention | Ventral Attention | -1.453 | -2.368 | -0.538 | 0.002 | 0.093 |
| | Dorsal Attention | Somatomotor | 0.662 | -0.695 | 2.018 | 0.330 | 1.000 |
| | Dorsal Attention | Visual | -0.120 | -0.849 | 0.609 | 0.742 | 1.000 |
| | Limbic | Limbic | 0.249 | -3.766 | 4.264 | 0.899 | 1.000 |
| | Limbic | Ventral Attention | -2.915 | -5.062 | -0.768 | 0.009 | 0.333 |
| | Limbic | Somatomotor | 0.796 | -2.478 | 4.069 | 0.624 | 1.000 |
| | Limbic | Visual | 1.724 | -1.783 | 5.232 | 0.325 | 1.000 |
| | Ventral Attention | Ventral Attention | -1.201 | -2.494 | 0.092 | 0.068 | 1.000 |
| | Ventral Attention | Somatomotor | 0.049 | -1.241 | 1.340 | 0.939 | 1.000 |
| | Ventral Attention | Visual | -1.138 | -2.181 | -0.096 | 0.033 | 1.000 |

| | | | | | | | |
|---|---|---|---|---|---|---|---|
| | Somatomotor | Somatomotor | 3.677 | 1.581 | 5.773 | 0.001 | 0.058 |
| | Somatomotor | Visual | 1.953 | 0.071 | 3.836 | 0.043 | 1.000 |
| | **Visual** | **Visual** | **4.065** | **2.171** | **5.958** | **<0.001** | **0.004** |
| **Within-class comparison** | | | | | | | |
| *LSD vs Psilocybin* | Control | Control | -0.590 | -1.545 | 0.365 | 0.219 | 1.000 |
| | Control | Default | -0.331 | -1.109 | 0.447 | 0.395 | 1.000 |
| | Control | Dorsal Attention | -0.844 | -1.856 | 0.168 | 0.100 | 1.000 |
| | Control | Limbic | -1.831 | -4.292 | 0.630 | 0.140 | 1.000 |
| | Control | Ventral Attention | -0.330 | -1.272 | 0.612 | 0.484 | 1.000 |
| | Control | Somatomotor | -0.340 | -1.306 | 0.627 | 0.483 | 1.000 |
| | Control | Visual | -1.542 | -3.244 | 0.160 | 0.074 | 1.000 |
| | Default | Default | -0.520 | -1.625 | 0.586 | 0.345 | 1.000 |
| | Default | Dorsal Attention | -0.622 | -1.434 | 0.190 | 0.129 | 1.000 |
| | Default | Limbic | -1.610 | -3.899 | 0.680 | 0.156 | 1.000 |
| | Default | Ventral Attention | -0.728 | -1.511 | 0.055 | 0.067 | 1.000 |
| | Default | Somatomotor | -0.719 | -1.761 | 0.324 | 0.170 | 1.000 |
| | Default | Visual | -0.768 | -1.952 | 0.415 | 0.196 | 1.000 |
| | Dorsal Attention | Dorsal Attention | -1.261 | -2.751 | 0.228 | 0.094 | 1.000 |
| | Dorsal Attention | Limbic | -1.729 | -4.436 | 0.978 | 0.200 | 1.000 |
| | Dorsal Attention | Ventral Attention | -1.619 | -3.155 | -0.083 | 0.040 | 1.000 |
| | Dorsal Attention | Somatomotor | -1.296 | -2.597 | 0.004 | 0.051 | 1.000 |
| | Dorsal Attention | Visual | -1.736 | -3.363 | -0.109 | 0.037 | 1.000 |
| | Limbic | Limbic | -0.618 | -3.635 | 2.399 | 0.674 | 1.000 |
| | Limbic | Ventral Attention | -2.317 | -4.929 | 0.295 | 0.079 | 1.000 |
| | Limbic | Somatomotor | -1.880 | -4.681 | 0.921 | 0.179 | 1.000 |
| | Limbic | Visual | -1.997 | -4.716 | 0.721 | 0.144 | 1.000 |
| | Ventral Attention | Ventral Attention | -0.823 | -2.533 | 0.888 | 0.334 | 1.000 |
| | Ventral Attention | Somatomotor | -1.145 | -2.435 | 0.146 | 0.080 | 1.000 |
| | Ventral Attention | Visual | -1.792 | -3.460 | -0.123 | 0.037 | 1.000 |
| | Somatomotor | Somatomotor | -1.487 | -3.302 | 0.327 | 0.105 | 1.000 |
| | Somatomotor | Visual | -2.222 | -4.104 | -0.340 | 0.023 | 1.000 |
| | Visual | Visual | -1.786 | -3.389 | -0.183 | 0.030 | 1.000 |
| *LSD vs Mescaline* | Control | Control | -0.018 | -0.950 | 0.913 | 0.968 | 1.000 |
| | Control | Default | 0.002 | -0.810 | 0.813 | 0.997 | 1.000 |
| | Control | Dorsal Attention | 0.208 | -0.389 | 0.804 | 0.488 | 1.000 |
| | Control | Limbic | 0.140 | -3.169 | 3.449 | 0.925 | 1.000 |
| | Control | Ventral Attention | -0.262 | -1.399 | 0.875 | 0.643 | 1.000 |
| | Control | Somatomotor | 0.100 | -0.642 | 0.843 | 0.787 | 1.000 |
| | Control | Visual | 0.720 | 0.031 | 1.409 | 0.041 | 1.000 |
| | Default | Default | 0.059 | -0.939 | 1.056 | 0.905 | 1.000 |
| | Default | Dorsal Attention | -0.128 | -0.907 | 0.652 | 0.742 | 1.000 |
| | Default | Limbic | -0.851 | -3.359 | 1.656 | 0.488 | 1.000 |
| | Default | Ventral Attention | -0.496 | -1.406 | 0.415 | 0.273 | 1.000 |
| | Default | Somatomotor | -0.072 | -1.029 | 0.886 | 0.880 | 1.000 |
| | Default | Visual | 0.495 | -0.154 | 1.144 | 0.132 | 1.000 |
| | Dorsal Attention | Dorsal Attention | -0.216 | -1.422 | 0.989 | 0.718 | 1.000 |
| | Dorsal Attention | Limbic | 0.341 | -2.610 | 3.292 | 0.800 | 1.000 |

| | | | | | | | |
|---|---|---|---|---|---|---|---|
| | Dorsal Attention | Ventral Attention | -0.171 | -0.905 | 0.563 | 0.638 | 1.000 |
| | Dorsal Attention | Somatomotor | -0.147 | -1.381 | 1.087 | 0.809 | 1.000 |
| | Dorsal Attention | Visual | 0.323 | -0.303 | 0.948 | 0.299 | 1.000 |
| | Limbic | Limbic | -0.235 | -3.105 | 2.634 | 0.868 | 1.000 |
| | Limbic | Ventral Attention | -2.528 | -5.078 | 0.023 | 0.052 | 1.000 |
| | Limbic | Somatomotor | 0.870 | -2.189 | 3.930 | 0.558 | 1.000 |
| | Limbic | Visual | 0.097 | -2.108 | 2.302 | 0.929 | 1.000 |
| | Ventral Attention | Ventral Attention | -0.723 | -2.282 | 0.837 | 0.351 | 1.000 |
| | Ventral Attention | Somatomotor | -0.493 | -1.704 | 0.717 | 0.413 | 1.000 |
| | Ventral Attention | Visual | 0.063 | -0.714 | 0.840 | 0.870 | 1.000 |
| | Somatomotor | Somatomotor | 0.824 | -0.929 | 2.577 | 0.343 | 1.000 |
| | Somatomotor | Visual | 0.362 | -0.588 | 1.312 | 0.442 | 1.000 |
| | Visual | Visual | 0.957 | -0.048 | 1.963 | 0.061 | 1.000 |
| *Mescaline vs Psilocybin* | Control | Control | -0.571 | -1.707 | 0.564 | 0.312 | 1.000 |
| | Control | Default | -0.332 | -1.290 | 0.626 | 0.483 | 1.000 |
| | Control | Dorsal Attention | -1.052 | -1.994 | -0.109 | 0.030 | 1.000 |
| | Control | Limbic | -1.971 | -5.159 | 1.217 | 0.186 | 1.000 |
| | Control | Ventral Attention | -0.068 | -1.332 | 1.195 | 0.913 | 1.000 |
| | Control | Somatomotor | -0.440 | -1.331 | 0.451 | 0.321 | 1.000 |
| | Control | Visual | -2.262 | -3.906 | -0.618 | 0.009 | 0.437 |
| | Default | Default | -0.578 | -1.880 | 0.724 | 0.369 | 1.000 |
| | Default | Dorsal Attention | -0.494 | -1.296 | 0.308 | 0.216 | 1.000 |
| | Default | Limbic | -0.758 | -3.254 | 1.737 | 0.527 | 1.000 |
| | Default | Ventral Attention | -0.232 | -1.268 | 0.803 | 0.643 | 1.000 |
| | Default | Somatomotor | -0.647 | -1.576 | 0.282 | 0.163 | 1.000 |
| | Default | Visual | -1.264 | -2.280 | -0.248 | 0.017 | 0.753 |
| | Dorsal Attention | Dorsal Attention | -1.045 | -2.442 | 0.351 | 0.135 | 1.000 |
| | Dorsal Attention | Limbic | -2.070 | -5.729 | 1.590 | 0.242 | 1.000 |
| | Dorsal Attention | Ventral Attention | -1.448 | -2.956 | 0.059 | 0.059 | 1.000 |
| | Dorsal Attention | Somatomotor | -1.149 | -2.538 | 0.239 | 0.099 | 1.000 |
| | Dorsal Attention | Visual | -2.059 | -3.589 | -0.528 | 0.010 | 0.497 |
| | Limbic | Limbic | -0.383 | -3.945 | 3.180 | 0.826 | 1.000 |
| | Limbic | Ventral Attention | 0.211 | -2.731 | 3.152 | 0.883 | 1.000 |
| | Limbic | Somatomotor | -2.751 | -5.516 | 0.015 | 0.051 | 1.000 |
| | Limbic | Visual | -2.094 | -5.169 | 0.981 | 0.172 | 1.000 |
| | Ventral Attention | Ventral Attention | -0.100 | -2.127 | 1.927 | 0.920 | 1.000 |
| | Ventral Attention | Somatomotor | -0.651 | -2.091 | 0.789 | 0.360 | 1.000 |
| | Ventral Attention | Visual | -1.854 | -3.265 | -0.444 | 0.013 | 0.602 |
| | Somatomotor | Somatomotor | -2.311 | -4.066 | -0.556 | 0.012 | 0.572 |
| | Somatomotor | Visual | -2.584 | -4.747 | -0.421 | 0.022 | 0.948 |
| | Visual | Visual | -2.743 | -4.441 | -1.045 | 0.002 | 0.128 |
| *MDMA vs d-Amphetamine* | Control | Control | 1.165 | 0.037 | 2.293 | 0.044 | 1.000 |
| | Control | Default | 1.136 | 0.214 | 2.058 | 0.018 | 0.852 |
| | Control | Dorsal Attention | 1.325 | 0.171 | 2.480 | 0.026 | 1.000 |
| | Control | Limbic | 2.906 | 0.100 | 5.712 | 0.043 | 1.000 |
| | Control | Ventral Attention | 1.025 | 0.160 | 1.891 | 0.022 | 1.000 |
| | Control | Somatomotor | 0.509 | -1.295 | 2.313 | 0.565 | 1.000 |

| | | | | | | | |
|---|---|---|---|---|---|---|---|
| | Control | Visual | 0.757 | -0.540 | 2.054 | 0.236 | 1.000 |
| | Default | Default | 1.618 | 0.470 | 2.766 | 0.009 | 0.417 |
| | Default | Dorsal Attention | 1.022 | 0.079 | 1.965 | 0.035 | 1.000 |
| | Default | Limbic | 1.561 | -0.572 | 3.693 | 0.145 | 1.000 |
| | Default | Ventral Attention | 1.007 | -0.056 | 2.070 | 0.062 | 1.000 |
| | Default | Somatomotor | 1.219 | -1.080 | 3.517 | 0.282 | 1.000 |
| | Default | Visual | 1.108 | -0.362 | 2.578 | 0.131 | 1.000 |
| | Dorsal Attention | Dorsal Attention | 1.800 | 0.008 | 3.591 | 0.049 | 1.000 |
| | Dorsal Attention | Limbic | 1.063 | -1.573 | 3.698 | 0.412 | 1.000 |
| | Dorsal Attention | Ventral Attention | 1.254 | -0.035 | 2.543 | 0.056 | 1.000 |
| | Dorsal Attention | Somatomotor | 1.498 | -1.058 | 4.053 | 0.239 | 1.000 |
| | Dorsal Attention | Visual | 1.331 | -0.450 | 3.111 | 0.136 | 1.000 |
| | Limbic | Limbic | 1.553 | -2.703 | 5.810 | 0.450 | 1.000 |
| | Limbic | Ventral Attention | 0.928 | -1.455 | 3.311 | 0.429 | 1.000 |
| | Limbic | Somatomotor | 1.376 | -1.595 | 4.347 | 0.347 | 1.000 |
| | Limbic | Visual | -1.464 | -5.157 | 2.230 | 0.420 | 1.000 |
| | Ventral Attention | Ventral Attention | 2.058 | 0.198 | 3.918 | 0.032 | 1.000 |
| | Ventral Attention | Somatomotor | 1.893 | -0.151 | 3.936 | 0.068 | 1.000 |
| | Ventral Attention | Visual | 0.044 | -0.528 | 0.616 | 0.876 | 1.000 |
| | Somatomotor | Somatomotor | 0.311 | -2.162 | 2.784 | 0.792 | 1.000 |
| | Somatomotor | Visual | 0.762 | -1.453 | 2.978 | 0.485 | 1.000 |
| | Visual | Visual | -1.195 | -3.208 | 0.818 | 0.231 | 1.000 |
| **Between-class comparison** | | | | | | | |
| ***Psilocybin vs d-Amphetamin*** | **Control** | **Control** | **2.221** | **1.231** | **3.210** | **<0.001** | **0.002** |
| | **Control** | **Default** | **1.984** | **1.232** | **2.736** | **<0.001** | **<0.001** |
| | **Control** | **Dorsal Attention** | **2.059** | **1.128** | **2.990** | **<0.001** | **0.003** |
| | **Control** | **Limbic** | **5.567** | **2.391** | **8.744** | **<0.001** | **0.032** |
| | **Control** | **Ventral Attention** | **2.093** | **1.147** | **3.040** | **<0.001** | **0.002** |
| | Control | Somatomotor | 0.023 | -1.160 | 1.205 | 0.970 | 1.000 |
| | Control | Visual | 2.539 | 0.831 | 4.246 | 0.005 | 0.135 |
| | **Default** | **Default** | **3.033** | **1.820** | **4.246** | **<0.001** | **<0.001** |
| | **Default** | **Dorsal Attention** | **1.899** | **1.147** | **2.651** | **<0.001** | **<0.001** |
| | **Default** | **Limbic** | **6.387** | **3.374** | **9.401** | **<0.001** | **0.005** |
| | **Default** | **Ventral Attention** | **1.935** | **1.136** | **2.734** | **<0.001** | **0.001** |
| | Default | Somatomotor | 0.827 | -0.652 | 2.305 | 0.265 | 1.000 |
| | **Default** | **Visual** | **2.058** | **0.857** | **3.258** | **0.001** | **0.041** |
| | **Dorsal Attention** | **Dorsal Attention** | **2.612** | **1.098** | **4.125** | **0.001** | **0.040** |
| | Dorsal Attention | Limbic | 4.100 | 0.616 | 7.585 | 0.022 | 0.598 |
| | **Dorsal Attention** | **Ventral Attention** | **3.463** | **1.919** | **5.006** | **<0.001** | **0.002** |
| | Dorsal Attention | Somatomotor | 0.855 | -0.798 | 2.508 | 0.302 | 1.000 |
| | Dorsal Attention | Visual | 2.587 | 0.913 | 4.261 | 0.003 | 0.105 |
| | Limbic | Limbic | 0.107 | -4.332 | 4.547 | 0.961 | 1.000 |
| | **Limbic** | **Ventral Attention** | **6.270** | **3.216** | **9.324** | **<0.001** | **0.008** |
| | Limbic | Somatomotor | 2.286 | -1.690 | 6.263 | 0.253 | 1.000 |
| | Limbic | Visual | 1.950 | -2.102 | 6.002 | 0.338 | 1.000 |
| | Ventral Attention | Ventral Attention | 3.003 | 1.044 | 4.962 | 0.003 | 0.105 |
| | Ventral Attention | Somatomotor | 1.350 | -0.261 | 2.960 | 0.098 | 1.000 |

| | | | | | | | |
|---|---|---|---|---|---|---|---|
| | **Ventral Attention** | **Visual** | **3.435** | **1.749** | **5.121** | **<0.001** | **0.008** |
| | Somatomotor | Somatomotor | -2.257 | -4.789 | 0.276 | 0.079 | 1.000 |
| | Somatomotor | Visual | 0.775 | -1.655 | 3.205 | 0.523 | 1.000 |
| | Visual | Visual | 0.163 | -2.015 | 2.341 | 0.881 | 1.000 |
| ***LSD vs d-Amphetamine*** | **Control** | **Control** | **1.631** | **0.869** | **2.393** | **<0.001** | **0.003** |
| | **Control** | **Default** | **1.653** | **1.034** | **2.272** | **<0.001** | **<0.001** |
| | **Control** | **Dorsal Attention** | **1.215** | **0.624** | **1.805** | **<0.001** | **0.004** |
| | Control | Limbic | 3.736 | 1.205 | 6.268 | 0.005 | 0.153 |
| | **Control** | **Ventral Attention** | **1.763** | **0.935** | **2.591** | **<0.001** | **0.003** |
| | Control | Somatomotor | -0.317 | -0.957 | 0.323 | 0.315 | 1.000 |
| | Control | Visual | 0.997 | 0.177 | 1.816 | 0.018 | 0.373 |
| | **Default** | **Default** | **2.513** | **1.641** | **3.385** | **<0.001** | **<0.001** |
| | **Default** | **Dorsal Attention** | **1.277** | **0.686** | **1.868** | **<0.001** | **0.002** |
| | **Default** | **Limbic** | **4.778** | **2.556** | **7.000** | **<0.001** | **0.005** |
| | **Default** | **Ventral Attention** | **1.207** | **0.638** | **1.776** | **<0.001** | **0.006** |
| | Default | Somatomotor | 0.108 | -1.169 | 1.385 | 0.863 | 1.000 |
| | Default | Visual | 1.289 | 0.500 | 2.078 | 0.002 | 0.056 |
| | Dorsal Attention | Dorsal Attention | 1.350 | 0.370 | 2.330 | 0.008 | 0.210 |
| | Dorsal Attention | Limbic | 2.372 | -0.112 | 4.855 | 0.060 | 1.000 |
| | **Dorsal Attention** | **Ventral Attention** | **1.844** | **1.023** | **2.664** | **<0.001** | **0.002** |
| | Dorsal Attention | Somatomotor | -0.441 | -1.721 | 0.838 | 0.485 | 1.000 |
| | Dorsal Attention | Visual | 0.851 | 0.049 | 1.653 | 0.038 | 0.718 |
| | Limbic | Limbic | -0.511 | -4.193 | 3.171 | 0.773 | 1.000 |
| | **Limbic** | **Ventral Attention** | **3.953** | **2.050** | **5.856** | **<0.001** | **0.007** |
| | Limbic | Somatomotor | 0.406 | -2.873 | 3.685 | 0.802 | 1.000 |
| | Limbic | Visual | -0.047 | -3.506 | 3.411 | 0.978 | 1.000 |
| | Ventral Attention | Ventral Attention | 2.180 | 0.839 | 3.522 | 0.002 | 0.068 |
| | Ventral Attention | Somatomotor | 0.205 | -1.159 | 1.569 | 0.761 | 1.000 |
| | Ventral Attention | Visual | 1.643 | 0.458 | 2.828 | 0.007 | 0.199 |
| | **Somatomotor** | **Somatomotor** | **-3.744** | **-5.741** | **-1.747** | **<0.001** | **0.033** |
| | Somatomotor | Visual | -1.447 | -3.313 | 0.420 | 0.122 | 1.000 |
| | Visual | Visual | -1.623 | -3.370 | 0.124 | 0.067 | 1.000 |
| ***Mescaline vs d-Amphetamine*** | **Control** | **Control** | **1.649** | **0.674** | **2.625** | **0.001** | **0.049** |
| | **Control** | **Default** | **1.652** | **0.856** | **2.447** | **<0.001** | **0.007** |
| | **Control** | **Dorsal Attention** | **1.007** | **0.484** | **1.530** | **<0.001** | **0.014** |
| | Control | Limbic | 3.596 | 0.050 | 7.142 | 0.047 | 1.000 |
| | **Control** | **Ventral Attention** | **2.025** | **0.837** | **3.213** | **0.001** | **0.048** |
| | Control | Somatomotor | -0.417 | -1.426 | 0.591 | 0.410 | 1.000 |
| | Control | Visual | 0.277 | -0.241 | 0.795 | 0.287 | 1.000 |
| | **Default** | **Default** | **2.455** | **1.339** | **3.571** | **<0.001** | **0.004** |
| | **Default** | **Dorsal Attention** | **1.405** | **0.637** | **2.173** | **<0.001** | **0.025** |
| | **Default** | **Limbic** | **5.629** | **2.522** | **8.736** | **<0.001** | **0.029** |
| | **Default** | **Ventral Attention** | **1.703** | **0.738** | **2.667** | **0.001** | **0.048** |
| | Default | Somatomotor | 0.180 | -1.265 | 1.624 | 0.803 | 1.000 |
| | Default | Visual | 0.794 | 0.204 | 1.384 | 0.009 | 0.276 |
| | Dorsal Attention | Dorsal Attention | 1.567 | 0.283 | 2.851 | 0.018 | 0.501 |
| | Dorsal Attention | Limbic | 2.031 | -1.442 | 5.504 | 0.238 | 1.000 |

| | | | | | | | |
|---|---|---|---|---|---|---|---|
| | **Dorsal Attention** | **Ventral Attention** | **2.014** | **1.063** | **2.966** | **<0.001** | **0.004** |
| | Dorsal Attention | Somatomotor | -0.294 | -1.920 | 1.331 | 0.717 | 1.000 |
| | Dorsal Attention | Visual | 0.529 | -0.355 | 1.413 | 0.237 | 1.000 |
| | Limbic | Limbic | -0.276 | -4.600 | 4.048 | 0.897 | 1.000 |
| | Limbic | Ventral Attention | 6.481 | 3.707 | 9.255 | <0.001 | 0.001 |
| | Limbic | Somatomotor | -0.464 | -4.677 | 3.748 | 0.825 | 1.000 |
| | Limbic | Visual | -0.144 | -3.810 | 3.522 | 0.937 | 1.000 |
| | Ventral Attention | Ventral Attention | 2.903 | 0.995 | 4.811 | 0.004 | 0.127 |
| | Ventral Attention | Somatomotor | 0.698 | -0.888 | 2.284 | 0.380 | 1.000 |
| | Ventral Attention | Visual | 1.581 | 0.444 | 2.717 | 0.007 | 0.230 |
| | **Somatomotor** | **Somatomotor** | **-4.568** | **-7.077** | **-2.059** | **<0.001** | **0.028** |
| | Somatomotor | Visual | -1.809 | -3.759 | 0.141 | 0.068 | 1.000 |
| | Visual | Visual | -2.580 | -4.500 | -0.660 | 0.010 | 0.281 |
| *Psilocybin vs MDMA* | Control | Control | 1.056 | -0.262 | 2.373 | 0.113 | 1.000 |
| | Control | Default | 0.848 | -0.287 | 1.982 | 0.139 | 1.000 |
| | Control | Dorsal Attention | 0.733 | -0.680 | 2.147 | 0.301 | 1.000 |
| | Control | Limbic | 2.661 | -1.071 | 6.393 | 0.159 | 1.000 |
| | Control | Ventral Attention | 1.068 | -0.086 | 2.222 | 0.069 | 1.000 |
| | Control | Somatomotor | -0.486 | -2.166 | 1.194 | 0.560 | 1.000 |
| | Control | Visual | 1.782 | -0.278 | 3.842 | 0.088 | 1.000 |
| | Default | Default | 1.415 | -0.129 | 2.960 | 0.071 | 1.000 |
| | Default | Dorsal Attention | 0.877 | -0.292 | 2.046 | 0.138 | 1.000 |
| | Default | Limbic | 4.827 | 1.596 | 8.058 | 0.004 | 0.213 |
| | Default | Ventral Attention | 0.928 | -0.351 | 2.207 | 0.150 | 1.000 |
| | Default | Somatomotor | -0.392 | -2.541 | 1.757 | 0.711 | 1.000 |
| | Default | Visual | 0.949 | -0.915 | 2.814 | 0.310 | 1.000 |
| | Dorsal Attention | Dorsal Attention | 0.812 | -1.382 | 3.007 | 0.459 | 1.000 |
| | Dorsal Attention | Limbic | 3.038 | -1.007 | 7.082 | 0.138 | 1.000 |
| | Dorsal Attention | Ventral Attention | 2.208 | 0.363 | 4.054 | 0.020 | 0.900 |
| | Dorsal Attention | Somatomotor | -0.643 | -3.044 | 1.758 | 0.591 | 1.000 |
| | Dorsal Attention | Visual | 1.257 | -1.171 | 3.685 | 0.303 | 1.000 |
| | Limbic | Limbic | -1.446 | -7.187 | 4.294 | 0.609 | 1.000 |
| | Limbic | Ventral Attention | 5.342 | 1.718 | 8.967 | 0.005 | 0.223 |
| | Limbic | Somatomotor | 0.910 | -3.284 | 5.105 | 0.664 | 1.000 |
| | Limbic | Visual | 3.413 | -0.184 | 7.011 | 0.062 | 1.000 |
| | Ventral Attention | Ventral Attention | 0.945 | -1.210 | 3.101 | 0.382 | 1.000 |
| | Ventral Attention | Somatomotor | -0.543 | -2.513 | 1.427 | 0.580 | 1.000 |
| | **Ventral Attention** | **Visual** | **3.391** | **1.621** | **5.161** | **<0.001** | **0.022** |
| | Somatomotor | Somatomotor | -2.568 | -5.157 | 0.021 | 0.052 | 1.000 |
| | Somatomotor | Visual | 0.013 | -2.768 | 2.793 | 0.993 | 1.000 |
| | Visual | Visual | 1.358 | -1.148 | 3.863 | 0.281 | 1.000 |
| *LSD vs MDMA* | Control | Control | 0.466 | -0.718 | 1.651 | 0.429 | 1.000 |
| | Control | Default | 0.517 | -0.538 | 1.572 | 0.325 | 1.000 |
| | Control | Dorsal Attention | -0.111 | -1.369 | 1.148 | 0.859 | 1.000 |
| | Control | Limbic | 0.830 | -2.331 | 3.991 | 0.596 | 1.000 |
| | Control | Ventral Attention | 0.738 | -0.295 | 1.770 | 0.157 | 1.000 |
| | Control | Somatomotor | -0.826 | -2.356 | 0.705 | 0.275 | 1.000 |

| | | | | | | | |
|---|---|---|---|---|---|---|---|
| | Control | Visual | 0.240 | -1.261 | 1.740 | 0.748 | 1.000 |
| | Default | Default | 0.896 | -0.378 | 2.169 | 0.157 | 1.000 |
| | Default | Dorsal Attention | 0.255 | -0.844 | 1.355 | 0.640 | 1.000 |
| | Default | Limbic | 3.217 | 0.629 | 5.806 | 0.016 | 0.726 |
| | Default | Ventral Attention | 0.200 | -0.947 | 1.347 | 0.720 | 1.000 |
| | Default | Somatomotor | -1.111 | -3.125 | 0.904 | 0.264 | 1.000 |
| | Default | Visual | 0.181 | -1.491 | 1.853 | 0.826 | 1.000 |
| | Dorsal Attention | Dorsal Attention | -0.449 | -2.416 | 1.517 | 0.644 | 1.000 |
| | Dorsal Attention | Limbic | 1.309 | -1.975 | 4.593 | 0.424 | 1.000 |
| | Dorsal Attention | Ventral Attention | 0.590 | -0.764 | 1.944 | 0.381 | 1.000 |
| | Dorsal Attention | Somatomotor | -1.939 | -4.085 | 0.207 | 0.074 | 1.000 |
| | Dorsal Attention | Visual | -0.479 | -2.450 | 1.492 | 0.622 | 1.000 |
| | Limbic | Limbic | -2.064 | -7.244 | 3.115 | 0.412 | 1.000 |
| | Limbic | Ventral Attention | 3.025 | 0.175 | 5.876 | 0.038 | 1.000 |
| | Limbic | Somatomotor | -0.970 | -4.630 | 2.690 | 0.594 | 1.000 |
| | Limbic | Visual | 1.416 | -1.465 | 4.298 | 0.324 | 1.000 |
| | Ventral Attention | Ventral Attention | 0.123 | -1.483 | 1.728 | 0.877 | 1.000 |
| | Ventral Attention | Somatomotor | -1.688 | -3.324 | -0.051 | 0.044 | 1.000 |
| | Ventral Attention | Visual | 1.599 | 0.282 | 2.917 | 0.019 | 0.816 |
| | **Somatomotor** | **Somatomotor** | **-4.055** | **-6.077** | **-2.033** | **<0.001** | **0.028** |
| | Somatomotor | Visual | -2.209 | -4.300 | -0.118 | 0.039 | 1.000 |
| | Visual | Visual | -0.428 | -2.563 | 1.706 | 0.683 | 1.000 |
| *Mescaline vs MDMA* | Control | Control | 0.484 | -0.829 | 1.798 | 0.461 | 1.000 |
| | Control | Default | 0.515 | -0.648 | 1.679 | 0.376 | 1.000 |
| | Control | Dorsal Attention | -0.318 | -1.535 | 0.898 | 0.596 | 1.000 |
| | Control | Limbic | 0.690 | -3.344 | 4.724 | 0.728 | 1.000 |
| | Control | Ventral Attention | 1.000 | -0.353 | 2.352 | 0.144 | 1.000 |
| | Control | Somatomotor | -0.926 | -2.513 | 0.661 | 0.241 | 1.000 |
| | Control | Visual | -0.480 | -1.792 | 0.832 | 0.458 | 1.000 |
| | Default | Default | 0.837 | -0.638 | 2.312 | 0.257 | 1.000 |
| | Default | Dorsal Attention | 0.383 | -0.801 | 1.567 | 0.518 | 1.000 |
| | Default | Limbic | 4.069 | 0.809 | 7.328 | 0.016 | 0.706 |
| | Default | Ventral Attention | 0.696 | -0.677 | 2.068 | 0.311 | 1.000 |
| | Default | Somatomotor | -1.039 | -3.166 | 1.088 | 0.325 | 1.000 |
| | Default | Visual | -0.314 | -1.897 | 1.269 | 0.686 | 1.000 |
| | Dorsal Attention | Dorsal Attention | -0.233 | -2.304 | 1.838 | 0.821 | 1.000 |
| | Dorsal Attention | Limbic | 0.968 | -3.015 | 4.951 | 0.623 | 1.000 |
| | Dorsal Attention | Ventral Attention | 0.760 | -0.671 | 2.192 | 0.289 | 1.000 |
| | Dorsal Attention | Somatomotor | -1.792 | -4.177 | 0.593 | 0.137 | 1.000 |
| | Dorsal Attention | Visual | -0.802 | -2.815 | 1.212 | 0.422 | 1.000 |
| | Limbic | Limbic | -1.829 | -7.514 | 3.856 | 0.515 | 1.000 |
| | **Limbic** | **Ventral Attention** | **5.553** | **2.210** | **8.896** | **0.002** | **0.083** |
| | Limbic | Somatomotor | -1.840 | -6.307 | 2.626 | 0.409 | 1.000 |
| | Limbic | Visual | 1.319 | -1.828 | 4.467 | 0.402 | 1.000 |
| | Ventral Attention | Ventral Attention | 0.845 | -1.269 | 2.959 | 0.425 | 1.000 |
| | Ventral Attention | Somatomotor | -1.195 | -3.128 | 0.739 | 0.219 | 1.000 |
| | Ventral Attention | Visual | 1.537 | 0.264 | 2.810 | 0.019 | 0.822 |

| | | | | | | | |
|---|---|---|---|---|---|---|---|
| | **Somatomotor** | **Somatomotor** | **-4.879** | **-7.447** | **-2.311** | **<0.001** | **0.022** |
| | Somatomotor | Visual | -2.571 | -4.878 | -0.265 | 0.030 | 1.000 |
| | Visual | Visual | -1.385 | -3.673 | 0.903 | 0.227 | 1.000 |

*Lower and Upper denote the lower and upper 95% confidence intervals, respectively; p (FWER) reflect Holm-Bonferroni adjusted p-values across 53 tests within a contrast (e.g., psychedelics vs. stimulants)*
