## Supplemental Table 13 for "Psychedelics distinctly alter brain entropy and complexity compared to psychostimulants"

| region_name | network | n_sessions_surv | survival_percentage | quality_flag |
|---|---|---|---|---|
| 7Networks_LH_Cont_Par_1 | Cont | 269 | 100 | OK |
| 7Networks_LH_Cont_Par_2 | Cont | 269 | 100 | OK |
| 7Networks_LH_Cont_Par_3 | Cont | 269 | 100 | OK |
| 7Networks_LH_Cont_Temp_1 | Cont | 122 | 45 | POOR |
| 7Networks_LH_Cont_OFC_1 | Cont | 265 | 98 | OK |
| 7Networks_LH_Cont_PFCl_1 | Cont | 269 | 100 | OK |
| 7Networks_LH_Cont_PFCl_2 | Cont | 269 | 100 | OK |
| 7Networks_LH_Cont_PFCl_3 | Cont | 269 | 100 | OK |
| 7Networks_LH_Cont_PFCl_4 | Cont | 269 | 100 | OK |
| 7Networks_LH_Cont_PFCl_5 | Cont | 269 | 100 | OK |
| 7Networks_LH_Cont_pCun_1 | Cont | 269 | 100 | OK |
| 7Networks_LH_Cont_Cing_1 | Cont | 269 | 100 | OK |
| 7Networks_LH_Cont_Cing_2 | Cont | 269 | 100 | OK |
| 7Networks_RH_Cont_Par_1 | Cont | 269 | 100 | OK |
| 7Networks_RH_Cont_Par_2 | Cont | 269 | 100 | OK |
| 7Networks_RH_Cont_Par_3 | Cont | 269 | 100 | OK |
| 7Networks_RH_Cont_Temp_1 | Cont | 99 | 36 | POOR |
| 7Networks_RH_Cont_PFCv_1 | Cont | 269 | 100 | OK |
| 7Networks_RH_Cont_PFCl_1 | Cont | 269 | 100 | OK |
| 7Networks_RH_Cont_PFCl_2 | Cont | 269 | 100 | OK |
| 7Networks_RH_Cont_PFCl_3 | Cont | 269 | 100 | OK |
| 7Networks_RH_Cont_PFCl_4 | Cont | 269 | 100 | OK |
| 7Networks_RH_Cont_PFCl_5 | Cont | 269 | 100 | OK |
| 7Networks_RH_Cont_PFCl_6 | Cont | 269 | 100 | OK |
| 7Networks_RH_Cont_PFCl_7 | Cont | 269 | 100 | OK |
| 7Networks_RH_Cont_pCun_1 | Cont | 269 | 100 | OK |
| 7Networks_RH_Cont_Cing_1 | Cont | 269 | 100 | OK |
| 7Networks_RH_Cont_Cing_2 | Cont | 269 | 100 | OK |
| 7Networks_RH_Cont_PFCmp_1 | Cont | 269 | 100 | OK |

| | | | | |
|---|---|---|---|---|
| 7Networks_RH_Cont_PFCmp_2 | Cont | 269 | 100 | OK |
| 7Networks_LH_Default_Temp_1 | Default | 229 | 85 | OK |
| 7Networks_LH_Default_Temp_2 | Default | 67 | 24 | POOR |
| 7Networks_LH_Default_Temp_3 | Default | 269 | 100 | OK |
| 7Networks_LH_Default_Temp_4 | Default | 269 | 100 | OK |
| 7Networks_LH_Default_Temp_5 | Default | 269 | 100 | OK |
| 7Networks_LH_Default_Par_1 | Default | 269 | 100 | OK |
| 7Networks_LH_Default_Par_2 | Default | 269 | 100 | OK |
| 7Networks_LH_Default_Par_3 | Default | 269 | 100 | OK |
| 7Networks_LH_Default_Par_4 | Default | 269 | 100 | OK |
| 7Networks_LH_Default_PFC_1 | Default | 269 | 100 | OK |
| 7Networks_LH_Default_PFC_2 | Default | 269 | 100 | OK |
| 7Networks_LH_Default_PFC_3 | Default | 269 | 100 | OK |
| 7Networks_LH_Default_PFC_4 | Default | 127 | 47 | POOR |
| 7Networks_LH_Default_PFC_5 | Default | 269 | 100 | OK |
| 7Networks_LH_Default_PFC_6 | Default | 269 | 100 | OK |
| 7Networks_LH_Default_PFC_7 | Default | 269 | 100 | OK |
| 7Networks_LH_Default_PFC_8 | Default | 269 | 100 | OK |
| 7Networks_LH_Default_PFC_9 | Default | 269 | 100 | OK |
| 7Networks_LH_Default_PFC_10 | Default | 269 | 100 | OK |
| 7Networks_LH_Default_PFC_11 | Default | 269 | 100 | OK |
| 7Networks_LH_Default_PFC_12 | Default | 269 | 100 | OK |
| 7Networks_LH_Default_PFC_13 | Default | 269 | 100 | OK |
| 7Networks_LH_Default_pCunPCC_1 | Default | 269 | 100 | OK |
| 7Networks_LH_Default_pCunPCC_2 | Default | 269 | 100 | OK |
| 7Networks_LH_Default_pCunPCC_3 | Default | 269 | 100 | OK |
| 7Networks_LH_Default_pCunPCC_4 | Default | 269 | 100 | OK |
| 7Networks_LH_Default_PHC_1 | Default | 269 | 100 | OK |
| 7Networks_RH_Default_Par_1 | Default | 269 | 100 | OK |
| 7Networks_RH_Default_Par_2 | Default | 269 | 100 | OK |

| | | | | |
|---|---|---|---|---|
| 7Networks_RH_Default_Par_3 | Default | 269 | 100 | OK |
| 7Networks_RH_Default_Temp_1 | Default | 269 | 100 | OK |
| 7Networks_RH_Default_Temp_2 | Default | 190 | 71 | OK |
| 7Networks_RH_Default_Temp_3 | Default | 269 | 100 | OK |
| 7Networks_RH_Default_Temp_4 | Default | 269 | 100 | OK |
| 7Networks_RH_Default_Temp_5 | Default | 269 | 100 | OK |
| 7Networks_RH_Default_PFCv_1 | Default | 269 | 100 | OK |
| 7Networks_RH_Default_PFCdPFCm_1 | Default | 249 | 93 | OK |
| 7Networks_RH_Default_PFCdPFCm_2 | Default | 269 | 100 | OK |
| 7Networks_RH_Default_PFCdPFCm_3 | Default | 269 | 100 | OK |
| 7Networks_RH_Default_PFCdPFCm_4 | Default | 269 | 100 | OK |
| 7Networks_RH_Default_PFCdPFCm_5 | Default | 269 | 100 | OK |
| 7Networks_RH_Default_PFCdPFCm_6 | Default | 269 | 100 | OK |
| 7Networks_RH_Default_PFCdPFCm_7 | Default | 269 | 100 | OK |
| 7Networks_RH_Default_pCunPCC_1 | Default | 269 | 100 | OK |
| 7Networks_RH_Default_pCunPCC_2 | Default | 269 | 100 | OK |
| 7Networks_RH_Default_pCunPCC_3 | Default | 269 | 100 | OK |
| 7Networks_LH_DorsAttn_Post_1 | DorsAttn | 269 | 100 | OK |
| 7Networks_LH_DorsAttn_Post_2 | DorsAttn | 269 | 100 | OK |
| 7Networks_LH_DorsAttn_Post_3 | DorsAttn | 269 | 100 | OK |
| 7Networks_LH_DorsAttn_Post_4 | DorsAttn | 269 | 100 | OK |
| 7Networks_LH_DorsAttn_Post_5 | DorsAttn | 269 | 100 | OK |
| 7Networks_LH_DorsAttn_Post_6 | DorsAttn | 269 | 100 | OK |
| 7Networks_LH_DorsAttn_Post_7 | DorsAttn | 269 | 100 | OK |
| 7Networks_LH_DorsAttn_Post_8 | DorsAttn | 269 | 100 | OK |
| 7Networks_LH_DorsAttn_Post_9 | DorsAttn | 269 | 100 | OK |
| 7Networks_LH_DorsAttn_Post_10 | DorsAttn | 269 | 100 | OK |
| 7Networks_LH_DorsAttn_FEF_1 | DorsAttn | 269 | 100 | OK |
| 7Networks_LH_DorsAttn_FEF_2 | DorsAttn | 269 | 100 | OK |
| 7Networks_LH_DorsAttn_PrCv_1 | DorsAttn | 269 | 100 | OK |

| | | | | |
|---|---|---|---|---|
| 7Networks_RH_DorsAttn_Post_1 | DorsAttn | 263 | 98 | OK |
| 7Networks_RH_DorsAttn_Post_2 | DorsAttn | 269 | 100 | OK |
| 7Networks_RH_DorsAttn_Post_3 | DorsAttn | 269 | 100 | OK |
| 7Networks_RH_DorsAttn_Post_4 | DorsAttn | 269 | 100 | OK |
| 7Networks_RH_DorsAttn_Post_5 | DorsAttn | 269 | 100 | OK |
| 7Networks_RH_DorsAttn_Post_6 | DorsAttn | 269 | 100 | OK |
| 7Networks_RH_DorsAttn_Post_7 | DorsAttn | 269 | 100 | OK |
| 7Networks_RH_DorsAttn_Post_8 | DorsAttn | 269 | 100 | OK |
| 7Networks_RH_DorsAttn_Post_9 | DorsAttn | 269 | 100 | OK |
| 7Networks_RH_DorsAttn_Post_10 | DorsAttn | 269 | 100 | OK |
| 7Networks_RH_DorsAttn_FEF_1 | DorsAttn | 269 | 100 | OK |
| 7Networks_RH_DorsAttn_FEF_2 | DorsAttn | 269 | 100 | OK |
| 7Networks_RH_DorsAttn_PrCv_1 | DorsAttn | 269 | 100 | OK |
| 7Networks_LH_Limbic_OFC_1 | Limbic | 58 | 22 | POOR |
| 7Networks_LH_Limbic_OFC_2 | Limbic | 0 | 0 | ZERO |
| 7Networks_LH_Limbic_TempPole_1 | Limbic | 30 | 11 | POOR |
| 7Networks_LH_Limbic_TempPole_2 | Limbic | 0 | 0 | ZERO |
| 7Networks_LH_Limbic_TempPole_3 | Limbic | 62 | 23 | POOR |
| 7Networks_LH_Limbic_TempPole_4 | Limbic | 269 | 100 | OK |
| 7Networks_RH_Limbic_OFC_1 | Limbic | 0 | 0 | ZERO |
| 7Networks_RH_Limbic_OFC_2 | Limbic | 164 | 61 | OK |
| 7Networks_RH_Limbic_OFC_3 | Limbic | 106 | 39 | POOR |
| 7Networks_RH_Limbic_TempPole_1 | Limbic | 84 | 31 | POOR |
| 7Networks_RH_Limbic_TempPole_2 | Limbic | 0 | 0 | ZERO |
| 7Networks_RH_Limbic_TempPole_3 | Limbic | 103 | 38 | POOR |
| 7Networks_LH_SalVentAttn_ParOper_1 | SalVentAttn | 269 | 100 | OK |
| 7Networks_LH_SalVentAttn_ParOper_2 | SalVentAttn | 269 | 100 | OK |
| 7Networks_LH_SalVentAttn_ParOper_3 | SalVentAttn | 269 | 100 | OK |
| 7Networks_LH_SalVentAttn_FrOperIns_1 | SalVentAttn | 269 | 100 | OK |
| 7Networks_LH_SalVentAttn_FrOperIns_2 | SalVentAttn | 269 | 100 | OK |

| | | | | |
|---|---|---|---|---|
| 7Networks_LH_SalVentAttn_FrOperIns_3 | SalVentAttn | 269 | 100 | OK |
| 7Networks_LH_SalVentAttn_FrOperIns_4 | SalVentAttn | 269 | 100 | OK |
| 7Networks_LH_SalVentAttn_PFCl_1 | SalVentAttn | 269 | 100 | OK |
| 7Networks_LH_SalVentAttn_Med_1 | SalVentAttn | 269 | 100 | OK |
| 7Networks_LH_SalVentAttn_Med_2 | SalVentAttn | 269 | 100 | OK |
| 7Networks_LH_SalVentAttn_Med_3 | SalVentAttn | 269 | 100 | OK |
| 7Networks_RH_SalVentAttn_TempOccPar_ | SalVentAttn | 269 | 100 | OK |
| 7Networks_RH_SalVentAttn_TempOccPar_ | SalVentAttn | 269 | 100 | OK |
| 7Networks_RH_SalVentAttn_TempOccPar_ | SalVentAttn | 269 | 100 | OK |
| 7Networks_RH_SalVentAttn_PrC_1 | SalVentAttn | 269 | 100 | OK |
| 7Networks_RH_SalVentAttn_FrOperIns_1 | SalVentAttn | 269 | 100 | OK |
| 7Networks_RH_SalVentAttn_FrOperIns_2 | SalVentAttn | 269 | 100 | OK |
| 7Networks_RH_SalVentAttn_FrOperIns_3 | SalVentAttn | 269 | 100 | OK |
| 7Networks_RH_SalVentAttn_FrOperIns_4 | SalVentAttn | 269 | 100 | OK |
| 7Networks_RH_SalVentAttn_Med_1 | SalVentAttn | 269 | 100 | OK |
| 7Networks_RH_SalVentAttn_Med_2 | SalVentAttn | 269 | 100 | OK |
| 7Networks_RH_SalVentAttn_Med_3 | SalVentAttn | 269 | 100 | OK |
| 7Networks_LH_SomMot_1 | SomMot | 269 | 100 | OK |
| 7Networks_LH_SomMot_2 | SomMot | 269 | 100 | OK |
| 7Networks_LH_SomMot_3 | SomMot | 269 | 100 | OK |
| 7Networks_LH_SomMot_4 | SomMot | 269 | 100 | OK |
| 7Networks_LH_SomMot_5 | SomMot | 269 | 100 | OK |
| 7Networks_LH_SomMot_6 | SomMot | 269 | 100 | OK |
| 7Networks_LH_SomMot_7 | SomMot | 269 | 100 | OK |
| 7Networks_LH_SomMot_8 | SomMot | 269 | 100 | OK |
| 7Networks_LH_SomMot_9 | SomMot | 269 | 100 | OK |
| 7Networks_LH_SomMot_10 | SomMot | 269 | 100 | OK |
| 7Networks_LH_SomMot_11 | SomMot | 269 | 100 | OK |
| 7Networks_LH_SomMot_12 | SomMot | 269 | 100 | OK |
| 7Networks_LH_SomMot_13 | SomMot | 269 | 100 | OK |

| | | | | |
|---|---|---|---|---|
| 7Networks_LH_SomMot_14 | SomMot | 269 | 100 | OK |
| 7Networks_LH_SomMot_15 | SomMot | 269 | 100 | OK |
| 7Networks_LH_SomMot_16 | SomMot | 269 | 100 | OK |
| 7Networks_RH_SomMot_1 | SomMot | 269 | 100 | OK |
| 7Networks_RH_SomMot_2 | SomMot | 269 | 100 | OK |
| 7Networks_RH_SomMot_3 | SomMot | 269 | 100 | OK |
| 7Networks_RH_SomMot_4 | SomMot | 269 | 100 | OK |
| 7Networks_RH_SomMot_5 | SomMot | 269 | 100 | OK |
| 7Networks_RH_SomMot_6 | SomMot | 269 | 100 | OK |
| 7Networks_RH_SomMot_7 | SomMot | 269 | 100 | OK |
| 7Networks_RH_SomMot_8 | SomMot | 269 | 100 | OK |
| 7Networks_RH_SomMot_9 | SomMot | 269 | 100 | OK |
| 7Networks_RH_SomMot_10 | SomMot | 269 | 100 | OK |
| 7Networks_RH_SomMot_11 | SomMot | 269 | 100 | OK |
| 7Networks_RH_SomMot_12 | SomMot | 269 | 100 | OK |
| 7Networks_RH_SomMot_13 | SomMot | 269 | 100 | OK |
| 7Networks_RH_SomMot_14 | SomMot | 269 | 100 | OK |
| 7Networks_RH_SomMot_15 | SomMot | 269 | 100 | OK |
| 7Networks_RH_SomMot_16 | SomMot | 269 | 100 | OK |
| 7Networks_RH_SomMot_17 | SomMot | 269 | 100 | OK |
| 7Networks_RH_SomMot_18 | SomMot | 269 | 100 | OK |
| 7Networks_RH_SomMot_19 | SomMot | 269 | 100 | OK |
| 7Networks_LH_Vis_1 | Vis | 269 | 100 | OK |
| 7Networks_LH_Vis_2 | Vis | 269 | 100 | OK |
| 7Networks_LH_Vis_3 | Vis | 269 | 100 | OK |
| 7Networks_LH_Vis_4 | Vis | 269 | 100 | OK |
| 7Networks_LH_Vis_5 | Vis | 163 | 61 | OK |
| 7Networks_LH_Vis_6 | Vis | 269 | 100 | OK |
| 7Networks_LH_Vis_7 | Vis | 269 | 100 | OK |
| 7Networks_LH_Vis_8 | Vis | 269 | 100 | OK |

| | | | | |
|---|---|---|---|---|
| 7Networks_LH_Vis_9 | Vis | 269 | 100 | OK |
| 7Networks_LH_Vis_10 | Vis | 269 | 100 | OK |
| 7Networks_LH_Vis_11 | Vis | 269 | 100 | OK |
| 7Networks_LH_Vis_12 | Vis | 269 | 100 | OK |
| 7Networks_LH_Vis_13 | Vis | 269 | 100 | OK |
| 7Networks_LH_Vis_14 | Vis | 269 | 100 | OK |
| 7Networks_RH_Vis_1 | Vis | 239 | 89 | OK |
| 7Networks_RH_Vis_2 | Vis | 269 | 100 | OK |
| 7Networks_RH_Vis_3 | Vis | 269 | 100 | OK |
| 7Networks_RH_Vis_4 | Vis | 269 | 100 | OK |
| 7Networks_RH_Vis_5 | Vis | 269 | 100 | OK |
| 7Networks_RH_Vis_6 | Vis | 269 | 100 | OK |
| 7Networks_RH_Vis_7 | Vis | 269 | 100 | OK |
| 7Networks_RH_Vis_8 | Vis | 245 | 91 | OK |
| 7Networks_RH_Vis_9 | Vis | 269 | 100 | OK |
| 7Networks_RH_Vis_10 | Vis | 269 | 100 | OK |
| 7Networks_RH_Vis_11 | Vis | 269 | 100 | OK |
| 7Networks_RH_Vis_12 | Vis | 269 | 100 | OK |
| 7Networks_RH_Vis_13 | Vis | 269 | 100 | OK |
| 7Networks_RH_Vis_14 | Vis | 267 | 99 | OK |
| 7Networks_RH_Vis_15 | Vis | 269 | 100 | OK |
