## Supplemental Table 14 for "Psychedelics distinctly alter brain entropy and complexity compared to psychostimulants"

| network | n_regions_total | mean_coverage | min_coverage | max_coverage | survival_rate_pc | n_sessions_below_threshold |
|---|---|---|---|---|---|---|
| **Cont** | 30 | 29 | 27 | 30 | 96 | 0 |
| **Default** | 46 | 44 | 41 | 46 | 96 | 0 |
| **DorsAttn** | 26 | 26 | 25 | 26 | 99 | 0 |
| **Limbic** | 12 | 33 | 1 | 8 | 27 | 120 |
| **SalVentAttn** | 22 | 22 | 22 | 22 | 100 | 0 |
| **SomMot** | 35 | 35 | 35 | 35 | 100 | 0 |
| **Vis** | 29 | 28 | 27 | 29 | 98 | 0 |
