## Supplemental Table 16 for "Psychedelics distinctly alter brain entropy and complexity compared to psychostimulants"

| Variable | Condition | P1 | P2 | P3 |
|---|---|---|---|---|
| *Lempel-Ziv complexity* | | | | |
| | Placebo | 0.632 ± 0.011 | 0.638 ± 0.011 | 0.630 ± 0.013 |
| | LSD | 0.638 ± 0.011 | 0.644 ± 0.008 | 0.642 ± 0.009 |
| *Global NSC* | | | | |
| | Placebo | 0.698 ± 0.012 | 0.698 ± 0.013 | 0.698 ± 0.016 |
| | LSD | 0.707 ± 0.021 | 0.711 ± 0.018 | 0.712 ± 0.021 |
| *Meta-state complexity* | | | | |
| | Placebo | 0.857 ± 0.062 | 0.847 ± 0.074 | 0.822 ± 0.062 |
| | LSD | 0.849 ± 0.046 | 0.857 ± 0.067 | 0.863 ± 0.079 |
| *Absolute modularity* | | | | |
| | Placebo | 0.287 ± 0.079 | 0.307 ± 0.078 | 0.296 ± 0.074 |
| | LSD | 0.163 ± 0.076 | 0.186 ± 0.074 | 0.179 ± 0.080 |
